# Identifying molecular mediators of the relationship between body mass index and endometrial cancer risk: a Mendelian randomization analysis

**DOI:** 10.1101/2021.12.10.21267599

**Authors:** Emma Hazelwood, Eleanor Sanderson, Vanessa Y Tan, Katherine S Ruth, Timothy M Frayling, Niki Dimou, Marc J Gunter, Laure Dossus, Claire Newton, Neil Ryan, Dimitri J Pournaras, Tracy A O’Mara, George Davey Smith, Richard M Martin, James Yarmolinsky

**Author notes:** Corresponding author: James Yarmolinsky, PhD, MRC Integrative Epidemiology Unit, Population Health Sciences Bristol Medical School, University of Bristol Bristol, UK.

## Abstract

**Background:** Endometrial cancer is the most common gynaecological cancer in high-income countries. Elevated body mass index (BMI) is an established modifiable risk factor for this condition and is estimated to confer a larger effect on endometrial cancer risk than any other cancer site. However, the molecular mechanisms underpinning this association remain unclear. We used Mendelian randomization (MR) to evaluate the causal role of 14 molecular risk factors (hormonal, metabolic, and inflammatory markers) in endometrial cancer risk. We then evaluated and quantified the potential mediating role of these molecular traits in the relationship between BMI and endometrial cancer.

**Methods and Findings:** Genetic instruments to proxy 14 molecular risk factors and BMI were constructed by identifying single-nucleotide polymorphisms (SNPs) reliably associated (*P* < 5.0 x 10^-8^) with each respective risk factor in previous genome-wide association studies (GWAS). Summary statistics for the association of these SNPs with overall and subtype-specific endometrial cancer risk (12,906 cases and 108,979 controls) were obtained from a GWAS meta-analysis of the Endometrial Cancer Association Consortium (ECAC), Epidemiology of Endometrial Cancer Consortium (E2C2), and UK Biobank. SNPs were combined into multi-allelic models and odds ratios (ORs) and 95% confidence intervals (95% CIs) were generated using inverse-variance weighted random-effects models. The mediating roles of the molecular risk factors in the relationship between BMI and endometrial cancer were then estimated using multivariable MR. In MR analyses, there was strong evidence that BMI (OR per SD increase: 1.88, 95% CI: 1.69 to 2.09, *P* = 3.87 x 10^-31^), total testosterone (OR per inverse normal transformed nmol/L increase: 1.64, 95% CI: 1.43 to 1.88, *P* = 1.71 x 10^-12^), bioavailable testosterone (OR per inverse normal transformed nmol/L increase: 1.46, 95% CI: 1.29 to 1.65, *P* = 3.48 x 10^-9^), fasting insulin (OR per natural log transformed pmol/L increase: 3.93, 95% CI: 2.29 to 6.74, *P* = 7.18 x 10^-7^) and sex hormone-binding globulin (SHBG, OR per inverse normal transformed nmol/L increase: 0.71, 95% CI: 0.59 to 0.85, *P* = 2.07 x 10^-4^) had a causal effect on endometrial cancer risk. Additionally, there was suggestive evidence that total serum cholesterol (OR per mg/dL increase: 0.90, 95% CI: 0.81 to 1.00, *P* = 4.01 x 10^-2^) had an effect on endometrial cancer risk. In mediation analysis using multivariable MR, we found evidence for a mediating role of fasting insulin (19% total effect mediated, 95% CI: 5 to 34%, *P* = 9.17 x 10^-3^), bioavailable testosterone (15% mediated, 95% CI: 10 to 20%, *P* = 1.43 x 10^-8^), and SHBG (7% mediated, 95% CI: 1 to 12%, *P* = 1.81 x 10^-2^) in the relationship between BMI and endometrial cancer risk. The primary limitations of this analysis include the assumption of linear relationships across univariable and multivariable analyses and the restriction of analyses to individuals of European ancestry.

**Conclusions:** Our comprehensive Mendelian randomization analysis provides insight into potential causal mechanisms linking BMI with endometrial cancer risk and suggests pharmacological targeting of insulinemic and hormonal traits as a potential strategy for the prevention of endometrial cancer.

## Introduction

Endometrial cancer is the most common gynaecological cancer in high-income countries and the second most common globally [1, 2]. In 2020, there were 417,367 new cases diagnosed and 97,370 endometrial cancer-related deaths worldwide [3]. In contrast to several other cancer types where incidence rates have been declining over the past two decades, the global incidence of endometrial cancer continues to increase [4–8].

Elevated body mass index (BMI) is an established risk factor for endometrial cancer and is estimated to confer a larger effect on risk of this malignancy than any other cancer type [9–11]. A recent meta-analysis of 30 prospective studies reported that each 5 kg/m^2^ increase in BMI was associated with a 54% (95% CI: 47 to 61%) higher risk of endometrial cancer [12–14]. It is estimated that excess adiposity accounts for 34% of global endometrial cancer cases, with the increasing incidence of endometrial cancer mirroring rising levels of obesity worldwide [15–17]. Lifestyle and dietary interventions encouraging maintenance of a healthy weight therefore remain cornerstones for the primary prevention of endometrial cancer [9]. Alongside weight management strategies, greater characterisation of the molecular mechanisms underpinning an effect of excess adiposity on endometrial cancer could provide a complementary approach to cancer prevention through the development of pharmacological interventions targeting these traits in high-risk groups.

Observational epidemiological studies have reported associations between several hormonal, metabolic, and inflammatory factors linked to obesity and endometrial cancer, including bioavailable testosterone, sex hormone-binding globulin (SHBG), oestradiol and fasting insulin [18–22]. However, conventional observational studies are susceptible to residual confounding (due to unmeasured or imprecisely measured confounders), reverse causation, and other forms of bias which undermine robust causal inference. Therefore, the causal nature of these risk factors, and thus their suitability as effective intervention targets for endometrial cancer prevention, remains unclear.

Mendelian randomization (MR) is an analytical approach that uses germline genetic variants as instruments (“proxies”) for risk factors to evaluate the causal effects of these factors on disease outcomes in observational settings [23, 24]. Since germline genetic variants are randomly assorted at meiosis, MR analyses should be less prone to confounding by lifestyle and environmental factors than conventional observational studies. Furthermore, since germline genetic variants are fixed at conception, MR analyses are not subject to reverse causation bias. The statistical power and precision of MR analysis can be increased by employing a “two-sample MR” framework in which summary genetic association data from two independent samples – one representing genetic variant-exposure associations and one representing genetic variant-outcome associations – are synthesised to estimate causal effects [25].

Recent MR studies have suggested potential causal relationships between circulating levels of several molecular traits, including low-density lipoprotein (LDL) cholesterol, insulin, total and bioavailable testosterone, and sex hormone-binding globulin (SHBG) and endometrial cancer risk, and have confirmed a causal role of BMI in endometrial cancer risk [17, 26–32]. However, many previously reported molecular risk factors for endometrial cancer from conventional observational studies remain untested in an MR framework, meaning the causal relevance of these factors in disease onset is unclear. Additionally, no MR studies to date have attempted to quantify the potential mediating role of these factors in the relationship between BMI and endometrial cancer risk.

Given the unclear causal relevance of previously reported molecular traits in endometrial cancer aetiology, we used a two-sample MR approach to evaluate the causal role of 14 endogenous sex hormones, metabolic traits, and inflammatory markers in endometrial cancer risk (overall and in endometrioid and non-endometrioid subtypes). We then used multivariable MR to evaluate and quantify the mediating role of these molecular traits in the relationship between BMI and endometrial cancer risk.

## Methods

Our analytical strategy was as follows: first, we attempted to corroborate previous MR findings that there was evidence of a causal relationship between BMI and endometrial cancer risk (overall and by histological subtype); second, we examined evidence for a causal relationship between previously reported molecular factors and endometrial cancer risk (overall and by histological subtype); third, we evaluated the causal relationship between BMI and those molecular risk factors that were confirmed to influence endometrial cancer risk (overall and by histological subtype); finally, we performed a mediation analysis to quantify the proportion of the total effect of BMI on endometrial cancer risk that was mediated by each identified trait.

### Endometrial cancer study population

Summary genetic association data on overall and subtype-specific endometrial cancer risk were obtained from a genome-wide association study (GWAS) of 12,906 cases (including 8,758 endometrioid and 1,230 non-endometrioid endometrial cancer cases) and up to 108,979 controls of European ancestry [31]. This meta-GWAS combined 17 previously reported studies from the Endometrial Cancer Association Consortium (ECAC), the Epidemiology of Endometrial Cancer Consortium (E2C2), and UK Biobank, with four studies contributing samples to more than one genotyping project. Participants were recruited from Australia, Belgium, Germany, Poland, Sweden, the UK, and the USA and associations were adjusted for principal components of ancestry. Genotyping was performed using one of several Illumina arrays and imputation was performed using the 1000 Genomes Phase 3 reference panel [33]. Further information on this meta-GWAS including study-specific genotyping, imputation, and quality control procedures is provided in S2 Appendix.

### Identification of previously reported molecular risk factors for endometrial cancer

We performed two pragmatic searches of the literature using PubMed. The first search identified previously published MR analyses of molecular risk factors for endometrial cancer. The second search identified narrative or systematic reviews of potential molecular mechanisms underpinning the relationship between obesity and endometrial cancer (additional information on search strategies used in literature reviews is presented in **S3 Appendix**). Combined, these literature reviews identified 20 unique molecular traits which could mediate the effect of BMI on endometrial cancer risk, of which 14 had suitable genetic instruments available. These traits include nine metabolic factors (low-density lipoprotein (LDL) cholesterol, high-density lipoprotein (HDL) cholesterol, total serum cholesterol, triglycerides, blood glucose, fasting insulin, insulin-like growth factor 1 (IGF-1), adiponectin, and leptin); three endogenous sex hormones or traits that regulate their bioactivity (total and bioavailable testosterone, and SHBG); and two inflammatory markers (interleukin-6 (IL-6) and C-reactive protein (CRP), measured as high-sensitivity CRP) (**Fig 1**) [29, 34–41]. Summary genetic association data on BMI were obtained from a GWAS of 681,275 individuals of European ancestry [42]. Additional information on participant demographics and covariates included in adjustment strategies across each GWAS are presented in **S4 Table**. All studies contributing data to these analyses had the relevant institutional review board approval from each country, in accordance with the Declaration of Helsinki, and all participants provided informed consent.

**Fig 1.**
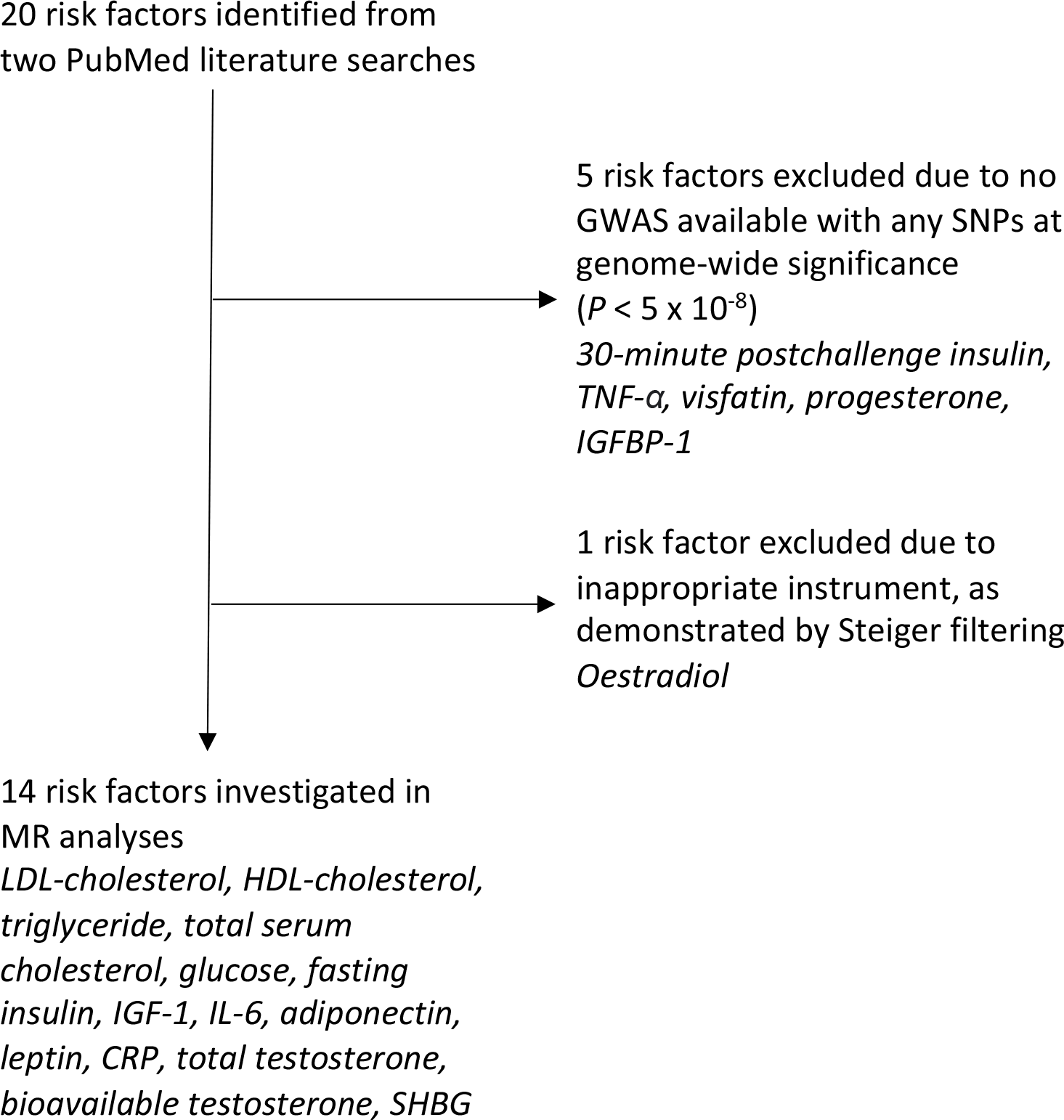
Flowchart detailing the process of identifying previously reported risk factors with suitable genetic instruments. TNF-α = tumour necrosis factor-α, IGFBP-1 = insulin-like growth factor-binding protein-1, LDL = low-density lipoprotein, HDL = high-density lipoprotein, IGF-1 = insulin-like growth factor-1, IL-6 = interleukin-6, CRP = C-reactive protein, SHBG = sex hormone-binding globulin.

### Statistical analyses

MR analysis can generate unbiased estimates of causal effects of risk factors on disease outcomes if the following assumptions are met: (i) the instrument strongly associates with the exposure (“relevance”), (ii) there is no confounding of the instrument-outcome relationship (“exchangeability”), and (iii) the instrument only affects the outcome through the exposure (“exclusion restriction”) (**Fig. 2**) [43].

**Fig 2.**
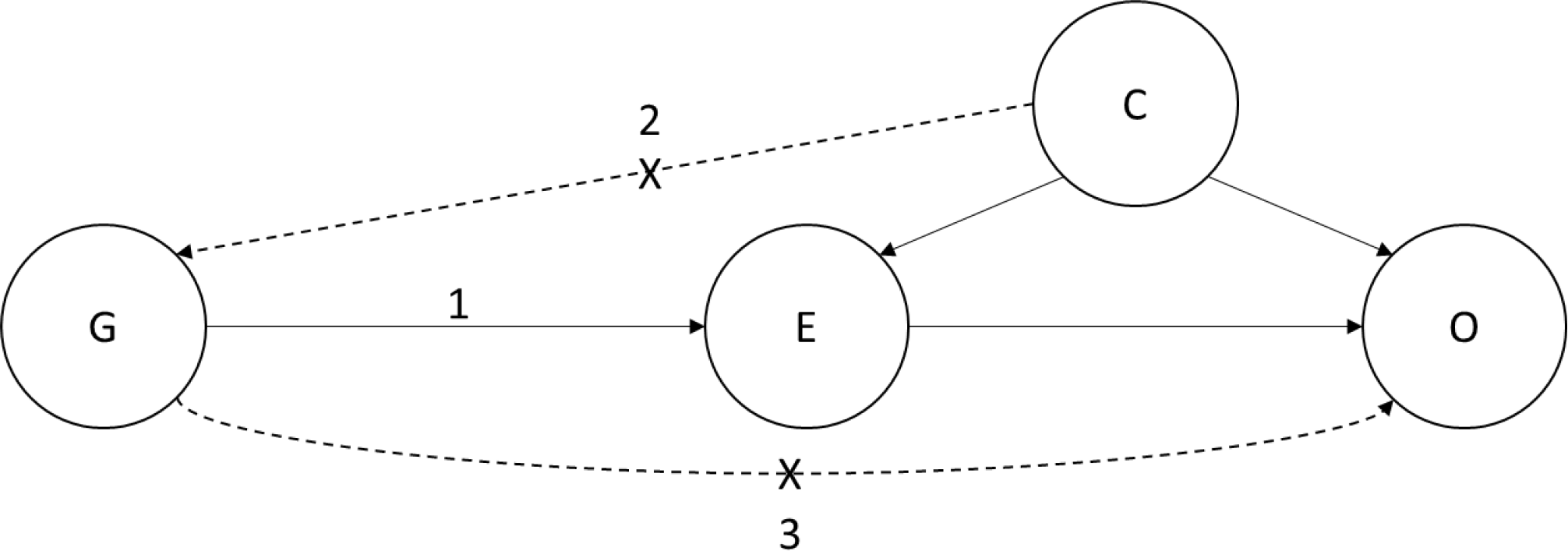
DAG demonstrating the core assumptions of Mendelian randomization. DAG = directed acyclic diagram, G = genetic instrument, E = exposure, O = outcome, C = confounding factors. Arrows labelled 1, 2, and 3 represent the three core assumptions of MR: (1) the instrument strongly associates with the exposure (“relevance”); (2) there is no confounding of the instrument-outcome relationship (“exchangeability”); and (3) the instrument only affects the outcome through the exposure (“exclusion restriction”). MR uses genetic instruments to proxy exposures in order to strengthen causal inference in observational epidemiological settings. As these genetic instruments are randomly inherited at meiosis, they should not be affected by conventional confounding factors like environmental, lifestyle, and behavioural traits. In addition, since germline genetic variants are fixed at conception and cannot be altered by subsequent exposures, they are not susceptible to reverse causation. Finally, germline genotype can be measured relatively precisely using modern genotyping technologies which minimises measurement error. Collectively, these properties of germline genetic variants (along with technologies that measure them) permit MR analyses to minimise many of the sources of bias which can undermine robust causal inference in conventional observational epidemiological analyses.

To construct genetic instruments for BMI and previously reported molecular risk factors, we obtained single-nucleotide polymorphisms (SNPs) reliably (*P* < 5 x 10^-8^) and independently (r^2^ < 0.001) associated with each trait. To construct a genetic instrument for leptin, we restricted genetic variants to *cis*-acting SNPs (i.e. in or within ±100kb from the gene encoding the protein). For leptin, IL-6 and CRP analyses, SNPs were permitted to be in weak linkage disequilibrium (LD) (r^2^ < 0.10) to maximise instrument strength. For all traits where instruments consisted of SNPs in weak LD (i.e. leptin, IL-6 and CRP), standard errors for causal estimates were inflated to account for correlation between SNPs with reference to the 1000 Genomes Phase 3 reference panel [33, 44].

For traits instrumented by a single SNP, the Wald ratio was used to generate effect estimates and the delta method was used to approximate standard errors [45]. For traits instrumented by two or more SNPs, inverse-variance weighted (IVW) random-effects models were used to estimate causal effects [45]. A Bonferroni correction was applied as a heuristic to account for multiple testing in MR analyses for the 15 risk factors (14 molecular traits and BMI) investigated. Results below this threshold were classified as “strong evidence” (*P* < 3.33 x 10^-3^ (0.05/15 traits)), whereas results between this threshold and *P* < 0.05 were classified as “suggestive evidence”.

When using genetic instruments, there is potential for horizontal pleiotropy - when a genetic variant influences an outcome through a biological pathway independent to the exposure, a violation of the “exclusion restriction” criterion [46]. We evaluated the presence of horizontal pleiotropy by performing various sensitivity analyses. First, for instruments consisting of ≥ 10 SNPs, we re-calculated causal estimates obtained from IVW models using MR-Egger regression, weighted median estimation, and weighted mode estimation (additional information on these sensitivity analyses is provided in **S2 Appendix**) [46–48]. Each of these models makes different assumptions regarding the nature of horizontal pleiotropy in the genetic instrument and therefore performing all three can provide complementary support to IVW models in evaluating the presence of horizontal pleiotropy. These models were not employed when instruments consisted of < 10 SNPs because of their reduced statistical power to detect horizontal pleiotropy in these settings (additional information on these sensitivity analyses is provided in **S2 Appendix**). Second, we performed “leave-one-out” analyses for all findings showing strong or suggestive evidence of effects in IVW models (*P* < 0.05) for traits where instruments consisted of ≥ 10 SNPs and findings were consistent across MR-Egger, weighted median, and weighted mode sensitivity analyses or where instruments consisted of < 10 SNPs. This approach sequentially removes each SNP from an instrument and then re-calculates the overall effect estimate to examine robustness of findings to individual influential SNPs in IVW models.

Instruments were derived from sex-combined GWAS for all traits other than those related to endogenous sex hormones to maximise statistical power where there was limited evidence of sex-specificity of SNP associations. As a sensitivity analysis we also re-performed MR analyses using sex-specific instruments where possible. For BMI, all analyses with strong or suggestive evidence for an effect (*P* < 0.05) were repeated using genome-wide significant (*P* < 5.0 x 10^-8^) variants identified in female-specific analyses. Likewise, for fasting insulin and CRP analyses, the effect estimates and standard errors of SNPs used to instrument these traits were replaced with female-specific values where there was previous evidence of sex-specificity of associations (trait-specific criteria for identifying sex-specific effects are presented in **S2 Appendix**). Findings from sex-specific sensitivity analyses are presented in **Tables S8, S17, S28-29**. Finally, Steiger filtering was performed across all analyses to identify and subsequently remove any SNPs which explained more variance in the outcome than the exposure (i.e. suggesting misspecification of the causal direction between traits) [49].

### Mediation analysis

For all molecular traits that were identified as being on the causal pathway between BMI and endometrial cancer risk, we used multivariable MR to generate estimates of the direct effect (i.e. the remaining effect of the exposure on the outcome when the effect of the candidate mediator on the outcome has been adjusted for) and indirect effect (i.e. the effect of the exposure on the outcome through the candidate mediator) using the product of coefficients method [50]. The proportion of the total effect of BMI on endometrial cancer risk (“proportion mediated”) that was mediated by each molecular trait was calculated using these estimates. In the case of fasting insulin, due to weak instrument bias, several different approaches were employed to attempt to maximise conditional instrument strength (for further information on these analyses see **S2 Appendix**). Standard errors for the proportion mediated were calculated using the delta method [51]. In addition, we aimed to perform additional mediation analyses combining all mediators into a single model to examine the extent to which these mediators influenced endometrial cancer independently or via shared biological pathways (presumed relationships between BMI, fasting insulin, SHBG, bioavailable testosterone, and endometrial cancer risk are presented in **Fig 3**). When all putative mediators were combined into a single model with BMI, however, there was persistent weak instrument bias. Of various alternate approaches examined to minimise this bias, the restriction of models to pairs of mediators (without inclusion of BMI) was found to generate the largest conditional F-statistics for each mediator included in the model (for further information on these analyses see **S2 Appendix**).

**Fig 3.**
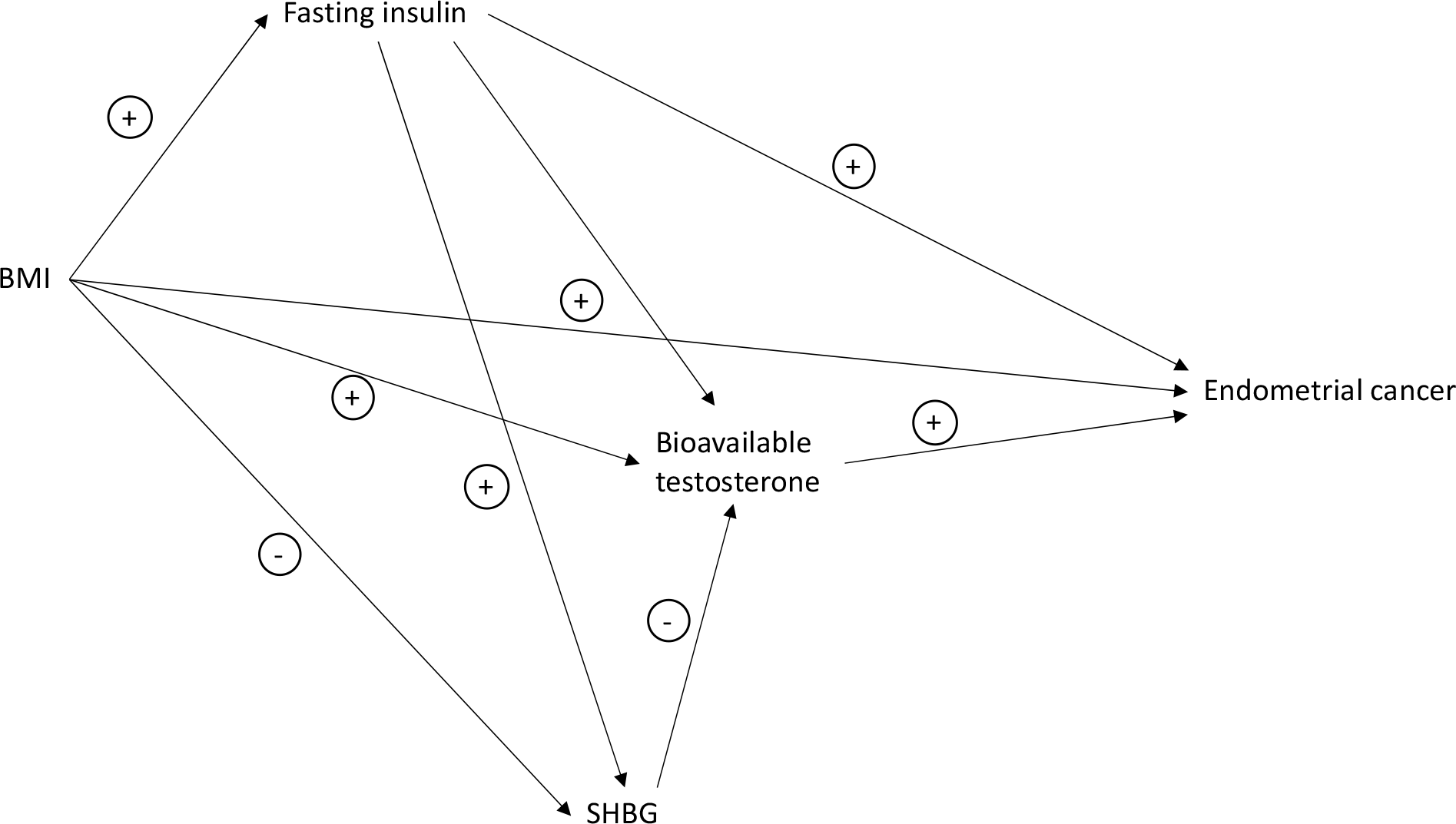
DAG demonstrating the proposed causal interactions of BMI, SHBG, fasting insulin, and bioavailable testosterone on endometrial cancer (overall and the endometrioid histological subtype). DAG = directed acyclic diagram, BMI = body mass index, SHBG = sex hormone-binding globulin.

### Sample overlap sensitivity analyses

There was moderate sample overlap (52.2-62.4%) across some analyses which can bias MR estimates toward the confounded observational estimate in the presence of weak instrument bias [52] (**S5 Table**). This bias can be inflated by “Winner’s curse”, in which weights for genetic instruments are derived from discovery samples that overlap with outcome samples. Though instruments in this analysis were constructed from genome-wide significant variants (*P* < 5.0 x 10^-8^) which should minimise the possibility of weak instrument bias, we performed the following sensitivity analyses to evaluate whether our findings could be influenced by sample overlap: i) for analyses examining the effect of blood glucose on endometrial cancer risk, we re-performed MR analyses using alternate GWAS data for this trait where there was no sample overlap [53, 54]; ii) for analyses examining the effect of BMI on total testosterone, bioavailable testosterone, SHBG and endometrial cancer, we re-performed MR analyses using alternate GWAS data for BMI where there was no sample overlap [55]; and iii) for analyses examining the effect of total testosterone, bioavailable testosterone, SHBG and IGF-1 on endometrial cancer (where suitable alternate GWAS data were not available), we re-constructed instruments for sex hormones using more conservative *P*-value thresholds (*P* < 5.0 x 10^-9^, *P* < 5.0 x 10^-10^) to minimise the potential for inclusion of weak instruments into analyses and then re-performed MR analyses. Similarly, in mediation analysis, due to the presence of sample overlap and possible influence of Winner’s curse, for any trait with sample overlap in the same multivariable MR model the analysis was repeated with a more stringent *P* value (*P* < 5.0 x 10^-9^) used for instrument construction.

All statistical analyses were performed using R (Vienna, Austria) version 4.0.2. Additional information on statistical packages used across various analyses is presented in **S2 Appendix**.

## Results

### Evaluating the effect of BMI on endometrial cancer risk

In MR analyses, there was strong evidence for an effect of BMI on risk of overall endometrial cancer (odds ratio (OR) per SD (4.7 kg/m^2^) increase in BMI: 1.88, 95% confidence interval (CI): 1.69 to 2.09, *P* = 3.87 x 10^-31^) (**Fig 4**, **Table 2**). This finding was consistent across sensitivity analyses examining evidence of horizontal pleiotropy, including MR-Egger, weighted median, and weighted mode models, in analyses using a female-specific BMI instrument, in analyses exploring potential Winner’s curse bias in instrument construction, and in the leave-one-out analysis (**S7 Figure, S8 Table, S9 Table**).

**Fig 4.**
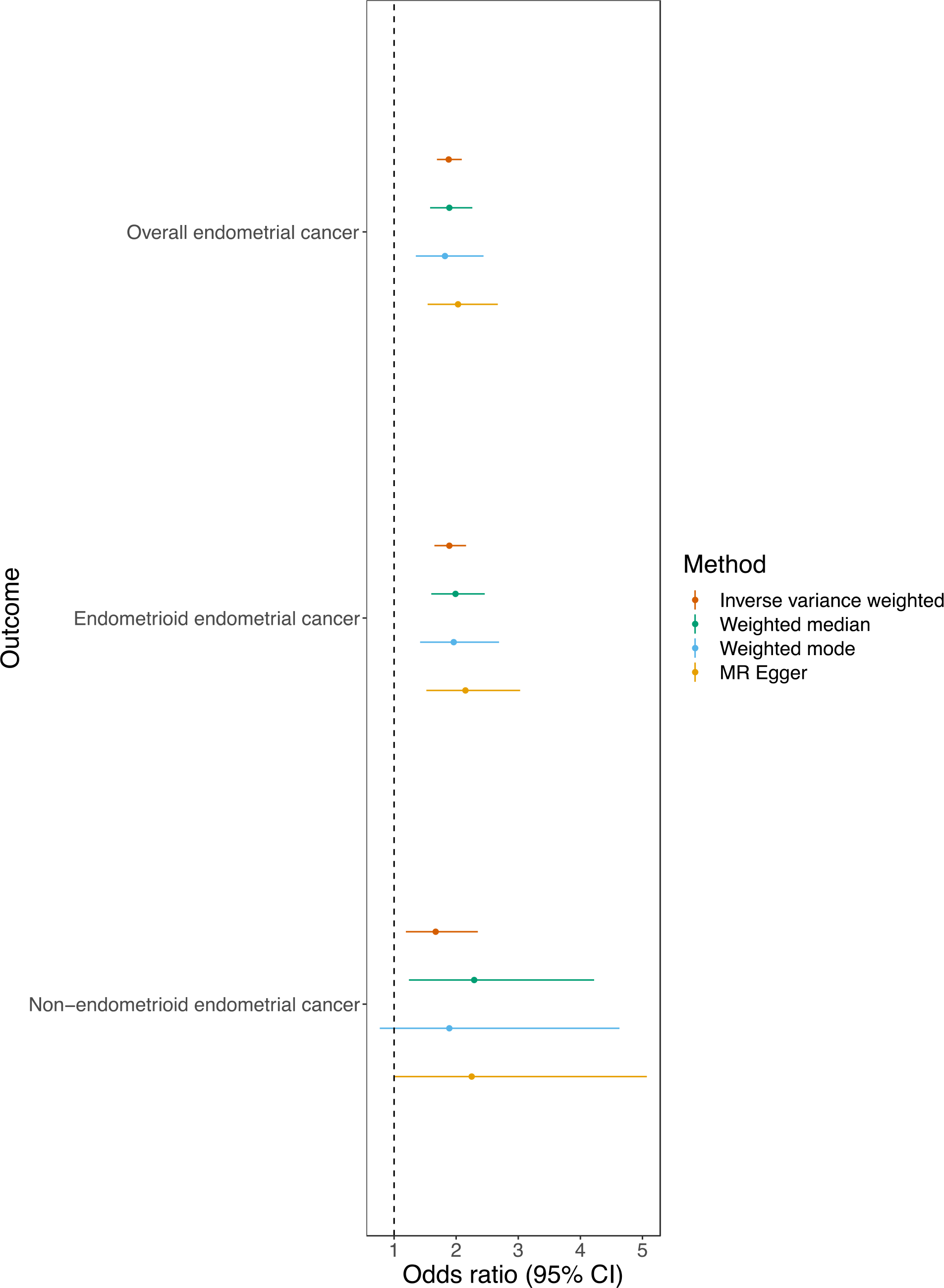
Mendelian randomization analysis of BMI on overall and subtype-specific endometrial cancer risk. Results of MR analyses examining the effect of adult BMI on risk of overall and subtype-specific endometrial cancer risk.

**Table 1.** Details of the instruments used for exposures.

| Exposure | GWAS | Sample size | Number of SNPs | R<sup>2</sup> | F-statistic | Sex-specificity |
| --- | --- | --- | --- | --- | --- | --- |
| Adult BMI | Yengo et al. [42] | 681,275 | 507 | 0.078 | 57,847 | Combined |
| LDL-cholesterol | Willer et al. [34] | 188,577 | 81 | 0.182 | 15,002 | Combined |
| HDL-cholesterol | Willer et al. [34] | 188,577 | 89 | 0.055 | 10,978 | Combined |
| Triglyceride | Willer et al. [34] | 188,577 | 55 | 0.052 | 9,811 | Combined |
| Total serum<br>cholesterol | Willer et al. [34] | 188,577 | 88 | 0.063 | 12,696 | Combined |
| Glucose | Neale et al. [36] | 361,194 | 109 | 0.036 | 11,776 | Female |
| Fasting insulin<br>(unadjusted for<br>BMI) | Lagou et al. [116] | 98,210 | 14 | 0.005 | 523 | Combined |
| Fasting insulin<br>(adjusted for<br>BMI) | Chen et al. [117] | 150,571 | 14 | 0.006 | 865 | Combined |
| IGF-1 (cis and<br>trans variants) | Sinnott-Armstrong<br>et al. [37] | 358,072 | 413 | 0.036 | 13,367 | Combined |
| IGF-1 (cis<br>variants) | Larsson et al.<br>[118] | 358,072 | 1 | 0.002 | 814 | Combined |
| IL-6 | Georgakis et al.<br>[38] | 204,402 | 7 | 0.004 | 911 | Combined |
| Adiponectin (cis<br>and trans<br>variants) | Locke et al. [119] | 14,172 | 3 | 0.023 | 328 | Combined |
| Adiponectin (cis<br>variants) | Locke et al. [119] | 14,172 | 3 | 0.023 | 334 | Combined |
| Leptin | Folkersen et al.<br>[120] | 30,931 | 1 | 0.001 | 34 | Combined |
| CRP (cis and<br>trans variants) | Ligthart et al. [41] | 204,402 | 45 | 0.035 | 7,414 | Combined |
| CRP (cis<br>variants) | C Reactive Protein<br>Coronary Heart<br>Disease Genetics<br>Collaboration<br>(CCGC) [121] | 105,476 | 4 | 0.010 | 1,030 | Combined |
| Total<br>testosterone | Ruth et al. [29] | 230,454 | 131 | 0.052 | 10,103 | Female |
| Bioavailable<br>testosterone | Ruth et al. [29] | 188,507 | 147 | 0.054 | 10,599 | Female |
| SHBG | Ruth et al. [29] | 189,473 | 215 | 0.122 | 26,286 | Female |

BMI is scaled to an SD increase (4.7 kg/m<sup>2</sup>). For the analysis involving the plasma proteome, due to the requirement of increased statistical power in order to overcome the multiple testing burden, alternative summary genetic association data for BMI were obtained from a genome-wide association study of 681,275 individuals of European ancestry (note that this summary genetic data could not be used for other analyses due to substantial overlap of participants with summary genetic data of other traits) [42]. The CRP GWAS included some individuals of non-European ancestry and adjusted for ancestry where applicable. BMI = body mass index, LDL = low-density lipoprotein, HDL = high density lipoprotein, IGF-1 = Insulin-like growth factor-1, IL-6 = interleukin-6, CRP = C-reactive protein, SHBG = sex hormone-binding globulin, LD = linkage disequilibrium. For instrument construction of IGF-1 (cis and trans variants), a *P* value of 5 x 10<sup>-6</sup> was used.

**Table 2.** Results of MR analyses examining the effect of adult BMI on risk of overall and subtype-specific endometrial cancer risk.

| Outcome | Method | OR (95% CI) | P value |
| --- | --- | --- | --- |
| Overall | IVW | 1.88 (1.69 to 2.09) | $3.87 \times 10^{-31}$ |
| endometrial | Weighted median | 1.89 (1.58 to 2.26) | $5.18 \times 10^{-12}$ |
| cancer | Weighted mode | 1.82 (1.35 to 2.44) | $9.88 \times 10^{-5}$ |
| | MR Egger | 2.03 (1.54 to 2.67) | $8.41 \times 10^{-7}$ |
| Endometrioid | IVW | 1.89 (1.65 to 2.16) | $1.67 \times 10^{-20}$ |
| endometrial | Weighted median | 1.99 (1.60 to 2.46) | $3.24 \times 10^{-10}$ |
| cancer | Weighted mode | 1.96 (1.42 to 2.69) | $4.67 \times 10^{-5}$ |
| | MR Egger | 2.15 (1.52 to 3.03) | $1.74 \times 10^{-5}$ |
| Non-endometrioid | IVW | 1.67 (1.19 to 2.35) | $3.03 \times 10^{-3}$ |
| endometrial | Weighted median | 2.29 (1.24 to 4.22) | $8.24 \times 10^{-3}$ |
| cancer | Weighted mode | 1.89 (0.77 to 4.63) | $1.63 \times 10^{-1}$ |
| | MR Egger | 2.25 (0.99 to 5.07) | $5.28 \times 10^{-2}$ |

ORs are shown per increase in SD (4.7 kg/m<sup>2</sup>) BMI. BMI = body mass index, IVW = inverse-variance weighted.

In subtype-stratified analyses, there was evidence to support an effect of BMI on risk of both endometrioid (OR per SD (4.7 kg/m^2^) increase in BMI: 1.89, 95% CI: 1.65 to 2.16, *P* = 1.67 x 10^-20^) and non-endometrioid endometrial cancer (OR per SD (4.7 kg/m^2^) increase in BMI: 1.67, 95% CI: 1.19 to 2.35, *P* = 3.03 x 10^-3^) (**Fig 4**, **Table 2**). These findings were robust to sensitivity analyses for endometrioid endometrial cancer; however, findings were less consistent for non-endometrioid endometrial cancer in sensitivity analyses using female-specific BMI instruments (**S10-11 Figure**, **S8-9 Table**). Therefore, only overall and endometrioid endometrial cancer were included in follow-up analyses.

### Evaluating the effect of previously reported molecular risk factors on endometrial cancer risk

When examining the effect of previously reported molecular risk factors on overall endometrial cancer risk, there was strong evidence for an effect of total testosterone (OR per increase in inverse-normal transformed (INT) nmol/L total testosterone: 1.64, 95% CI: 1.43 to 1.88, *P* = 1.71 x 10^-12^), bioavailable testosterone (OR per increase in INT nmol/L bioavailable testosterone: 1.46, 95% CI: 1.29 to 1.65, *P* = 3.48 x 10^-9^), fasting insulin (OR per increase in natural log transformed pmol/L fasting insulin 3.93, 95% CI: 2.29 to 6.74, *P* = 7.18 x 10^-7^), and SHBG (OR per increase in INT nmol/L SHBG 0.71, 95% CI: 0.59 to 0.85, *P* = 2.07 x 10^-4^) on endometrial cancer risk (**Fig 5; Table 3**). In addition, there was suggestive evidence for an effect of total serum cholesterol (OR per increase in SD (41.7 mg/dL) total serum cholesterol 0.90, 95% CI: 0.81 to 1.00, *P* = 4.01 x 10^-2^) on overall endometrial cancer risk. These findings were consistent across sensitivity analyses (**S12-S16 Figure, S17 Table, S8-S9 Table**).

**Fig 5.**
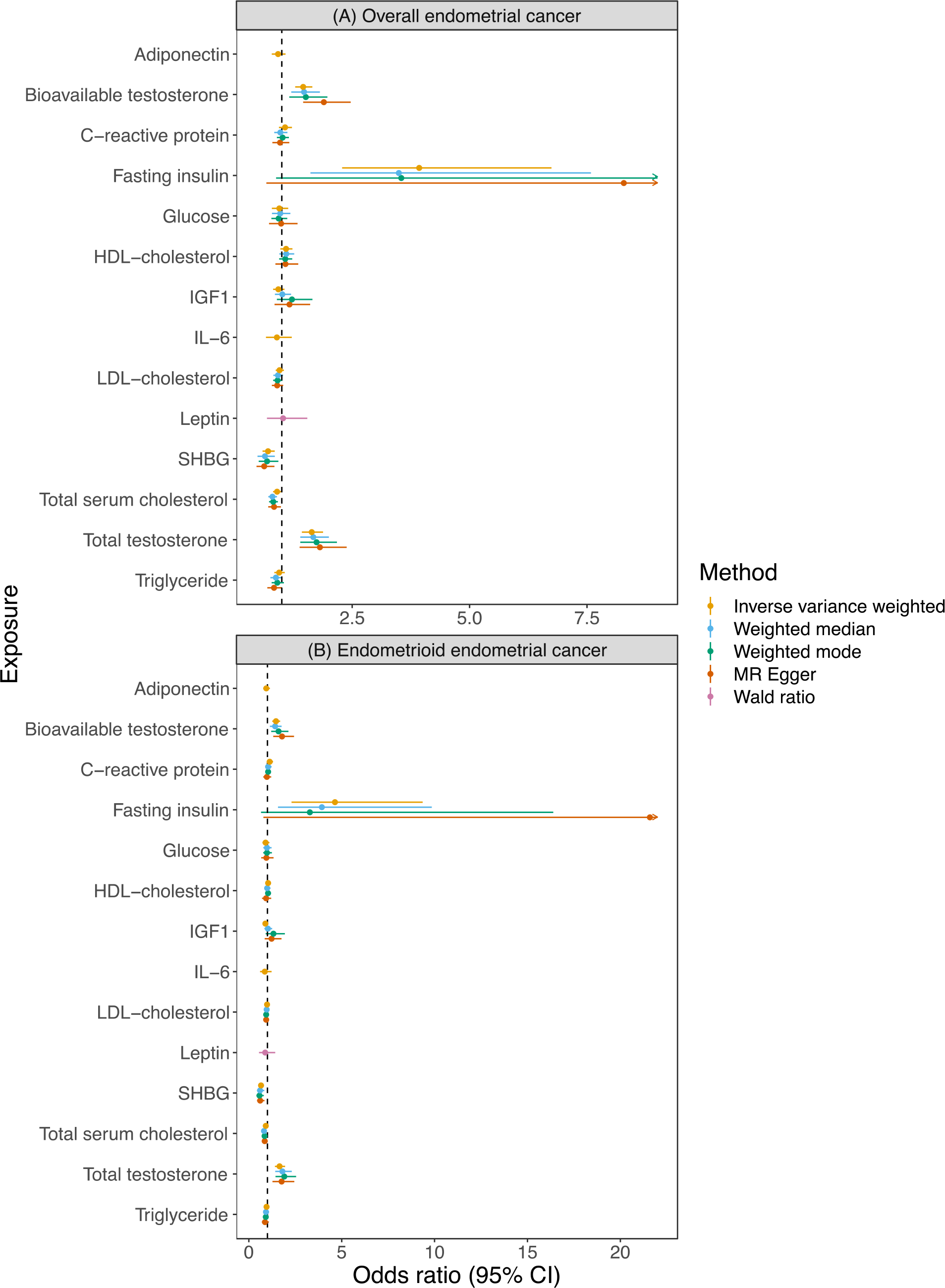
Mendelian randomization analysis of total serum cholesterol, fasting insulin, total testosterone, bioavailable testosterone, and sex hormone-binding globulin (SHBG) on overall and endometrioid endometrial cancer risk. LDL = low-density lipoprotein, HDL = high-density lipoprotein, IGF-1 = insulin-like growth factor-1, IL-6 = interleukin-6, CRP = C-reactive protein, SHBG = sex hormone-binding globulin. (A) Results of MR analyses examining the effects of previously reported molecular risk factors on risk of overall endometrial cancer risk. (B) Results of MR analyses examining the effects of previously reported molecular risk factors on risk of endometrioid subtype endometrial cancer risk.

**Table 3.**
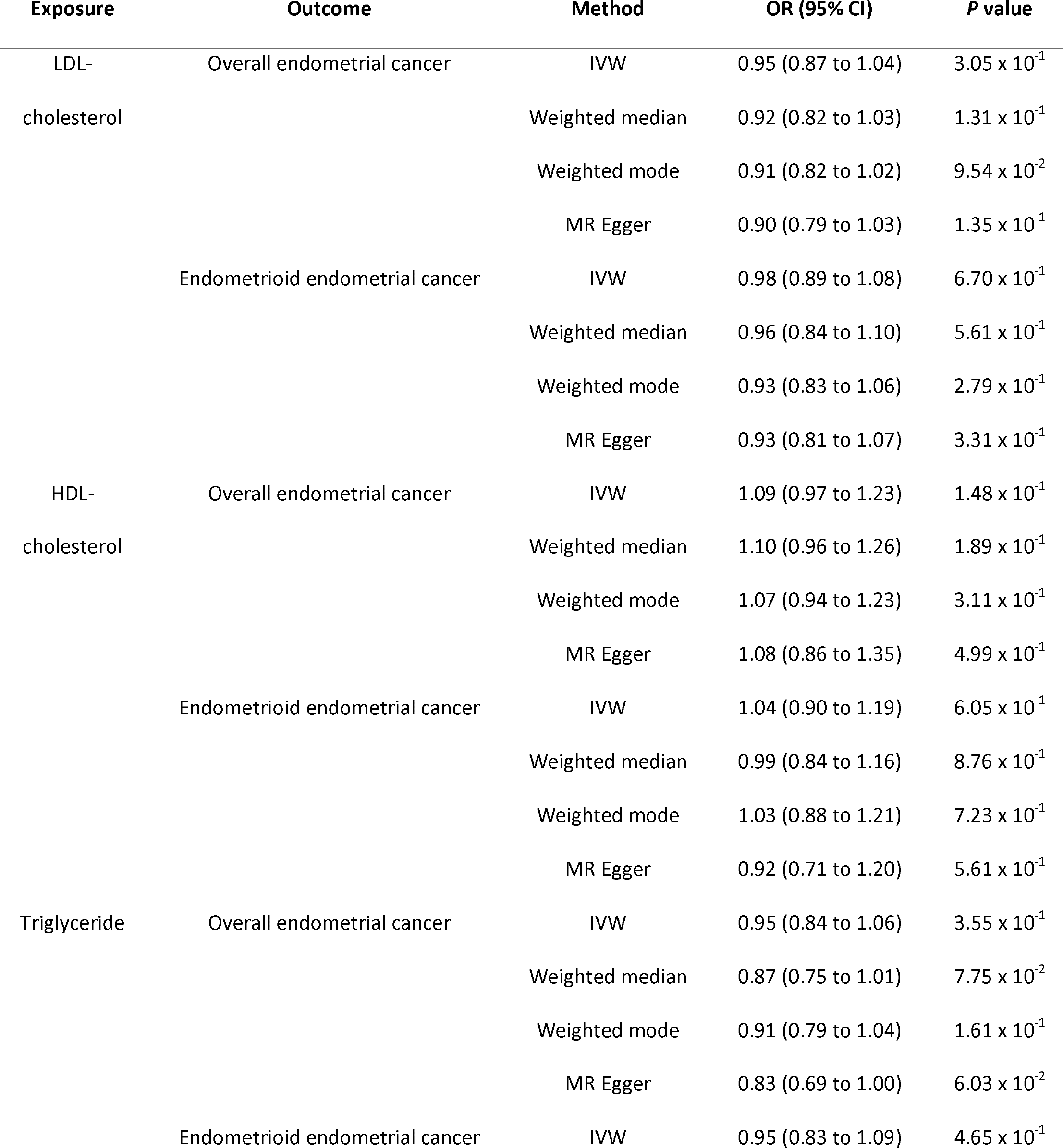
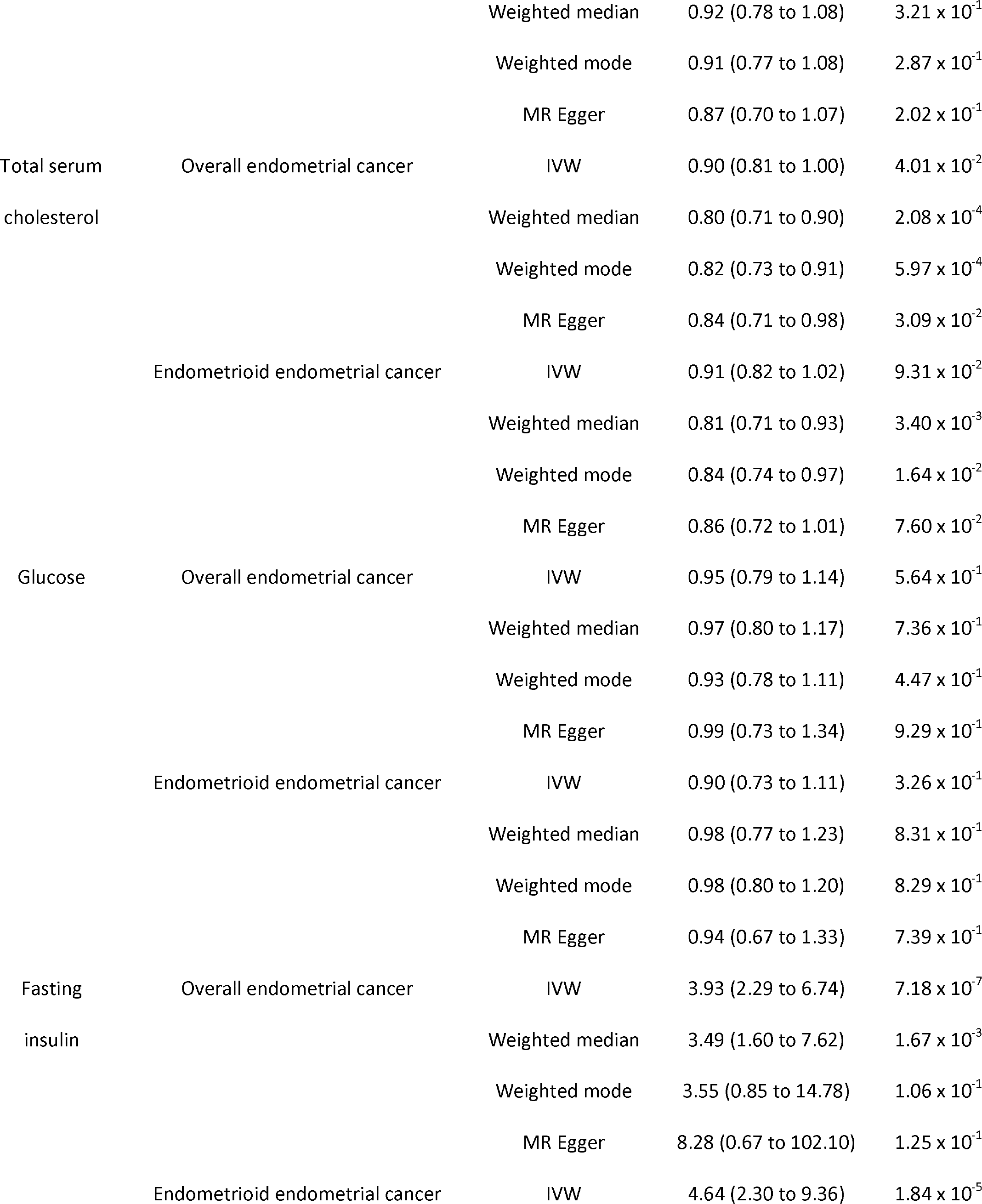
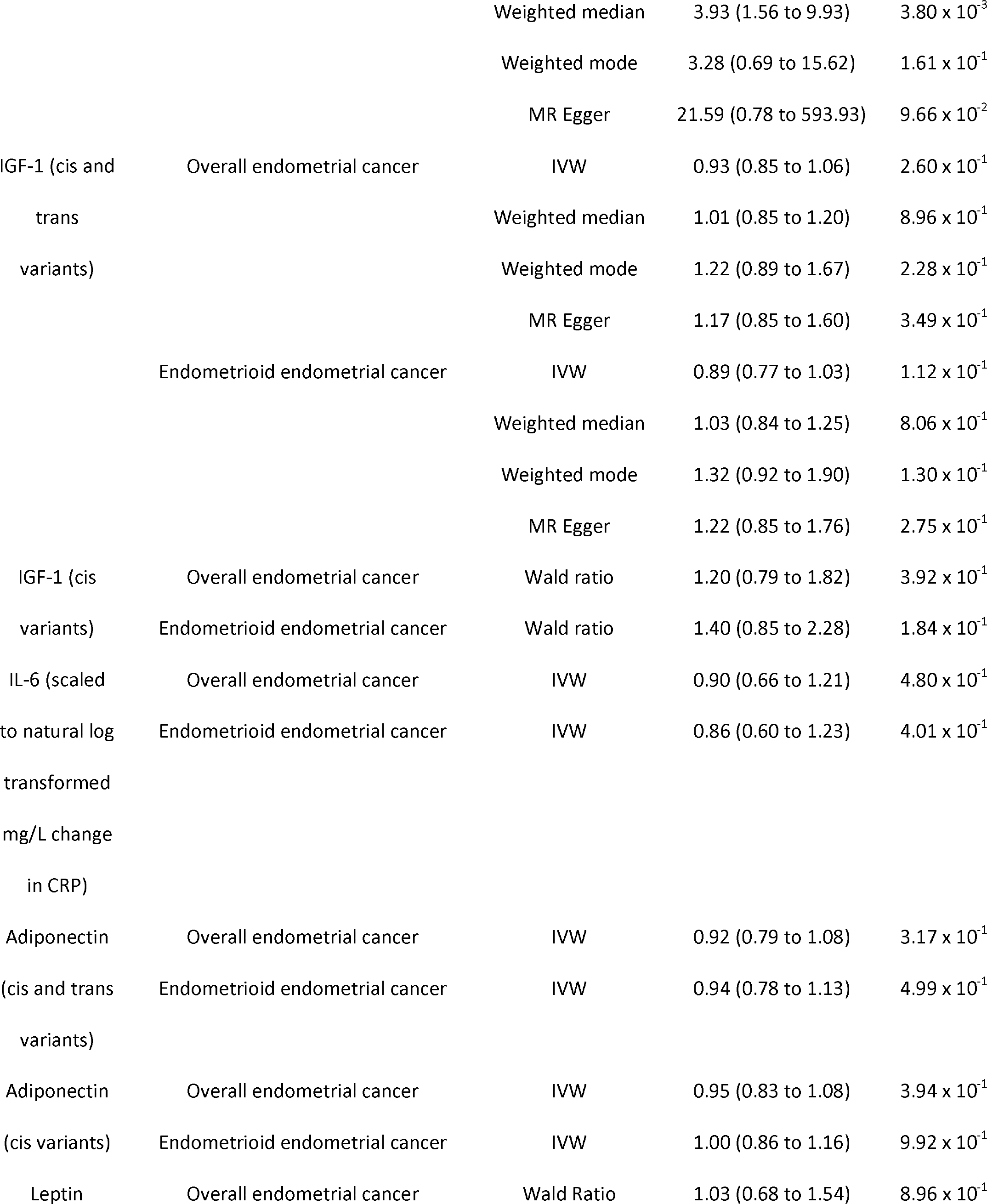
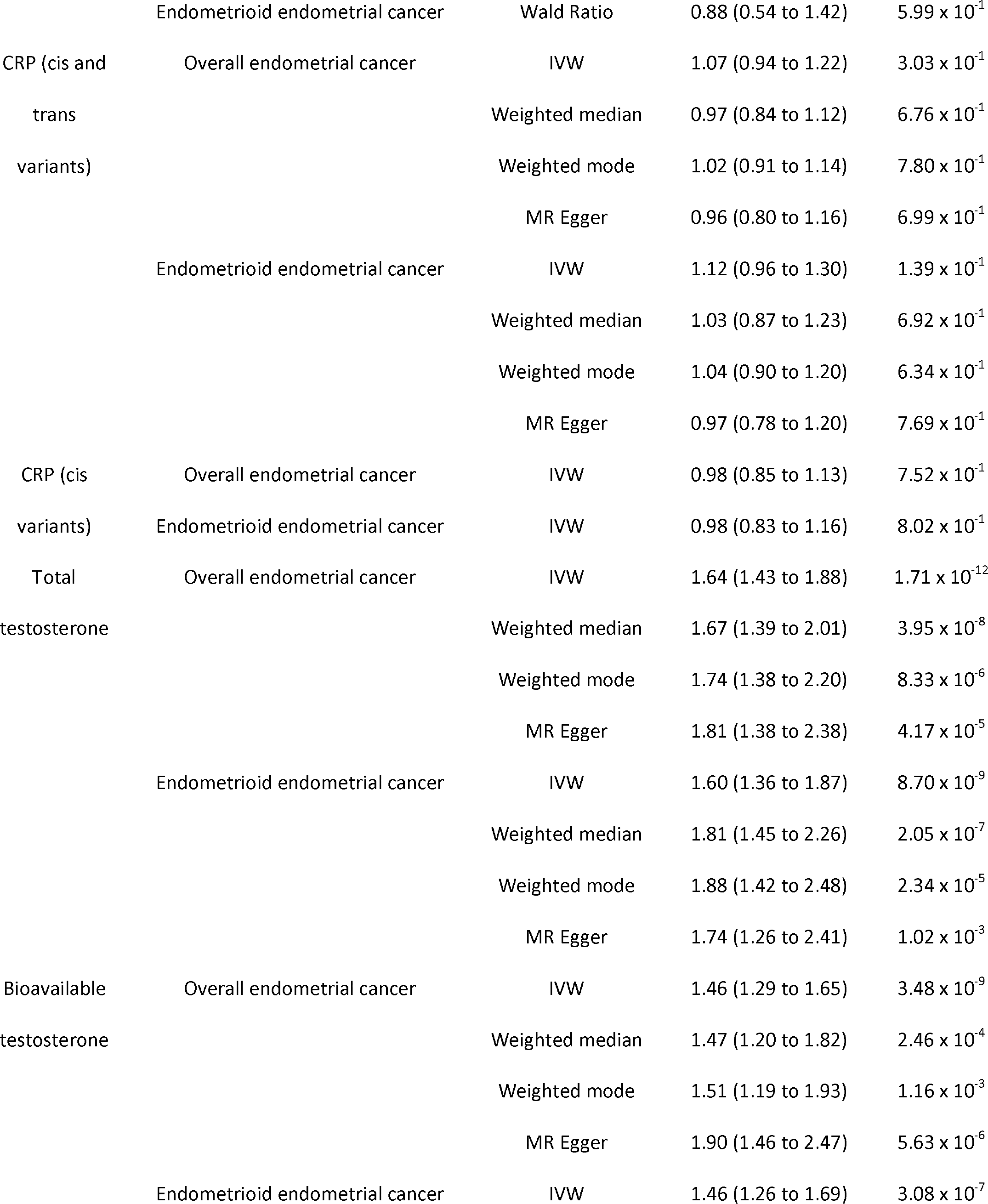
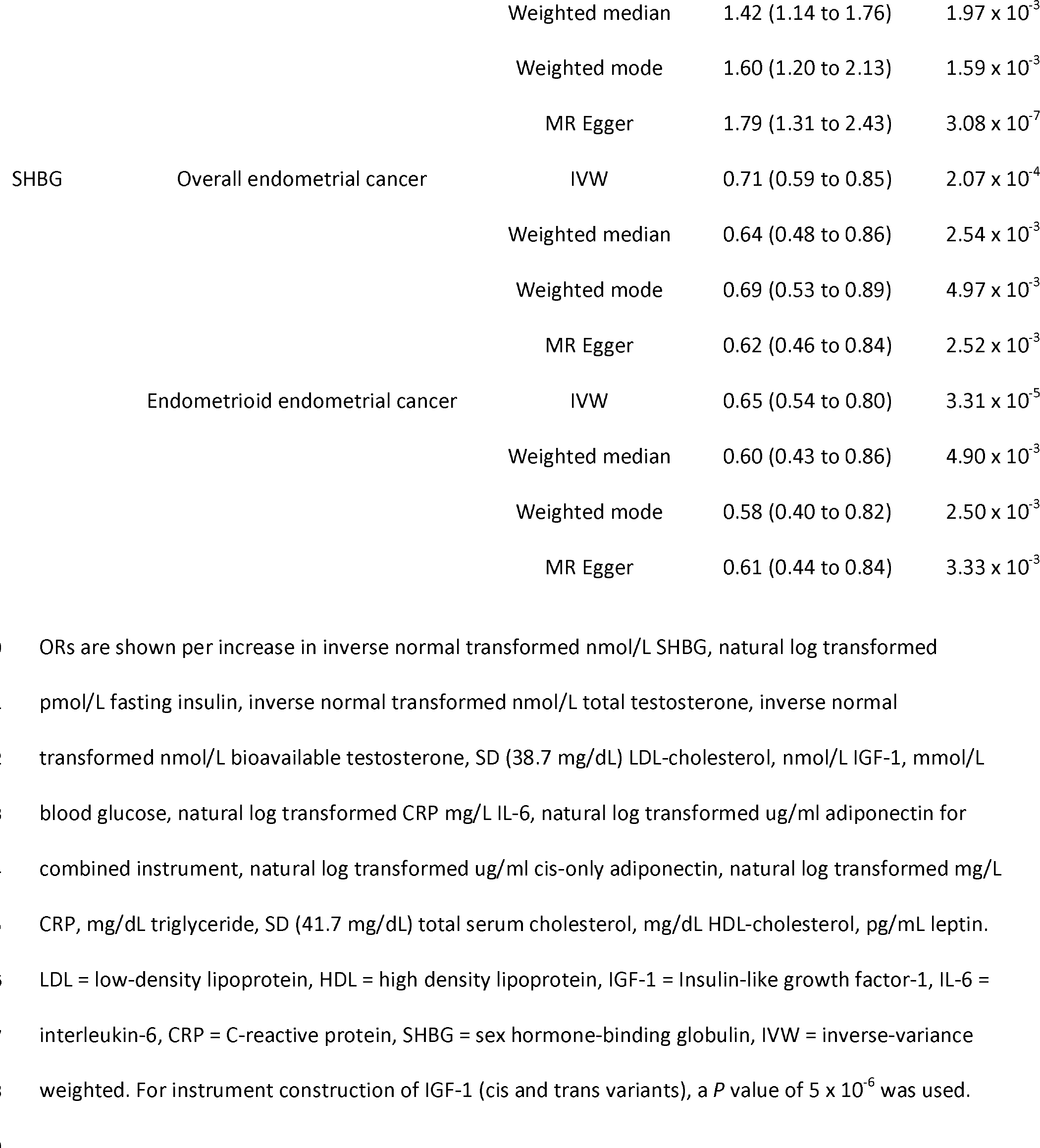
Results of MR analyses examining the effect of previously reported and novel potential molecular risk factors and overall and endometrioid subtype endometrial cancer risk.

In subtype-stratified analyses, there was strong evidence to support an effect of total testosterone (OR per increase in INT nmol/L total testosterone: 1.60, 95% CI: 1.36 to 1.87, *P* = 8.70 x 10^-9^), bioavailable testosterone (OR per increase in INT nmol/L bioavailable testosterone: 1.46, 95% CI: 1.29 to 1.65, *P* = 3.48 x 10^-9^), fasting insulin (OR per increase in natural log transformed pmol/L fasting insulin: 4.64, 95% CI: 2.30 to 9.36, *P* = 1.84 x 10^-5^), and SHBG (OR per increase in INT nmol/L SHBG: 0.65, 95% CI: 0.54 to 0.80, *P* = 3.31 x 10^-5^) on endometrioid endometrial cancer risk (**Fig 5; Table 3**). Findings were consistent across all sensitivity analyses (**S19-S22 Figure, S9 Table**, **S17 Table**).

### Evaluating the effect of BMI on previously reported molecular risk factors

When examining the effect of BMI on molecular traits confirmed to have a causal effect on endometrial cancer (either overall or endometrioid subtype), there was evidence for an effect of BMI on fasting insulin (change in natural log transformed fasting insulin: 0.17, 95% CI: 0.15 to 0.19, *P* = 1.51 x 10^-74^), SHBG (change in INT SHBG: -0.17, 95% CI: -0.19 to -0.16, *P* = 4.86 x 10^-125^), bioavailable testosterone (change in INT bioavailable testosterone: 0.26, 95% CI: 0.23 to 0.29, *P* = 9.97 x 10^-68^), total testosterone (change in INT total testosterone: 0.08, 95% CI: 0.05 to 0.11, *P* = 9.04 x 10^-10^), and CRP (change in ln-transformed CRP: 0.35, 95% CI: 0.32 to 0.38, *P* = 2.67 x 10^-127^) (**Fig 6; Table 4**). The direction of effect in MR analyses examining the effect of BMI on total testosterone was inconsistent when employing a weighted mode model, suggesting the potential presence of horizontal pleiotropy. Although there was little evidence for a causal effect of BMI on total serum cholesterol in the IVW model, there was some evidence for an effect across all three MR sensitivity analysis models, suggesting that horizontal pleiotropy may be biasing the IVW estimate towards the null. All other findings were consistent across the various sensitivity analyses (**S23-S27 Figure, S8 Table, S17-S18 Table**).

**Fig 6.**
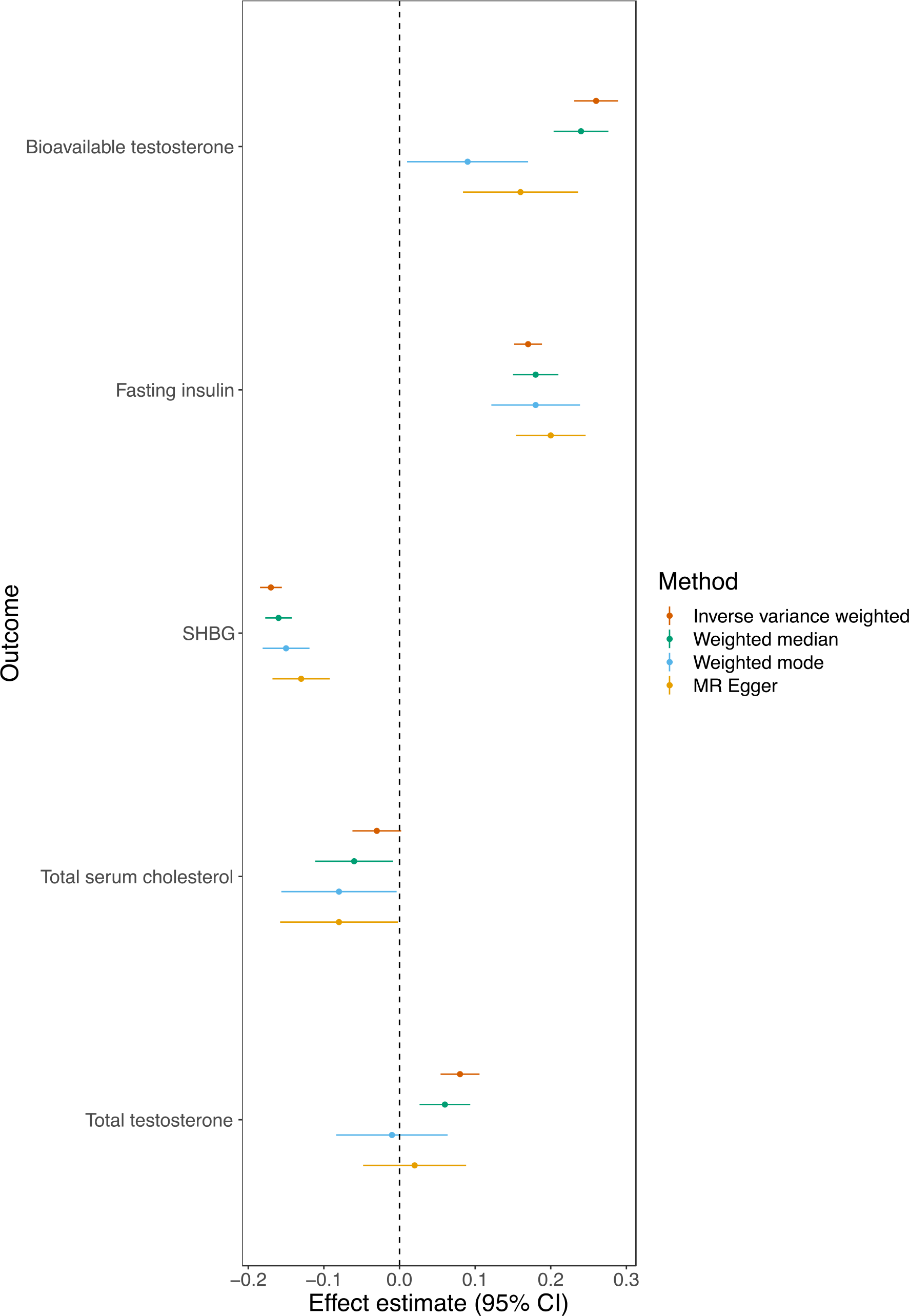
Mendelian randomization analysis of adult BMI on previously reported endometrial cancer risk factors. SHBG = sex hormone-binding globulin, LDL = low-density lipoprotein, CRP = C-reactive protein.

**Table 4.**
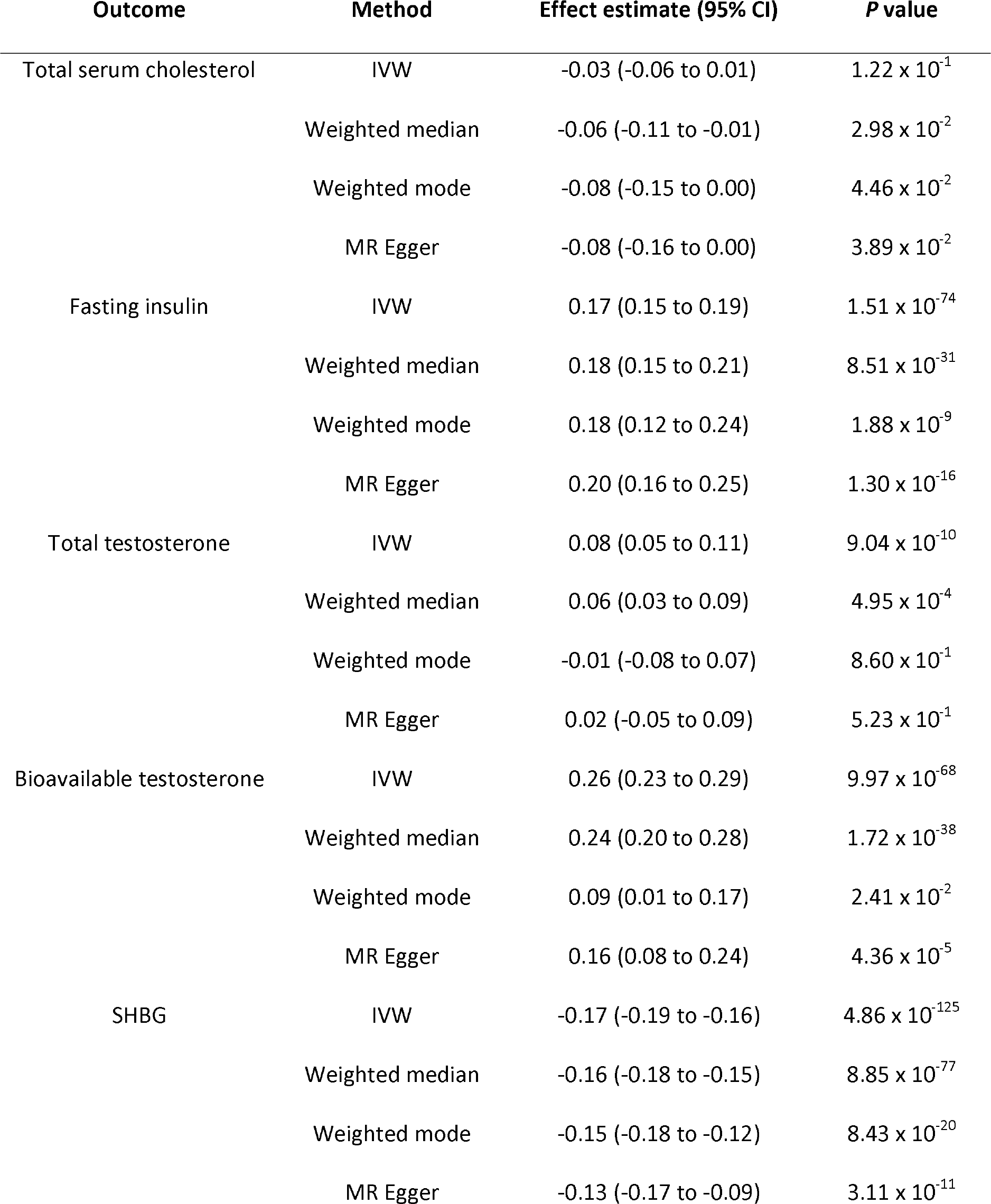
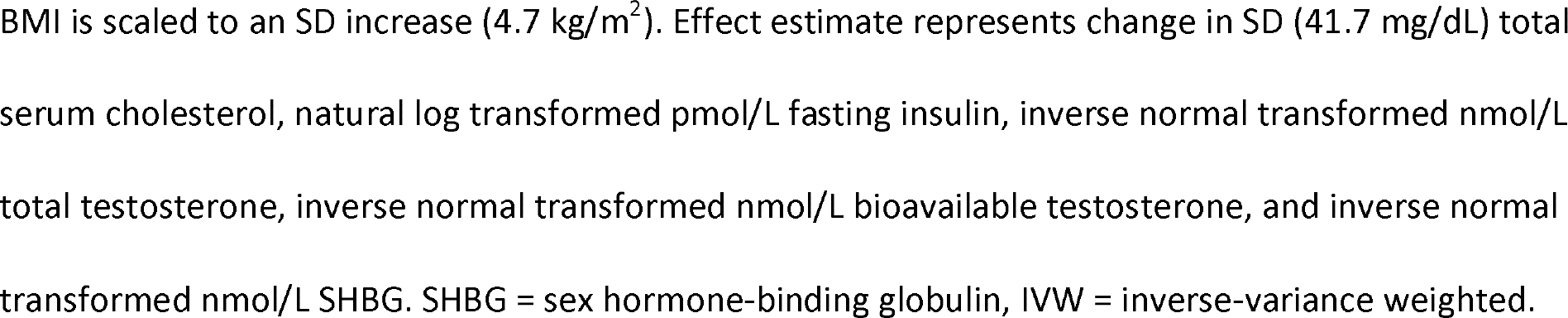
Results of MR analyses examining the effect of BMI on previously reported molecular risk factors.

| Outcome | Method | Effect estimate (95% CI) | P value |
| --- | --- | --- | --- |
| Total serum cholesterol | IVW | -0.03 (-0.06 to 0.01) | $1.22 \times 10^{-1}$ |
| | Weighted median | -0.06 (-0.11 to -0.01) | $2.98 \times 10^{-2}$ |
| | Weighted mode | -0.08 (-0.15 to 0.00) | $4.46 \times 10^{-2}$ |
| | MR Egger | -0.08 (-0.16 to 0.00) | $3.89 \times 10^{-2}$ |
| Fasting insulin | IVW | 0.17 (0.15 to 0.19) | $1.51 \times 10^{-74}$ |
| | Weighted median | 0.18 (0.15 to 0.21) | $8.51 \times 10^{-31}$ |
| | Weighted mode | 0.18 (0.12 to 0.24) | $1.88 \times 10^{-9}$ |
| | MR Egger | 0.20 (0.16 to 0.25) | $1.30 \times 10^{-16}$ |
| Total testosterone | IVW | 0.08 (0.05 to 0.11) | $9.04 \times 10^{-10}$ |
| | Weighted median | 0.06 (0.03 to 0.09) | $4.95 \times 10^{-4}$ |
| | Weighted mode | -0.01 (-0.08 to 0.07) | $8.60 \times 10^{-1}$ |
| | MR Egger | 0.02 (-0.05 to 0.09) | $5.23 \times 10^{-1}$ |
| Bioavailable testosterone | IVW | 0.26 (0.23 to 0.29) | $9.97 \times 10^{-68}$ |
| | Weighted median | 0.24 (0.20 to 0.28) | $1.72 \times 10^{-38}$ |
| | Weighted mode | 0.09 (0.01 to 0.17) | $2.41 \times 10^{-2}$ |
| | MR Egger | 0.16 (0.08 to 0.24) | $4.36 \times 10^{-5}$ |
| SHBG | IVW | -0.17 (-0.19 to -0.16) | $4.86 \times 10^{-125}$ |
| | Weighted median | -0.16 (-0.18 to -0.15) | $8.85 \times 10^{-77}$ |
| | Weighted mode | -0.15 (-0.18 to -0.12) | $8.43 \times 10^{-20}$ |
| | MR Egger | -0.13 (-0.17 to -0.09) | $3.11 \times 10^{-11}$ |

BMI is scaled to an SD increase ( $4.7 \text{ kg/m}^2$ ). Effect estimate represents change in SD (41.7 mg/dL) total serum cholesterol, natural log transformed pmol/L fasting insulin, inverse normal transformed nmol/L total testosterone, inverse normal transformed nmol/L bioavailable testosterone, and inverse normal transformed nmol/L SHBG. SHBG = sex hormone-binding globulin, IVW = inverse-variance weighted.

### Mendelian randomization mediation analysis

In mediation analyses evaluating the potential mediating role of molecular traits previously shown to be on the causal pathway between BMI and endometrial cancer, there was evidence for a mediating role of bioavailable testosterone (15% mediated, 95% CI: 10 to 20%, *P* = 1.43 x 10^-8^), fasting insulin (11% of total effect mediated, 95% CI: 1 to 21%, *P* = 2.89 x 10^-2^), and SHBG (7% mediated, 95% CI: 1 to 12%, *P* = 1.81 x 10^-2^) in the relationship between BMI and overall endometrial cancer risk (**Table 5**). There was also evidence for a mediating role of bioavailable testosterone (15% mediated, 95% CI: 9 to 22%, *P* = 2.15 x 10^-6^) and fasting insulin (16% mediated, 95% CI: 1 to 21%, *P* = 2.89 x 10^-2^) in the relationship between BMI and endometrioid endometrial cancer risk (**Table 5**). Although there was little evidence for a mediating role of SHBG in the relationship between sex-combined BMI and endometrioid endometrial cancer (2% mediated, 95% CI: -9 to 14%, *P* = 6.87 x 10^-1^), in the female-specific BMI sensitivity analysis there was strong evidence for a mediating role of female-specific SHBG in the relationship between BMI and endometrioid endometrial cancer (8% mediated, 95% CI: 3 to 13%, *P* = 3.38 x 10^-3^). Other than this, findings were consistent across sex-specific BMI, fasting insulin and CRP sensitivity analyses (**S28-S29 Table**).

**Table 5.**
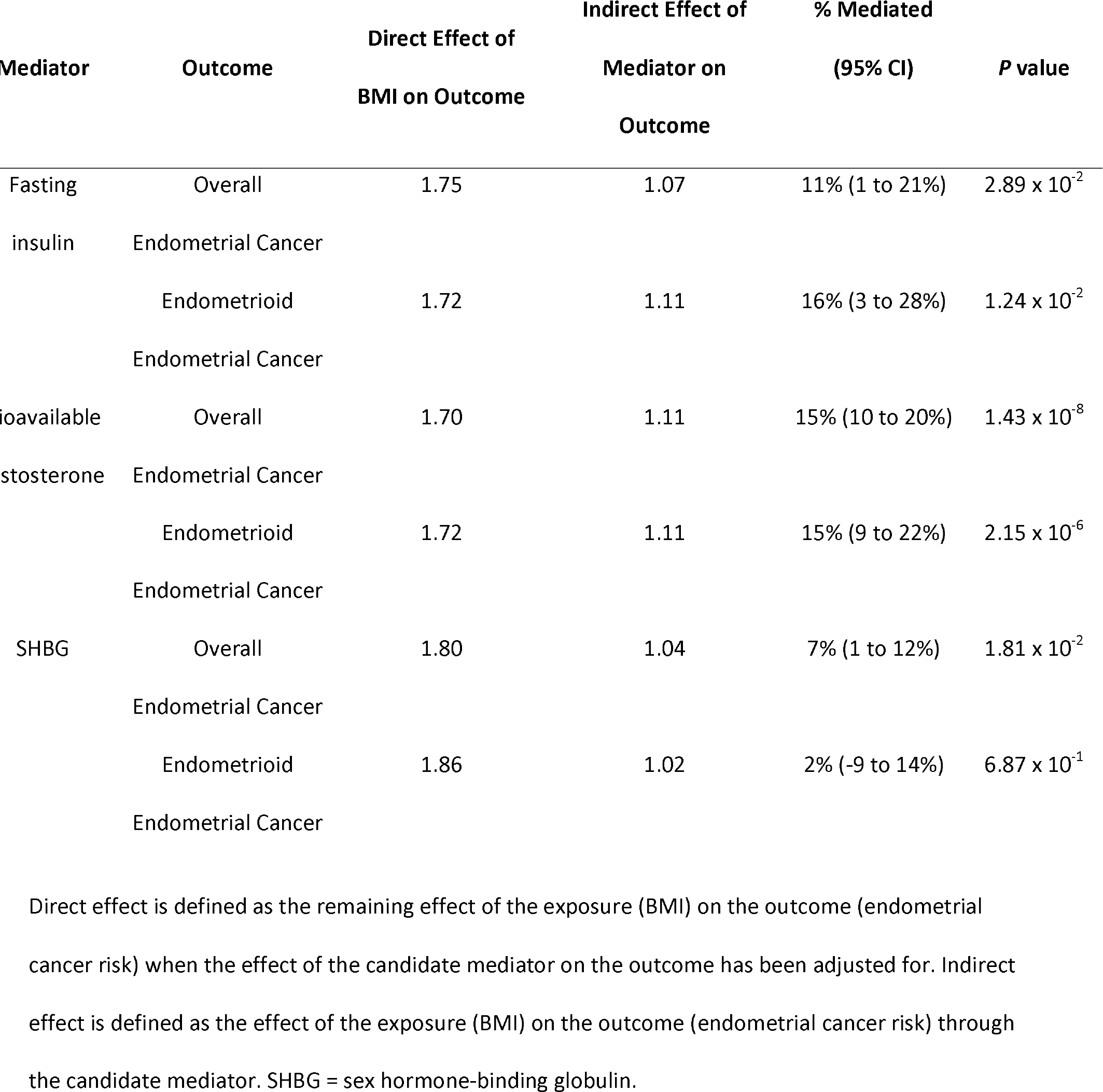
Results of multivariable MR mediation analysis examining the effect of BMI and endometrial cancer with previously reported molecular risk factors as potential mediators.

The conditional F-statistic for both fasting insulin (F=2) and BMI (F=6) in the multivariable MR performed to evaluate the “proportion mediated” by fasting insulin was < 10, indicating that there may be weak instrument bias in these analyses (i.e. over- or underestimation of the “proportion mediated” by fasting insulin) [56]. When re-performing the “proportion mediated” analysis for fasting insulin using an alternative approach (i.e. using an alternative fasting insulin instrument with a larger sample size and limiting the number of SNPs included in the BMI instrument to the 100 with the strongest evidence of association using an LD threshold r^2^<0.001), we found that fasting insulin mediated 19% (95% CI: 5 to 34%, *P* = 9.17 x 10^-3^) of the relationship between BMI and overall endometrial cancer risk and 21% (95% CI: 5 to 38%, *P* = 1.17 x 10^-2^) of the relationship between BMI and endometrioid endometrial cancer risk (**S30 Table**).

In mediation analyses combining pairs of mediators into a single model, the effect of fasting insulin on overall endometrial cancer risk attenuated (∼40% log OR reduction) when SHBG (a presumed downstream mediator of fasting insulin) was included in the model (OR per increase in natural log transformed pmol/L fasting insulin: 2.28, 95% CI: 1.34 to 3.86, *P* = 2.85 x 10^-3^). The effect of SHBG on overall endometrial cancer attenuated fully when bioavailable testosterone (a presumed downstream mediator of SHBG) was included in the model (OR per increase in INT nmol/L SHBG: 1.08, 95% CI: 0.86 to 1.36, P = 5.00 x 10^-1^) (**S34 Table**). The effect of fasting insulin on overall endometrial cancer was strongly attenuated when bioavailable testosterone was included in the model (OR per increase in natural log transformed pmol/L fasting insulin: 1.22, 95% CI: 0.48 to 3.11, P = 6.78 x 10^-1^), which could reflect mediation of the effect of fasting insulin on endometrial cancer via bioavailable testosterone or the presence of conditionally weak instruments in this model. This could result in over- or underestimation of the proportion of the effect of fasting insulin mediated by bioavailable testosterone.

For the endometrioid histological subtype, using the same approach, the effect of fasting insulin on endometrioid endometrial cancer did not markedly change (∼14% logOR reduction) when SHBG was included in the model (OR per increase in natural log transformed pmol/L fasting insulin: 3.74, 95% CI: 0.74 to 19.01, *P* = 1.56 x 10^-1^). However, the effect of SHBG on endometrioid endometrial cancer attenuated fully when bioavailable testosterone was included in the model (OR per increase in INT nmol/L SHBG: 1.16, 95% CI: 0.81 to 1.65, P = 4.12 x 10^-1^). As with analyses of overall endometrial cancer, when fasting insulin and bioavailable testosterone were combined into a single model, the effect of fasting insulin on endometrioid endometrial cancer attenuated toward the null (OR per increase in natural log transformed pmol/L fasting insulin: 1.05, 95% CI: 0.36 to 3.03, P = 9.33 x 10^-1^), potentially reflecting mediation via bioavailable testosterone or persistent weak instrument bias in this model.

## Discussion

Our systematic Mendelian randomization (MR) analysis of 14 previously reported molecular risk factors and body mass index (BMI) in 12,906 endometrial cancer cases and 108,979 controls provided evidence for roles of elevated BMI, fasting insulin, total and bioavailable testosterone, and sex hormone-binding globulin (SHBG) in risk of overall and endometrioid endometrial cancer. In mediation analyses, we found evidence that fasting insulin, bioavailable testosterone concentrations, and SHBG partially mediated the effect of BMI on overall endometrial cancer risk. When combining pairs of mediators together into a single model, we found evidence that an effect of fasting insulin on endometrial cancer was partially mediated by SHBG levels and that an effect of SHBG on endometrial cancer was largely mediated by bioavailable testosterone levels. An effect of fasting insulin on endometrial cancer risk was also strongly attenuated upon adjustment for bioavailable testosterone levels which could reflect mediation of this effect by bioavailable testosterone or conditionally weak instrument bias for fasting insulin concentrations in this analysis. Our analyses found little evidence that several previously reported molecular risk factors, including several metabolic factors (e.g. LDL-C, HDL-C, IGF-1, adiponectin, leptin) and inflammatory markers (CRP, IL-6), were causally implicated in overall or endometrioid endometrial cancer risk.

Several of the findings in this analysis are consistent with evidence from prior conventional observational and MR analyses. For example, the effect of BMI on endometrial cancer risk and the stronger evidence of an effect on endometrioid, as compared to non-endometrioid, endometrial cancer is well-established in the literature and has been shown previously in an MR analysis that used an alternative strategy for instrument construction. Our findings supporting a causal effect of BMI on endometrial cancer risk (OR 1.88, 95% CI: 1.69 to 2.09 per SD (4.7 kg/m^2^) increase) are larger in magnitude than those from pooled analyses of conventional observational analyses (e.g. the World Cancer Research Fund (WCRF) pooled analysis of 26 prospective studies: RR 1.50, 95% CI 1.42 to 1.59 per 5.0 kg/m^2^ increase), consistent with previous comparisons of observational and MR estimates across other cancer sites [57, 58]. Smaller magnitudes of effect in observational analyses may reflect regression dilution bias from single time-point measurements of BMI and/or reverse causation from cancer-induced weight loss, whereas MR estimates reflect accumulated exposure across the life-course and are unlikely to be influenced by reverse causation [59].

In agreement with previous MR analyses, our results suggest a causal role of fasting insulin, total and bioavailable testosterone, and SHBG in endometrial cancer risk, although these previous reports either employed smaller sample sizes than this analysis (e.g. fasting insulin analyses were performed in 1,287 endometrial cancer cases vs 12,906 cases in our analysis) or used somewhat differing methods to examine instrumental variable assumptions [28–30]. The restriction of an effect of BMI to bioavailable (and not total) testosterone is in agreement with previous observational studies which have suggested that BMI influences testosterone levels through decreased production of SHBG rather than a direct effect on testosterone production [60–64]. Additionally, important mediating roles of fasting insulin, bioavailable testosterone, and SHBG in the relationship between BMI and endometrial cancer are consistent with studies of bariatric surgery which have suggested protective effects of this procedure against endometrial cancer risk, along with reductions in insulin and bioavailable testosterone levels, and increases in SHBG levels [65–73]. Our findings supporting a role of BMI on these traits are also consistent with the important endocrine function of adipose tissue, which is involved in sex steroid metabolism [62, 74–79].

Potential aetiological roles of the molecular mediators identified in this analysis are consistent with the “unopposed oestrogen” hypothesis which postulates that endometrial carcinogenesis is driven by excess endogenous or exogenous oestrogen levels that are unopposed by progesterone [80–82]. We were unable to incorporate oestrogen into this analysis as we were unable to identify reliable genetic instruments for this trait. All three of the molecular mediators highlighted in this analysis, however, are known to influence oestrogen: bioavailable testosterone is aromatized to oestradiol; SHBG binds with high-affinity to both oestradiol and bioavailable testosterone [82–87]; and insulin increases androgen and decreases SHBG production [88–91]. We found that an inverse effect of SHBG on endometrial cancer risk was largely attenuated upon adjustment for bioavailable testosterone, suggesting a protective effect of SHBG may be driven via binding of biologically active fractions of circulating testosterone. The attenuation of an effect of fasting insulin on endometrial cancer upon adjustment for bioavailable testosterone could reflect mediation of this effect or the presence of conditionally weak instrument bias in this model. In support of the latter explanation, there is biological evidence that hyperinsulinemia and insulin resistance influence endometrial cancer via oestrogen-independent pathways. For example, insulin has been shown to bind directly to endometrial cells and promote proliferation, and can activate two pathways known to have an important role in carcinogenesis – the phosphatidylinositol-3-kinase-protein kinase B/Akt (PI3K-PKB/Akt) and Ras/Raf/mitogen-activated protein kinase (Ras/Raf/MAPK) pathways [91–96].

Some findings from this MR analysis are not in agreement with evidence from previous conventional observational studies. For example, our analyses found little evidence to support causal roles of several metabolic traits (e.g. circulating HDL-cholesterol, triglycerides, adiponectin, leptin) and inflammatory markers (CRP, IL-6) in endometrial cancer risk, despite these traits being linked to endometrial cancer risk in conventional observational analyses [18–22]. Several of these traits (e.g. HDL-cholesterol, LDL-cholesterol, triglycerides) represent highly correlated metabolic perturbations associated with the obese phenotype which may be too clustered to disentangle using conventional multivariable regression methods [97]. Consequently, some of the divergence in findings across previous conventional observational studies and this MR analysis could reflect residual confounding in the former. Another potential explanation for divergence in findings is the susceptibility of conventional observational studies to reverse causation (i.e., latent, undiagnosed endometrial cancer influencing levels of a presumed exposure). For example, a previously reported association of circulating IL-6 concentrations with endometrial cancer risk could reflect IL-6 secretion by endometrial cancer-associated fibroblasts rather than a role of IL-6 in endometrial cancer development [98, 99].

We were unable to replicate a previously reported MR-based inverse association of LDL cholesterol levels and endometrial cancer risk in the Endometrial Cancer Association Consortium (IVW OR per SD increase in LDL cholesterol: 0.90, 95% CI: 0.85 to 0.95, *P* = 8.39 x 10^-5^). In the previous analysis, SNPs were permitted to be in weak linkage disequilibrium (LD) (pairwise correlation r^2^ < 0.05 vs r^2^ < 0.001 in our analysis) and a Heterogeneity in Dependent Instruments (HEIDI) test was performed to identify potentially pleiotropic SNPs, resulting in the removal of 6 such SNPs from the 146 SNPs initially used as an instrument. We attempted to replicate these previously reported findings using a more stringent r^2^ threshold (i.e. r^2^<0.001) followed by use of the HEIDI test (resulting in the removal of 2 potentially pleiotropic SNPs) which resulted in a causal estimate that was closer in magnitude to that previously reported (IVW OR 0.93, 95% CI: 0.86 to 1.00, *P* = 4.10 x 10^-2^) (**S45 Table**). However, there was greater imprecision in our estimate compared to this previous analysis which could reflect the more liberal LD threshold employed in this earlier analysis.

Our MR analysis provides key insights into potential molecular pathways linking excess adiposity to endometrial cancer risk. This analysis has several strengths including the use of a systematic approach to collate previously reported molecular risk factors for endometrial cancer; the appraisal of their causal relevance in overall and endometrioid endometrial cancer aetiology using an MR framework which should be less prone to conventional issues of confounding and cannot be influenced by reverse causation; the employment of several complementary sensitivity analyses to rigorously assess for violations of MR assumptions; and the use of a summary data-based MR approach which permitted us to leverage large-scale GWAS data from several studies, enhancing statistical power and precision of causal estimates.

There are several limitations to our analysis. First, we were unable to evaluate the role of six previously reported molecular risk factors for endometrial cancer due to the absence of reliable genetic instruments for these traits. These six risk factors included oestradiol which is believed to be an important molecular mediator of the effect of BMI on endometrial cancer risk[9]. Second, some of the effect estimates for SNPs included in genetic instruments were obtained from discovery GWAS and have not been replicated in an independent sample which can result in “Winner’s curse” bias. There was sample overlap in this analysis across certain traits; however, the use of conventionally strong (*P* < 5.0 x 10^-8^) instruments for these traits and the consistency of most findings in sensitivity analyses examining their robustness to potential Winner’s curse bias suggests that this phenomenon was unlikely to markedly influence the results presented in this analysis. Third, although sex-specific sensitivity analyses were performed where data were available, some prior GWAS used in this analysis did not examine for heterogeneity of SNP effects by sex which prevented evaluation of the effect of certain traits on endometrial cancer risk using sex-specific instruments. Fourth, univariable and multivariable MR analyses presented here assume that relationships between exposures and outcomes are linear, although it has been previously suggested that the relationship between BMI and endometrial cancer may best be explained by a non-linear model [12, 100]. Fifth, our analysis was almost exclusively restricted to individuals of European ancestry to minimise bias from population stratification, which may limit the generalisability of our findings to non-European populations. Finally, while various sensitivity analyses were performed to examine violations of exchangeability and exclusion restriction criteria, these assumptions are unverifiable.

With the global incidence of overweight and obesity projected to increase and challenges in implementing successful weight loss strategies, pharmacological approaches targeting molecular mediators of the effect of obesity on endometrial cancer development may offer a viable approach for cancer prevention in high-risk groups [101–105]. Metformin, a safe and inexpensive first-line treatment for type 2 diabetes, could serve as a promising chemoprevention agent for endometrial cancer as it increases insulin sensitivity, thus reversing insulin resistance and lowering fasting insulin levels, alongside inhibiting endometrial proliferation [9, 106]. In addition, unlike some other oral hypoglycaemic medications, metformin users show a tendency toward sustained weight loss [107] . Bioavailable testosterone and SHBG also present potential pharmacological targets, though the multifaceted function of these hormones means that targeting these traits may result in adverse effects [108–113]. Phase II clinical trials examining the efficacy of a combination of contraceptive intrauterine devices, metformin, and weight loss interventions as a non-invasive treatment option for individuals with obesity with early-stage endometrial cancer have had encouraging results, and weight loss has been shown to improve oncological outcomes in women with endometrial cancer undergoing progestin treatment [114, 115].

Our systematic evaluation of 14 previously reported candidate mediators of the effect of BMI on endometrial cancer risk identifies fasting insulin, bioavailable testosterone, and SHBG as plausible mediators of this relationship. While we were unable to entirely disentangle the independent effects of these three traits, identification of a potential mediating role of these traits (and, in particular, fasting insulin) in endometrial carcinogenesis is nonetheless informative for the development of pharmacological interventions targeting these traits for cancer prevention. In this respect, future assessment of the effect of drugs which target molecular mediators identified in this analysis using a “drug-target Mendelian randomization” approach could inform on the potential efficacy of the repurposing of medications for endometrial cancer prevention.

## Conclusion

Our comprehensive Mendelian randomization analysis provides insight into potential causal mechanisms linking excess adiposity to endometrial cancer risk. We show that lifelong cumulative elevated BMI causes a larger increased risk than that reported in previous conventional observational studies. We found strong evidence for a mediating role of fasting insulin, bioavailable testosterone and SHBG in the effect of BMI on endometrial cancer risk. These results suggest pharmacological targeting of insulin-related and hormonal traits as a potential strategy for the prevention of endometrial cancer.

## Data Availability

This study involves only openly available human data, which can be obtained from: the GWAS catalogue (Adult BMI {Yengo et al}; LDL-C, HDL-C, triglycerides, total serum cholesterol {Willer et al}; leptin {Folkersen et al}, endometrial cancer {O'Mara et al)) and the IEU Open GWAS catalogue (Glucose {Neale et al}). Data on fasting insulin (Lagou et al) were obtained from the MAGIC consortium (https://magicinvestigators.org/downloads/). Data on IGF-1 (Sinnott-Armstrong et al) are available here: https://doi.org/10.35092/yhjc.12355382. Data on IL-6 were obtained from results presented in Georgakis et al. Circulation: Genomic and Precision Medicine. 2020;13:e002872 (https://www.ahajournals.org/doi/10.1161/CIRCGEN.119.002872). Data on adiponectin were obtained from Dastani et al. PLoS Genet 8(3): e1002607. (https://journals.plos.org/plosgenetics/article?id=10.1371/journal.pgen.1002607). Data on CRP were obtained from Ligthart et al. Am J Hum Genet. 2018 Nov 1;103(5):691-706 (https://www.cell.com/ajhg/fulltext/S0002-9297(18)30320-3). Data on testosterone (bioavailable and total) and SHBG were obtained by contacting Kate Ruth (k.s.ruth at exeter.ac.uk).

## Acknowledgements

The endometrial cancer genome-wide association analyses were supported by the National Health and Medical Research Council of Australia (APP552402, APP1031333, APP1109286, APP1111246 and APP1061779), the U.S. National Institutes of Health (R01-CA134958), European Research Council (EU FP7 Grant), Wellcome Trust Centre for Human Genetics (090532/Z/09Z) and Cancer Research UK. OncoArray genotyping of ECAC cases was performed with the generous assistance of the Ovarian Cancer Association Consortium (OCAC), which was funded through grants from the U.S. National Institutes of Health (CA1X01HG007491-01 (C.I. Amos), U19-CA148112 (T.A. Sellers), R01-CA149429 (C.M. Phelan) and R01-CA058598 (M.T. Goodman); Canadian Institutes of Health Research (MOP-86727 (L.E. Kelemen)) and the Ovarian Cancer Research Fund (A. Berchuck). We particularly thank the efforts of Cathy Phelan. OncoArray genotyping of the BCAC controls was funded by Genome Canada Grant GPH-129344, NIH Grant U19 CA148065, and Cancer UK Grant C1287/A16563. All studies and funders are listed in O’Mara et al (2018). The authors would like to thank the participants of the 1958 British birth cohort (BS58), Academic Medical Centre Amsterdam Premature Arosclerosis Study (AMC-PAS), Age, Gene/Environment Susceptibility (AGES) study, Anglo-Scandinavian Cardiac Outcome Trial (ASCOT), Aro-Express Biobank Study (AE), Arosclerosis Risk in Communities (ARIC) study, Arosclerotic Disease, Vascular Function, and Genetic Epidemiology (ADVANCE) study, Asian Indian Diabetic Heart Study (AIDHS) /Sikh Diabetes Study (SDS), Avon Longitudinal Study of Parents and Children (ALSPAC), Baltimore Longitudinal Study of Aging (BLSA), British Genetics of Hypertension (BRIGHT) Study, Busselton Health Study (BSN), C Reactive Protein Coronary Heart Disease Genetics Collaboration (CCGC), Cancer Prostate in Sweden 1 (CAPS1) study, Cancer Prostate in Sweden 2 (CAPS2) study, CardioGenics study, Cardiovascular Health Study (CHS), Cardiovascular Health Study (CHS), Cardiovascular Risk in Young Finns Study (YFS), Carotid Intima Media Thickness (IMT) and IMT-Progression as Predictors of Vascular Events in a High Risk European Population (IMPROVE) study, Cebu Longitudinal Health and Nutrition Survey (CLHNS), Charles Bronfman Institute for Personalized Medicine (IPM) BioMe Biobank Program, Cohorte Lausannoise (CoLaus) study, Cohorts for Heart and Aging Research in Genomic Epidemiology (CHARGE) Inflammation Working Group (CIWG), CROATIA-Vis study, Croatian Study (CROAS), Danish National Birth Cohort (DNBC), Data from an Epidemiological Study on Insulin Resistance syndrome (DESIR), Diabetes Epidemiology: Collaborative analysis of Diagnostic criteria in Europe (DECODE) study, Diabetes Genetics Initiative (DGI), DiaGen consortium, Dose-Responses to Exercise Training (DR’s EXTRA) study, Dundee study, Dutch and Belgian Lung Cancer Screening Trial (NELSON), Echinococcus Multilocularis and Internal Diseases in Leutkirch (EMIL) study, Edinburgh Artery Study (EAS), Efficiency and safety of varying frequency of whole blood donation (INTERVAL) study, Ely Study, Endometrial Cancer Association Consortium (ECAC), Epidemiology of Endometrial Cancer Consortium (E2C2), EpiHealth cohort, Erasmus Rucphen Family (ERF) study, Estonian Biobank, Estonian Genome Center of University of Tartu (EGCUT) study, European Prospective Investigation into Cancer and Nutrition (EPIC) study, Family Blood Pressure Program (FBPP), Family Blood Pressure Program (FBPP), Family Heart Study (FamHS), Fast Revascularisation during Instability in Coronary artery disease (FRISCII) study, Fendland study, Finland-United States Investigation of NIDDM Genetics (FUSION) study, FinMetSeq, Finnish Cardiovascular Study (FINCAVAS), Finnish Diabetes Prevention Study (DPS), Finnish Genetic Study of Arrhythmic Events (FinGesture), Finnish National Diabetes Prevention Program (FIN-D2D), Finnish Twin Cohort (FTC), Finrisk study, Framingham Heart Study (FRAM), Genetic Investigation of ANthropometric Traits (GIANT), Gene-Lifestyle interactions And Complex traits Involved in Elevated disease Risk Study (GLACIER), Genetic determinants of Obesity and Metabolic syndrome (DILGOM) study, Genetic Epidemiology Network of Arteriopathy (GENOA) study, Genetic Predisposition of Coronary Heart Disease in Patients Verified with Coronary Angiogram (COROGENE), Genetics of Diabetes Audit and Research Study (GoDartS), GenomEUtwin, German Myocard Infarct Family Study I (GerMiFSI), German Myocard Infarct Family Study II (GerMiFSII), Global Lipids Consortium (GLGC), Gonburg Osteoporosis and Obesity Determinants Study (GOOD), Health 2000 study, Health and Retirement Study (HRS), Health, Aging, and Body Composition Study (Health ABC), Health, Risk Factors, Training and Genetics (HERITAGE) Family Study, Heinz Nixdorf Recall (HNR), Hellenic study of Interactions between Single nucleotide polymorphisms and Eating in Arosclerosis Susceptibility (THISEAS), Helsinki Birth Cohort Study (HBCS), Heredity and Phenotype Intervention (HAPI) Heart Study, Invecchiare in Chianti (InCHIANTI) study, Kingston Gene-by-environment; subset of International Collaborative Study of Hypertension in Blacks (GXE), Kooperative Gesundheitsforschung in der Region Augsburg (KORA) study, Korcula study, Leiden Longevity Study (LLS), Leipzig adults study, Leipzig kids study, Lifelines cohort, London Life Sciences Prospective Population Study (LOLIPOP), LUdwigshafen RIsk and Cardiovascular Health (LURIC) study, MAGIC (the Meta-Analyses of Glucose and Insulin-related traits Consortium), Malmo Preventive Project-Re-examination (MPP-RES) study, Malmo Diet and Cancer Study (MDC), Medical research Council (MRC) National Survey of Health & Development (MRC NSHD), Medical Research Council (MRC) National Survey of Health and Development (NSHD), Medical Research Council/Uganda Virus Research Institute General Population Cohort (MRC/UVRIGPC), Metabolic Syndrome in Men (METSIM) study, Microisolates in South Tyrol Study (MICROS), Molecular Genetics of Schizophrenia (MGS), MOnica Risk, Genetics, Archiving and Monograph (MORGAM), Multiethnic Cohort Study (MEC), Myocardial Infarction Genetics Consortium (Migen), Nerlands Study of Depression and Anxiety (NESDA), Nerlands Twin Register (NTR), Nijmegen Bladder Cancer Study and Nijmegen Biomedical Study (RUNMC), Nord- Trøndelag Health Study (HUNT), Northern Finland Birth Cohort 1966 (NFBC66), Northern Finland Birth Cohort 1986 (NFBC86), Northern Sweden Population Health Study (NSPHS), Nurse’s Health Study (NHS), Orkney Complex Disease Study (ORCADES), Pakistan Risk of Myocardial Infarction Study (PROMIS), Precocious Coronary Artery Disease (PROCARDIS) study, Prevention of REnal and Vascular ENd-stage Disease (PREVEND) study, Prospective Investigation of Vasculature in Uppsala Seniors (PIVUS) study, PROspective study of Pravastatin in Elderly at Risk for vascular disease (PROSPER/PHASE), Prostate, Lung, Colorectal and Ovarian Cancer Screening Trial (PLCO), Quebec Family Study (QFS), Relationship between Insulin Sensitivity and Cardiovascular disease Study (RISC), Rotterdam study, SardiNIA Project, Seychelles Family Study (TANDEM), Sorbs study, Spanish Town study (SPT), STabilization of Arosclerotic plaque By Initiation of darapLadIb rapY (STABILITY), Stanley cohort, Stockholm Coronary Arosclerosis Risk Factor (SCARF) study, Stockholm Heart Epidemiology Program (SHEEP), Studies of Epidemiology and Risk factors in Cancer Heredity / UK Ovarian Cancer Population Study (SEARCH/UKOPS), Study of Health in Pomerania (SHIP), Suivi Temporaire Annuel Non Invasif de la Santé des Lorrains Assurés Sociaux (STANISLAS) cohort, Swedish And Singapore Breast Association Consortium (SASBAC), Swedish Twin Registry (STR), Taichi Consortium, Tracking Adolescents’ Individual Lives Survey (TRAILS), Tromsø Study, Twin study at Queensland Institute of Medical Research (QIMR), Twins UK, UK Biobank, Uppsala Longitudinal Study of Adult Men (ULSAM), Vitamin D and Type 2 Diabetes (D2d) study, Wellcome Trust Case Control Consortium (WTCCC), Whitehall study, Women’s Genome Health Study (WGHS), and Women’s Health Initiative (WHI) for their participation in these studies along with the principal investigators for generating the data utilised for this analysis and for making these data available in the public domain. Data on glycaemic traits have been contributed by MAGIC investigators and have been downloaded from www.magicinvestigators.org, and data on oestradiol and blood glucose have been contributed by the Neale lab and have been downloaded from www.nealelab.is.

## Supporting Information Captions

**S1 Appendix. List of unique molecular traits identified from literature search.**

**S2 Appendix. Supplementary methods.**

**S3 Appendix. PubMed search terms for literature review.**

**S4 Table. Additional information on GWAS, including covariates adjusted for.**

**S5 Table. Sample overlap between GWAS.**

**S6 Table. Conditional F-statistics for multivariable Mendelian randomization of BMI and mediators on endometrial cancer risk.**

**S7 Figure. Leave-one-out analysis for MR examining the effect of adult BMI on overall endometrial cancer risk.**

**S8 Table. Results of female BMI sensitivity analysis MR.**

**S9 Table. Results from sensitivity analyses examining the influence of Winner’s curse on GWAS with overlapping samples.**

**S10 Figure. Leave-one-out analysis for MR examining the effect of adult BMI on endometrioid endometrial cancer risk.**

**S11 Figure. Leave-one-out analysis for MR examining the effect of adult BMI on non-endometrioid endometrial cancer risk.**

**S12 Figure. Leave-one-out analysis for MR examining the effect of total testosterone level on endometrial cancer risk.**

**S13 Figure. Leave-one-out analysis for MR examining the effect of bioavailable testosterone level on endometrial cancer risk.**

**S14 Figure. Leave-one-out analysis for MR examining the effect of fasting insulin level on endometrial cancer risk.**

**S15 Figure. Leave-one-out analysis for MR examining the effect of SHBG level on endometrial cancer risk.**

**S16 Figure. Leave-one-out analysis for MR examining the effect of total serum cholesterol level on endometrial cancer risk.**

**S17 Table. Results of female-specific SNP fasting insulin sensitivity analysis MR.**

**S18 Table. Results of female-specific SNP CRP sensitivity analysis MR.**

**S19 Figure. Leave-one-out analysis for MR examining the effect of total testosterone level on endometrioid endometrial cancer risk.**

**S20 Figure. Leave-one-out analysis for MR examining the effect of bioavailable testosterone level on endometrioid endometrial cancer risk.**

**S21 Figure. Leave-one-out analysis for MR examining the effect of fasting insulin level on endometrioid endometrial cancer risk.**

**S22 Figure. Leave-one-out analysis for MR examining the effect of SHBG level on endometrioid endometrial cancer risk.**

**S23 Figure. Leave-one-out analysis for MR examining the effect of adult BMI on fasting insulin level.**

**S24 Figure. Leave-one-out analysis for MR examining the effect of adult BMI on SHBG level.**

**S25 Figure. Leave-one-out analysis for MR examining the effect of adult BMI on bioavailable testosterone level.**

**S26 Figure. Leave-one-out analysis for MR examining the effect of adult BMI on total serum cholesterol level.**

**S27 Figure. Leave-one-out analysis for MR examining the effect of adult BMI on C-reactive protein level.**

**S28 Table. Results of female-specific SNP BMI sensitivity mediation analysis.**

**S29 Table. Results of female-specific SNP fasting insulin mediation analysis.**

**S30 Table. Results of multivariable MR mediation analysis examining the effect of BMI and endometrial cancer with fasting insulin as a potential mediator with BMI-adjusted fasting insulin instrument and 100 SNPs from BMI instrument.**

**S31 Table. Results from sensitivity analyses examining the influence of Winner’s curse on GWAS with overlapping samples in analyses determining the mediating role of traits in the relationship between BMI and endometrial cancer risk.**

**S32 Table. Results from sensitivity analyses examining the influence of Winner’s curse on GWAS with overlapping samples in analyses determining the interdependent effects of mediators of the relationship between BMI and endometrial cancer risk.**

**S33 Table. Conditional F-statistics for further multivariable Mendelian randomization analyses.**

**S34 Table. Results of multivariable MR mediation analysis examining the effect of endometrial cancer with pairs of confirmed mediating molecular traits without BMI.**

**S35 Table. Results of multivariable MR mediation analysis examining the effect of BMI and endometrial cancer with fasting insulin as a potential mediator with 100 SNPs from BMI instrument.**

**S36 Table. Results of multivariable MR mediation analysis examining the effect of BMI and endometrial cancer with fasting insulin as a potential mediator with BMI-adjusted fasting insulin instrument.**

**S37 Table. Conditional F-statistics for fasting insulin (adjusted for BMI) and BMI with differing thresholds used to construct fasting insulin.**

**S38 Table. Results of multivariable MR mediation analysis examining the effect of BMI and endometrial cancer with all confirmed mediating molecular traits with BMI-adjusted fasting insulin.**

**S39 Table. Results of multivariable MR mediation analysis examining the effect of BMI and endometrial cancer with pairs of confirmed mediating molecular traits.**

**S40 Table. Results of multivariable MR mediation analysis examining the effect of BMI and endometrial cancer with all confirmed mediating molecular traits including fasting insulin adjusted for BMI with 100 SNPs from BMI instrument.**

**S41 Table. Results of multivariable MR mediation analysis examining the effect of BMI and endometrial cancer with pairs of confirmed mediating molecular traits with 100 SNPs from BMI instrument.**

**S42 Table. Results of multivariable MR mediation analysis examining the effect of endometrial cancer with all confirmed mediating molecular traits without BMI.**

**S43 Table. Results of sensitivity analysis examining the effect of only including 100 SNPs for the BMI instrument in MR analyses.**

**S44 Table. Conditional F-statistics with different levels of genetic correlation for SHBG and BMI in multivariable Mendelian randomization analyses.**

**S45 Table. Results of HEIDI test-filtered low-density lipoprotein (LDL) cholesterol and overall endometrial cancer MR.**

