## Supporting Information for "Identifying molecular mediators of the relationship between body mass index and endometrial cancer risk: a Mendelian randomization analysis"

### S1 Appendix. List of unique molecular traits identified from literature search.

1. Oestradiol
2. Low-density lipoprotein (LDL) cholesterol
3. High-density lipoprotein (HDL) cholesterol
4. Fasting insulin
5. Total testosterone
6. Bioavailable testosterone
7. Sex hormone-binding globulin (SHBG)
8. Insulin-like growth factor 1 (IGF-1)
9. Interleukin-6
10. Tumour necrosis factor-a
11. Adiponectin
12. Leptin
13. Insulin growth factor binding protein (IGFBP)-1
14. Blood glucose
15. Triglyceride
16. Total serum cholesterol
17. Visfatin
18. Postchallenge insulin
19. C-reactive protein
20. Progesterone

### S2 Appendix. Supplementary methods.

*Further GWAS information*

The Illumina arrays used for genotyping participants in the GWAS of endometrial cases and controls from which summary genetic data was obtained were the OncoArray chip, Illumina Human OmniExpress array, the Illumina Human 660W array, an Affymetrix Axiom® array, the Illumina Human Omni1-Quad array, the Illumina Infinium OmniExpressExome-8 array, the Axiom Genome-Wide Human CEU array, or the Illumina Infinium iSelect array. Genotypes were imputed using the 1000 Genomes Project v3 reference panel (combined with the UK10K reference panel for the Women’s Health Initiative and UK Biobank studies). For studies sequenced using the OncoArray chip, samples with a low call rate (below 95%), with excessively low or high heterozygosity, or which were estimated to be below 80% European ancestry were excluded, as were individuals who were male, XO or XXY. In addition, SNPs with a minor allele frequency of <1% or differing significantly from the European reference panel frequency and with a call rate of <98%, and SNPs that could not be linked to the 1000 Genomes Project reference panel were excluded. Additionally, where close relatives or duplicates were identified, if between studies only the sample with the most recent genotyping was included and if within the same study the “case” was preferentially included in case-control pairs, and for case-case and control-control pairs only the sample with the higher call rate was included. For studies from the E2C2 consortium or the Women’s Health Initiative, standard quality measures were followed and SNPs with a minor allele frequency of <1% were excluded. For each case within the Women’s Health Initiative, five controls were also included, randomly selected from the same study.

*Statistical analyses*

Statistical analyses were carried out in R version 4.0.2. MR analyses were carried out using R packages “TwoSampleMR” and “MendelianRandomization” [1]. In some cases, GWAS data was accessed through the OpenGWAS database API [2, 3]. In order to perform Heterogeneity in Dependent Instruments (HEIDI) testing of the LDL cholesterol instrument, R package “GSMR” was used, with GCTA64 used to estimate the linkage disequilibrium correlation matrix [4].

*MR models*

The inverse-variance weighted Mendelian randomization (MR) model will be biased if any genetic variant included in the instrument is horizontally pleiotropic (or if there is not “balanced” horizontal pleiotropy across two or more variants in an instrument). Therefore, it is necessary to perform several other MR models with slightly different assumptions alongside this model as a sensitivity analysis.

In the weighted median model, the contribution of an individual genetic variant to the genetic instrument is weighted according to its distance from the median effect [5]. The weighted median model will provide an unbiased estimate when valid genetic variants (i.e. those lacking horizontal pleiotropic effects) contribute at least 50% of the information in an instrument [5].

The weighted mode model uses a modal estimate to generate unbiases causal effect estimates, and has another core assumption known as the Zero Modal Pleiotropy Assumption (ZEMPA) [6]. In this model, over 50% of the weight of the genetic instrument can be contributed by genetic variants which display horizontal pleiotropy, but the most common amount of pleiotropy must be zero [7]. This MR model suffers from low statistical power [8].

Two forms of the MR-Egger method were performed – the regression and the intercept tests. MR-Egger regression can provide an unbiased estimate even if all genetic variants included in the genetic instrument have horizontally pleiotropic effects [5]. In MR-Egger regression, there is an additional assumption known as the instrument strength independent of direct effect (InSIDE) assumption, which requires that the magnitude of directional horizontal pleiotropic effects shows no association with the effect size of genetic variants on an exposure [9]. MR-Egger regression has less statistical power than weighted median and weighted mode methods [7]. The MR-Egger intercept test gives an estimate of the strength of the effect of bias resulting from directional pleiotropy (i.e. unbalanced horizontal pleiotropy across an instrument) in the effect estimate. In the inverse-variance weighted model, the intercept of the linear regression of the variant-outcome association on the variant-exposure association is constrained to zero. The MR-Egger slope performs the same regression but the intercept is not constrained to zero, and instead the intercept is a measure of the strength of bias resulting from directional horizontal pleiotropy in the genetic instrument [9].

*Sex-specific and cis-only sensitivity analyses*

For endogenous sex hormones, we constructed instruments to proxy each exposure from genome-wide significant (*P* < 5 x 10^-8^) variants identified in analyses restricted to women. Although a combined-sex instrument was used for BMI initially, as a sensitivity analysis all analyses with nominal evidence for an association (*P* < 0.05) were repeated using genome-wide significant variants identified in the same analysis restricted to women. For CRP, as a sensitivity analysis we constructed an instrument replacing effect estimates and standard errors of two SNPs (rs1260326, rs1805096) showing evidence of interaction by sex in previously reported heterogeneity testing (Benjamini-Hochberg corrected FDR *P* < 5 × 10^−4^) with female-specific values. For fasting insulin, the instrument included one SNP (rs10195252) showing nominal evidence of interaction by sex in previously reported heterogeneity testing (*P* = 0.039), and, although this did not pass the Bonferroni threshold (*P* < 0.0026), a similar sensitivity analysis was performed for this instrument. For proteins which had previously been proxied by a genetic instrument including both *trans* and *cis* instruments (CRP, adiponectin and IGF-1), as a sensitivity analysis the MR investigating a causal relationship between proteins and endometrial cancer was repeated using *cis*-only instruments (defined as variants in or within 100kb of the gene encoding that protein on either side of the gene). For instruments using *cis*-only instruments, as a sensitivity analysis instrument construction was repeated using a 500kb window.

*Analyses to overcome weak instrument bias*

Several approaches were employed in an attempt to overcome weak instrument bias in the multivariable MR with fasting insulin and BMI on endometrial cancer risk (overall and endometrioid subtype). Firstly, as this estimate is likely an overestimation due to the use of univariable estimates, in an attempt to improve conditional instrument strength in the multivariable MR, the number of SNPs included in the model from the BMI instrument was reduced to 100 - initially the 100 SNPs with the smallest *P* value, and then 100 randomly selected SNPs to determine the effect of bias inherent in SNP selection by *P* value (**S35 Table**). Secondly, an alternative fasting insulin instrument with a greater sample size was used. Summary statistics for this GWAS were only available for fasting insulin adjusted for BMI. Therefore, in order to recalculate the proportion of the total effect mediated by fasting insulin, a two-step approach was implemented involving using the previous fasting insulin instrument (unadjusted for BMI) to determine the effect of BMI on fasting insulin, and the more recently published fasting insulin instrument (adjusted for BMI) to determine the effect of fasting insulin on endometrial cancer (**S36 Table**). Thirdly, due to continued weak instrument bias, the BMI instrument was again limited to 100 SNPs as before (**S30 Table**). Finally, different *P* value and r^2^ thresholds were used to reconstruct the fasting insulin instrument (both the initially used and updated instruments) (**S38 Table**).

The updated fasting insulin instrument which had been adjusted for BMI was used in all analyses to maximise instrument strength. Initially, a multivariable MR was performed with all three identified mediators, as well as BMI, to determine the effect of each mediator on endometrial cancer independently of one another (**S38 Table**). However, this model again suffered from weak instrument bias. Therefore, all possible pairings of mediators were instead included in separate multivariable MR models (i.e., three multivariable MR analyses were performed: (i) BMI, fasting insulin and SHBG on endometrial cancer risk, (ii) BMI, fasting insulin and bioavailable testosterone on endometrial cancer risk, and (iii) BMI, bioavailable testosterone and SHBG on endometrial cancer risk) (**S39 Table**). However, all three of these models continued to suffer from weak instrument bias. Therefore, a similar process of limiting the number of SNPs from the BMI instrument included in the model to 100 as previously performed was repeated, initially for the first model with all three mediators and BMI (**S40 Table**), and then for the separate models including pairs of mediators and BMI (**S41 Table**). These two further analyses still had conditional F-statistics below 10, suggesting potential presence of weak instrument bias. Consequently, BMI was removed from the model and the analysis was repeated with just the three mediators (**S42 Table**). Again, due to weak instrument bias, all possible pairings of mediators were instead included in separate multivariable MR models without BMI (**S34 Table**). Although this resulted in models with conditional F-statistics above 10 for fasting insulin and SHBG on endometrial cancer risk and bioavailable testosterone and SHBG on endometrial cancer risk, weak instrument bias persisted for fasting insulin and bioavailable testosterone on endometrial cancer risk (fasting insulin F=5).

As a sensitivity analysis to confirm the validity of the analyses in which the BMI instrument was limited to 100 SNPs, the causal effect of BMI on endometrial cancer (overall and by subtype) and on fasting insulin was recalculated with univariable MR analyses limited to the same 100 SNPs (both top and randomly selected) (**S43 Table**).

Although conditional F-statistics were sufficiently high for the other mediators investigated, there was potential sample overlap between a further two risk factors and BMI and endometrial cancer, meaning there could be genetic correlation between the summary statistics which had not been accounted for (**S5 Table**). Due to the unavailability of individual-level summary statistics, as a sensitivity analysis different levels of genetic correlation were investigated for their effect on the conditional F-statistics of these instruments (**S44 Table**).

### S3 Appendix. PubMed search terms for literature review.

For identifying previous MR studies of endometrial cancer we used the following search terms:

(("mendelian randomization"[All Fields] or "mendelian randomization") OR (mendelian randomization analysis[MeSH Terms])) AND ((endometrial cancer[MeSH Terms]) OR ("endometrial cancer"[All Fields]))

For identifying review papers detailing potential molecular mediators of the relationship between endometrial cancer and adiposity we used the following search terms:

(((adipose tissue[MeSH Terms]) AND (endometrial cancer[MeSH Terms])) AND (risk factors[MeSH Terms])) AND ("Review"[pt])

### S4 Table. Additional information on GWAS, including covariates adjusted for.

| **Exposure** | **GWAS** | **Covariates adjusted for** |
| --- | --- | --- |
| Adult BMI | Yengo et al. [1] | Age, sex, recruitment centre, genotyping batches, principal components |
| LDL-cholesterol | Willer et al. [2] | Age, sex, principal components. Individuals known to be on lipid-lowering medications were excluded |
| HDL-cholesterol | Willer et al. [2] | Age, sex, principal components. Individuals known to be on lipid-lowering medications were excluded |
| Triglycerides | Willer et al. [2] | Age, sex, principal components. Individuals known to be on lipid-lowering medications were excluded |
| Total serum cholesterol | Willer et al. [2] | Age, sex, principal components. Individuals known to be on lipid-lowering medications were excluded |
| Blood glucose | Neale et al. [3] | Age, sex, age-squared, age by sex, age by sex-squared, principal components |
| Fasting insulin (unadjusted for BMI) | Lagou et al. [4] | Age, study site and principal components. Individual study results were corrected for residual inflation of test statistics using genomic control |
| Fasting insulin (adjusted for BMI) | Chen et al. [5] | BMI, principal components. Only included adults who did not have diabetes and had never used anti-diabetic medication. Further study-specific adjustments included age, sex, centre, and cohort |
| IGF-1 (cis and trans variants) | Sinnott-Armstrong et al. [6] | Age, sex, age by sex, age-squared, principal components, assessment centre, month, Townsend deprivation indices and interactions with age and sex, genotyping array, blood draw time and interactions with age and sex, blood draw time-squared, urine sample time and interactions with age and sex, urine sample time-squared, sample dilution factor, fasting time and interactions with age and sex, fasting time-squared, interactions of blood draw time and urine sample with dilution factor |
| IGF-1 (cis variants) | Larsson et al. [7] | Age, sex, principal components |
| IL-6 | Georgakis et al. [8] | None |
| Adiponectin (cis and trans variants) | Locke et al. [9] | Type 2 diabetes |
| Adiponectin (cis variants) | Locke et al. [9] | Type 2 diabetes |
| Leptin | Folkersen et al. [10] | Dependent on study: IMPROVE – site, age, sex, olink batch, individuals with gender mismatch, heterozygosity and cryptic relatedness were excluded. STANLEY – age, sex, MDS components, olink batch, individuals with gender mismatch, heterozygosity and cryptic relatedness were excluded. EpiHealth – age, sex, olink plate, MDS component 1-5, individuals with gender mismatch, heterozygosity (more than 5 SD from the mean), cryptic relatedness and ethnic outliers were excluded. PIVUS and ULSAM – age, sex, olink plate, storage time, MDS component 1-2, individuals with gender mismatch, heterozygosity (more than 5 SD from the mean), cryptic relatedness and ethnic outliers were excluded. INTERVAL – age, sex, season, plate, bleed to processing time (days), MDS components 1-3. Individuals with gender mismatch, heterozygosity and genetic outliers through multidimentional scaling were excluded. STABILITY – age, sex, four principal components, individuals with gender mismatch were excluded. Estonian BioBank – age, sex, olink plate, MDS components 1-10, individuals with gender mismatch, ethnic outliers, heterozygosity (more than 3 SD form the mean) and cryptic relatedness were excluded. ORCADES – age, sex, array, time in storage, season, plate number, plate row and column, 10 prinicpal components, individuals with ethnic outliers, duplicates, gender mismatch, and excess IBS incompatible with pedigree were excluded. VIS – age and sex. |
| CRP (cis and trans variants) | Ligthart et al. [11] | Age, sex, population substructure, relatedness where relevant. Individuals with auto-immune diseases, individuals taking immune-modulating agents, and individuals with CRP amounts 4 SD or more away from the mean were excluded |
| CRP (cis variants) | C Reactive Protein Coronary Heart Disease Genetics Collaboration (CCGC) [12] | Ancestry |
| Oestradiol | Neale et al. [3] | Age, sex, age-squared, age by sex, age by sex-squared, principal components |
| Total testosterone | Ruth et al. [13] | Genotyping chip, age at baseline, ten genetically derived principal components, fasting time, age, centre, chip/release of genetic data |
| Bioavailable testosterone | Ruth et al. [13] | Genotyping chip, age at baseline, ten genetically derived principal components, age, dilution, batch, minutes since blood draw, time of blood draw, menopause, operation status. Individuals who self-reported taking hormone-based medication including HRT and oral contraception at the time of the initial visit were excluded |
| SHBG | Ruth et al. [13] | Genotyping chip, age at baseline, ten genetically derived principal components, age, dilution, batch, mins since blood draw, time of blood draw, menopause, operation status. Individuals who self-reported taking hormone-based medication including HRT and oral contraception at the time of the initial visit were excluded |

BMI = body mass index, LDL = low-density lipoprotein, HDL = high density lipoprotein, IGF-1 = Insulin-like growth factor-1, IL-6 = interleukin-6, CRP = C-reactive protein, SHBG = sex hormone-binding globulin, LD = linkage disequilibrium.

### S5 Table. Sample overlap between GWAS.

| **Exposure** | **Outcome** | **Sample overlap** |
| --- | --- | --- |
| BMI | Overall endometrial cancer | 55.20% |
| SHBG | Overall endometrial cancer | 52.22% |
| Fasting insulin | Overall endometrial cancer | 0.00% |
| Total testosterone | Overall endometrial cancer | 52.22% |
| Bioavailable testosterone | Overall endometrial cancer | 52.22% |
| LDL-cholesterol | Overall endometrial cancer | 0.00% |
| IGF-1 | Overall endometrial cancer | 52.22% |
| Blood glucose | Overall endometrial cancer | 52.22% |
| IL-6 | Overall endometrial cancer | 0.00% |
| Adiponectin | Overall endometrial cancer | 0.00% |
| CRP | Overall endometrial cancer | 0.71% |
| Triglyceride | Overall endometrial cancer | 0.00% |
| Total serum cholesterol | Overall endometrial cancer | 0.00% |
| HDL-cholesterol | Overall endometrial cancer | 0.00% |
| Leptin | Overall endometrial cancer | 0.00% |
| cis IGF-1 | Overall endometrial cancer | 52.22% |
| cis CRP | Overall endometrial cancer | 0.00% |
| cis adiponectin | Overall endometrial cancer | 0.00% |
| BMI | Non-endometrioid endometrial cancer | 58.70% |
| BMI | Endometrioid endometrial cancer | 54.95% |
| SHBG | Endometrioid endometrial cancer | 53.38% |
| Fasting insulin | Endometrioid endometrial cancer | 0.00% |
| Total testosterone | Endometrioid endometrial cancer | 53.38% |
| Bioavailable testosterone | Endometrioid endometrial cancer | 53.38% |
| LDL-cholesterol | Endometrioid endometrial cancer | 0.00% |
| IGF-1 | Endometrioid endometrial cancer | 53.38% |
| Blood glucose | Endometrioid endometrial cancer | 53.38% |
| IL-6 | Endometrioid endometrial cancer | 0.00% |
| Adiponectin | Endometrioid endometrial cancer | 0.00% |
| CRP | Endometrioid endometrial cancer | 0.74% |
| Triglyceride | Endometrioid endometrial cancer | 0.00% |
| Total serum cholesterol | Endometrioid endometrial cancer | 0.00% |
| HDL-cholesterol | Endometrioid endometrial cancer | 0.00% |
| Leptin | Endometrioid endometrial cancer | 0.00% |
| cis IGF-1 | Endometrioid endometrial cancer | 53.38% |
| cis CRP | Endometrioid endometrial cancer | 0.00% |
| cis Adiponectin | Endometrioid endometrial cancer | 0.00% |
| BMI | SHBG | 62.40% |
| BMI | Fasting insulin | 3.40% |
| BMI | Total testosterone | 62.40% |
| BMI | Bioavailable testosterone | 62.40% |
| BMI | LDL-cholesterol | 7.88% |
| BMI | Total serum cholesterol | 8.45% |

BMI = body mass index, LDL = low-density lipoprotein, HDL = high density lipoprotein, IGF-1 = Insulin-like growth factor-1, IL-6 = interleukin-6, CRP = C-reactive protein, SHBG = sex hormone-binding globulin, LD = linkage disequilibrium.

### S6 Table. Conditional F-statistics for multivariable Mendelian randomization of BMI and mediators on endometrial cancer risk.

| **Mediator** | **Conditional F-statistics** |
| --- | --- |
| Fasting insulin | BMI: 6  Fasting insulin: 2 |
| Bioavailable testosterone | BMI: 43  Bioavailable testosterone: 13 |
| SHBG | BMI: 52  SHBG: 25 |
| C-reactive protein | BMI: 61  C-reactive protein: 24 |

BMI = body mass index, SHBG = sex hormone-binding globulin.

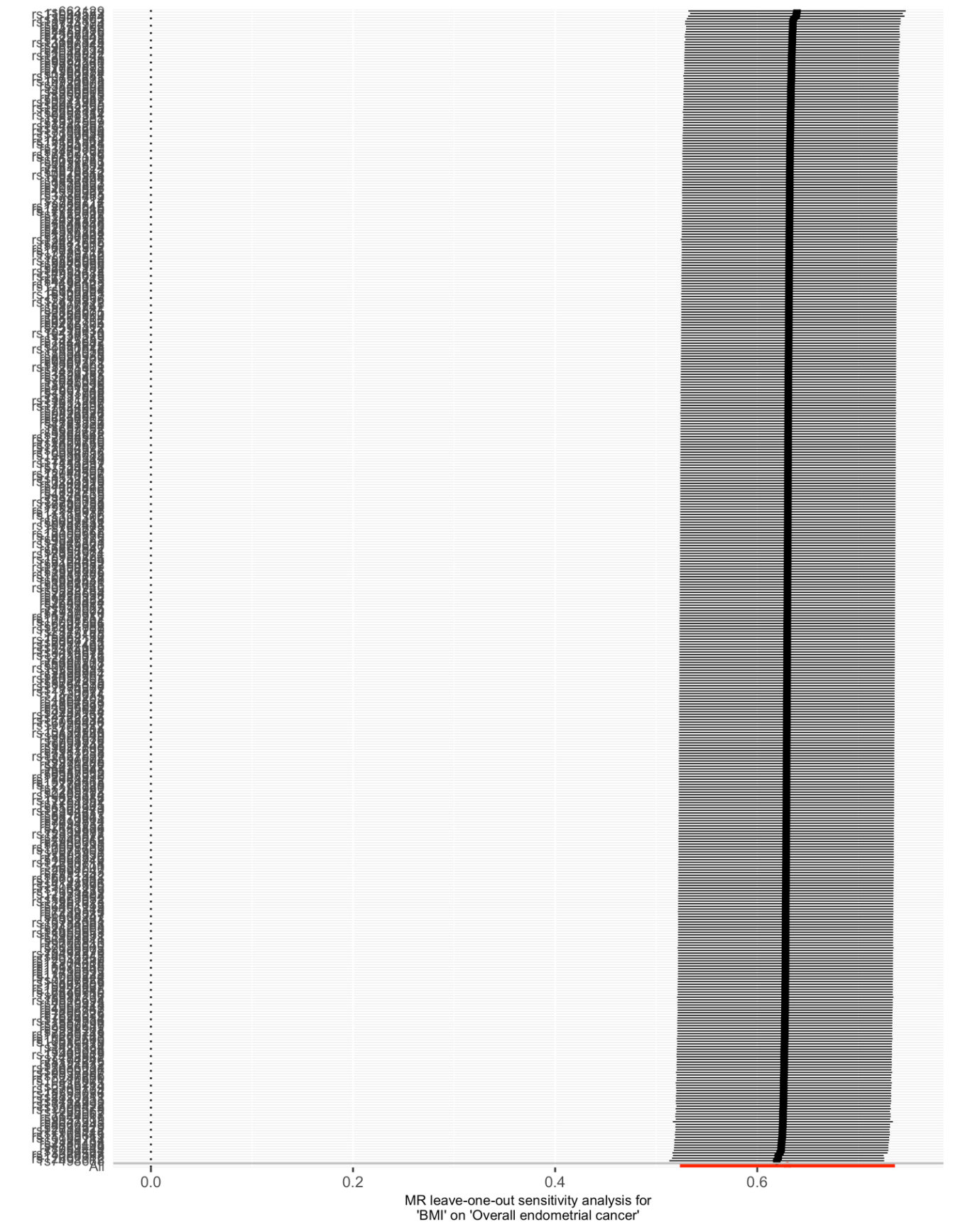

### S7 Figure. Leave-one-out analysis for MR examining the effect of adult BMI on overall endometrial cancer risk.

### S8 Table. Results of female BMI sensitivity analysis MR.

| **Outcome** | **Method** | **OR/Effect estimate (95% CI)** | ***P* value** |
| --- | --- | --- | --- |
| Overall endometrial cancer | IVW | 1.76 (1.46 to 2.11) | 1.36 x 10^-9^ |
|  | Weighted median | 1.75 (1.39 to 2.21) | 2.72 x 10^-6^ |
|  | Weighted mode | 1.69 (1.25 to 2.28) | 1.53 x 10^-3^ |
|  | MR Egger | 1.33 (0.81 to 2.18) | 2.67 x 10^-1^ |
| Endometrioid endometrial cancer | IVW | 1.76 (1.44 to 2.17) | 5.90 x 10^-8^ |
|  | Weighted median | 1.77 (1.34 to 2.32) | 4.92 x 10^-5^ |
|  | Weighted mode | 1.69 (1.18 to 2.42) | 6.63 x 10^-3^ |
|  | MR Egger | 1.58 (0.90 to 2.79) | 1.21 x 10^-1^ |
| Non-endometrioid endometrial cancer | IVW | 1.86 (1.16 to 2.98) | 9.60 x 10^-3^ |
|  | Weighted median | 1.54 (0.77 to 3.05) | 2.20 x 10^-1^ |
|  | Weighted mode | 1.38 (0.57 to 3.35) | 4.86 x 10^-1^ |
|  | MR Egger | 0.94 (0.27 to 3.36) | 9.29 x 10^-1^ |
| Sex Hormone-Binding Globulin (SHBG) | IVW | -0.10 (-0.13 to -0.08) | 2.58 x 10^-14^ |
|  | Weighted median | -0.11 (-0.14 to -0.09) | 3.22 x 10^-18^ |
|  | Weighted mode | -0.12 (-0.15 to -0.09) | 3.40 x 10^-9^ |
|  | MR Egger | -0.12 (-0.19 to -0.05) | 2.73 x 10^-3^ |
| Fasting Insulin | IVW | 0.18 (0.14 to 0.21) | 1.06 x 10^-23^ |
|  | Weighted median | 0.16 (0.12 to 0.20) | 2.70 x 10^-13^ |
|  | Weighted mode | 0.14 (0.06 to 0.23) | 1.66 x 10^-3^ |
|  | MR Egger | 0.18 (0.09 to 0.27) | 6.44 x 10^-4^ |
| Total Testosterone | IVW | 0.05 (-0.03 to 0.13) | 2.19 x 10^-1^ |
|  | Weighted median | 0.05 (-0.03 to 0.13) | 2.84 x 10^-1^ |
|  | Weighted mode | -0.02 (-0.18 to 0.13) | 7.88 x 10^-1^ |
|  | MR Egger | -0.20 (-0.41 to 0.01) | 1.00 x 10^-1^ |
| Bioavailable Testosterone | IVW | 0.15 (0.09 to 0.21) | 1.23 x 10^-6^ |
|  | Weighted median | 0.12 (0.03 to 0.21) | 6.99 x 10^-3^ |
|  | Weighted mode | 0.08 (-0.05 to 0.20) | 2.52 x 10^-1^ |
|  | MR Egger | 0.02 (-0.17 to 0.20) | 8.69 x 10^-1^ |
| Total Serum Cholesterol | IVW | -0.05 (-0.12 to 0.02) | 1.50 x 10^-1^ |
|  | Weighted median | -0.06 (-0.12 to 0.00) | 4.00 x 10^-2^ |
|  | Weighted mode | -0.08 (-0.15 to -0.01) | 3.84 x 10^-2^ |
|  | MR Egger | -0.11 (-0.29 to 0.06) | 2.20 x 10^-1^ |
| C-reactive protein | IVW | 0.35 (0.31 to 0.39) | 8.07 x 10^-70^ |
|  | Weighted median | 0.37 (0.31 to 0.43) | 3.84 x 10^-35^ |
|  | Weighted mode | 0.39 (0.31 to 0.47) | 1.92 x 10^-11^ |
|  | MR Egger | 0.36 (0.26 to 0.47) | 1.24 x 10^-7^ |
| LDL-cholesterol | IVW | 0.01 (-0.04 to 0.06) | 7.30 x 10^-1^ |
|  | Weighted median | 0.01 (-0.06 to 0.08) | 7.16 x 10^-1^ |
|  | Weighted mode | 0.01 (-0.09 to 0.11) | 8.20 x 10^-1^ |
|  | MR Egger | -0.11 (-0.24 to 0.03) | 1.29 x 10^-1^ |

OR per SD (4.7 kg/m^2^) increase in BMI. Effect estimates represent change in inverse normal transformed nmol/L SHBG, natural log transformed pmol/L fasting insulin, inverse normal transformed nmol/L total testosterone, inverse normal transformed nmol/L bioavailable testosterone, SD (41.7 mg/dL) total serum cholesterol, natural log transformed mg/L CRP, and SD (38.7 mg/dL) increase LDL-cholesterol.

### S9 Table. Results from sensitivity analyses examining the influence of Winner’s curse on GWAS with overlapping samples.

| **Exposure** | **GWAS** | ***P* value used for instrument construction** | **Outcome** | **Method** | **OR/Effect estimate**  **(95% CI)** | ***P* value** |
| --- | --- | --- | --- | --- | --- | --- |
| IGF-1 | Sinnott-Armstrong et al. [1] | 5 x 10^-9^ | Overall endometrial cancer | IVW | 0.96 (0.84 to 1.09) | 5.27 x 10^-1^ |
|  |  |  |  | Weighted median | 1.03 (0.85 to 1.24) | 7.94 x 10^-1^ |
|  |  |  |  | Weighted mode | 1.27 (0.90 to 1.80) | 1.79 x 10^-1^ |
|  |  |  |  | MR Egger | 1.19 (0.84 to 1.69) | 3.32 x 10^-1^ |
|  |  |  | Endometrioid endometrial cancer | IVW | 0.91 (0.78 to 1.07) | 2.70 x 10^-1^ |
|  |  |  |  | Weighted median | 1.02 (0.81 to 1.27) | 8.88 x 10^-1^ |
|  |  |  |  | Weighted mode | 1.35 (0.93 to 1.96) | 1.20 x 10^-1^ |
|  |  |  |  | MR Egger | 1.30 (0.86 to 1.97) | 2.21 x 10^-1^ |
|  |  | 5 x 10^-10^ | Overall endometrial cancer | IVW | 0.97 (0.85 to 1.11) | 6.27 x 10^-1^ |
|  |  |  |  | Weighted median | 1.02 (0.85 to 1.24) | 8.02 x 10^-1^ |
|  |  |  |  | Weighted mode | 1.29 (0.89 to 1.87) | 1.77 x 10^-1^ |
|  |  |  |  | MR Egger | 1.17 (0.82 to 1.68) | 3.86 x 10^-1^ |
|  |  |  | Endometrioid endometrial cancer | IVW | 0.92 (0.78 to 1.08) | 2.86 x 10^-1^ |
|  |  |  |  | Weighted median | 1.02 (0.81 to 1.27) | 8.73 x 10^-1^ |
|  |  |  |  | Weighted mode | 1.35 (0.92 to 1.96) | 1.24 x 10^-1^ |
|  |  |  |  | MR Egger | 1.26 (0.82 to 1.93) | 2.90 x 10^-1^ |
| Blood glucose | Kettunen et al. [2] | 5 x 10^-8^ | Overall endometrial cancer | IVW | 1.11 (0.83 to 1.48) | 4.80 x 10^-1^ |
|  |  |  |  | Weighted median | 1.04 (0.83 to 1.30) | 7.53 x 10^-1^ |
|  |  |  |  | Weighted mode | 1.00 (0.79 to 1.27) | 9.98 x 10^-1^ |
|  |  |  |  | MR Egger | 0.46 (0.19 to 1.11) | 3.34 x 10^-1^ |
|  |  |  | Endometrioid endometrial cancer | IVW | 1.13 (0.82 to 1.55) | 4.60 x 10^-1^ |
|  |  |  |  | Weighted median | 1.04 (0.79 to 1.37) | 7.68 x 10^-1^ |
|  |  |  |  | Weighted mode | 1.01 (0.76 to 1.33) | 9.67 x 10^-1^ |
|  |  |  |  | MR Egger | 0.43 (0.15 to 1.24) | 3.61 x 10^-1^ |
| SHBG | Ruth et al. [3] | 5 x 10^-9^ | Overall endometrial cancer | IVW | 0.71 (0.59 to 0.86) | 3.88 x 10^-4^ |
|  |  |  |  | Weighted median | 0.64 (0.48 to 0.85) | 2.00 x 10^-3^ |
|  |  |  |  | Weighted mode | 0.67 (0.51 to 0.87) | 3.39 x 10^-3^ |
|  |  |  |  | MR Egger | 0.63 (0.46 to 0.86) | 4.17 x 10^-3^ |
|  |  |  | Endometrioid endometrial cancer | IVW | 0.67 (0.54 to 0.82) | 1.87 x 10^-4^ |
|  |  |  |  | Weighted median | 0.60 (0.43 to 0.85) | 4.40 x 10^-3^ |
|  |  |  |  | Weighted mode | 0.58 (0.42 to 0.80) | 1.18 x 10^-3^ |
|  |  |  |  | MR Egger | 0.62 (0.44 to 0.88) | 7.72 x 10^-3^ |
|  |  | 5 x 10^-10^ | Overall endometrial cancer | IVW | 0.68 (0.56 to 0.82) | 9.02 x 10^-5^ |
|  |  |  |  | Weighted median | 0.64 (0.48 to 0.86) | 3.00 x 10^-3^ |
|  |  |  |  | Weighted mode | 0.67 (0.51 to 0.87) | 3.51x 10^-3^ |
|  |  |  |  | MR Egger | 0.64 (0.47 to 0.89) | 7.97 x 10^-3^ |
|  |  |  | Endometrioid endometrial cancer | IVW | 0.64 (0.52 to 0.79) | 2.77 x 10^-5^ |
|  |  |  |  | Weighted median | 0.60 (0.44 to 0.84) | 2.32 x 10^-3^ |
|  |  |  |  | Weighted mode | 0.59 (0.42 to 0.84) | 4.34 x 10^-3^ |
|  |  |  |  | MR Egger | 0.63 (0.44 to 0.89) | 9.98 x 10^-3^ |
| Bioavailable testosterone | Ruth et al. [3] | 5 x 10^-9^ | Overall endometrial cancer | IVW | 1.47 (1.29 to 1.67) | 8.58 x 10^-9^ |
|  |  |  |  | Weighted median | 1.48 (1.21 to 1.81) | 1.61 x 10^-4^ |
|  |  |  |  | Weighted mode | 1.53 (1.17 to 1.99) | 2.24 x 10^-3^ |
|  |  |  |  | MR Egger | 1.90 (1.44 to 2.52) | 8.58 x 10^-9^ |
|  |  |  | Endometrioid endometrial cancer | IVW | 1.46 (1.26 to 1.70) | 7.41 x 10^-7^ |
|  |  |  |  | Weighted median | 1.41 (1.13 to 1.76) | 2.05 x 10^-3^ |
|  |  |  |  | Weighted mode | 1.58 (1.19 to 2.09) | 1.94 x 10^-3^ |
|  |  |  |  | MR Egger | 1.70 (1.23 to 2.36) | 1.84 x 10^-3^ |
|  |  | 5 x 10^-10^ | Overall endometrial cancer | IVW | 1.48 (1.29 to 1.70) | 2.01 x 10^-8^ |
|  |  |  |  | Weighted median | 1.48 (1.20 to 1.81) | 1.77 x 10^-4^ |
|  |  |  |  | Weighted mode | 1.52 (1.16 to 1.98) | 2.81 x 10^-3^ |
|  |  |  |  | MR Egger | 2.04 (1.52 to 2.73) | 6.92 x 10^-6^ |
|  |  |  | Endometrioid endometrial cancer | IVW | 1.48 (1.26 to 1.73) | 9.81 x 10^-7^ |
|  |  |  |  | Weighted median | 1.42 (1.13 to 1.78) | 2.87 x 10^-3^ |
|  |  |  |  | Weighted mode | 1.63 (1.22 to 2.17) | 1.21 x 10^-3^ |
|  |  |  |  | MR Egger | 1.82 (1.30 to 2.55) | 8.18 x 10^-4^ |
| Total testosterone | Ruth et al. [3] | 5 x 10^-9^ | Overall endometrial cancer | IVW | 1.60 (1.38 to 1.85) | 3.76 x 10^-10^ |
|  |  |  |  | Weighted median | 1.63 (1.35 to 1.98) | 5.78 x 10^-7^ |
|  |  |  |  | Weighted mode | 1.73 (1.35 to 2.22) | 3.86 x 10^-5^ |
|  |  |  |  | MR Egger | 1.82 (1.35 to 2.46) | 1.77 x 10^-4^ |
|  |  |  | Endometrioid endometrial cancer | IVW | 1.58 (1.31 to 1.89) | 8.47 x 10^-7^ |
|  |  |  |  | Weighted median | 1.81 (1.43 to 2.28) | 6.00 x 10^-7^ |
|  |  |  |  | Weighted mode | 1.88 (1.44 to 2.44) | 1.03 x 10^-5^ |
|  |  |  |  | MR Egger | 1.81 (1.25 to 2.63) | 2.35 x 10^-3^ |
|  |  | 5 x 10^-10^ | Overall endometrial cancer | IVW | 1.60 (1.38 to 1.86) | 5.87 x 10^-10^ |
|  |  |  |  | Weighted median | 1.64 (1.36 to 1.98) | 2.49 x 10^-7^ |
|  |  |  |  | Weighted mode | 1.74 (1.38 to 2.19) | 1.21 x 10^-5^ |
|  |  |  |  | MR Egger | 1.81 (1.34 to 2.44) | 2.55 x 10^-4^ |
|  |  |  | Endometrioid endometrial cancer | IVW | 1.58 (1.32 to 1.90) | 6.20 x 10^-7^ |
|  |  |  |  | Weighted median | 1.81 (1.42 to 2.30) | 1.22 x 10^-6^ |
|  |  |  |  | Weighted mode | 1.89 (1.43 to 2.50) | 3.03 x 10^-5^ |
|  |  |  |  | MR Egger | 1.80 (1.25 to 2.61) | 2.52 x 10^-3^ |
| BMI | Locke et al.[4] | 5 x 10^-8^ | Overall endometrial cancer | IVW | 1.88 (1.62 to 2.19) | 3.41 x 10^-16^ |
|  |  |  |  | Weighted median | 1.79 (1.39 to 2.29) | 4.59 x 10^-6^ |
|  |  |  |  | Weighted mode | 1.75 (1.33 to 2.29) | 1.31 x 10^-4^ |
|  |  |  |  | MR Egger | 1.65 (1.14 to 2.39) | 9.85 x 10^-3^ |
|  |  |  | Endometrioid endometrial cancer | IVW | 1.98 (1.64 to 2.38) | 8.22 x 10^-13^ |
|  |  |  |  | Weighted median | 1.93 (1.46 to 2.56) | 4.35 x 10^-6^ |
|  |  |  |  | Weighted mode | 1.81 (1.31 to 2.50) | 5.83 x 10^-4^ |
|  |  |  |  | MR Egger | 1.77 (1.13 to 2.78) | 1.52 x 10^-2^ |
|  |  |  | SHBG | IVW | 0.89 (0.87 to 0.91) | 6.99 x 10^-20^ |
|  |  |  |  | Weighted median | 0.88 (0.86 to 0.90) | 3.19 x 10^-27^ |
|  |  |  |  | Weighted mode | 0.87 (0.85 to 0.90) | 2.34 x 10^-13^ |
|  |  |  |  | MR Egger | 0.91 (0.85 to 0.96) | 2.48 x 10^-3^ |
|  |  |  | Bioavailable testosterone | IVW | 1.20 (1.14 to 1.26) | 3.77 x 10^-13^ |
|  |  |  |  | Weighted median | 1.12 (1.06 to 1.18) | 4.64 x 10^-5^ |
|  |  |  |  | Weighted mode | 1.11 (1.05 to 1.17) | 2.73 x 10^-4^ |
|  |  |  |  | MR Egger | 1.09 (0.97 to 1.23) | 1.59 x 10^-1^ |
|  |  |  | Total testosterone | IVW | 1.07 (1.03 to 1.12) | 2.48 x 10^-3^ |
|  |  |  |  | Weighted median | 1.01 (0.96 to 1.05) | 7.82 x 10^-1^ |
|  |  |  |  | Weighted mode | 0.98 (0.93 to 1.04) | 4.88 x 10^-1^ |
|  |  |  |  | MR Egger | 0.95 (0.86 to 1.07) | 4.07 x 10^-1^ |

ORs are shown per increase in nmol/L IGF-1, SD (0.22 mmol/L) glucose, inverse normal transformed nmol/L SHBG, inverse normal transformed nmol/L bioavailable testosterone, inverse normal transformed nmol/L total testosterone, SD (4.6 kg/m^2^) BMI. BMI = body mass index, SHBG = sex hormone-binding globulin, IVW = inverse weighted variance, IGF-1 = insulin-like growth factor-1.

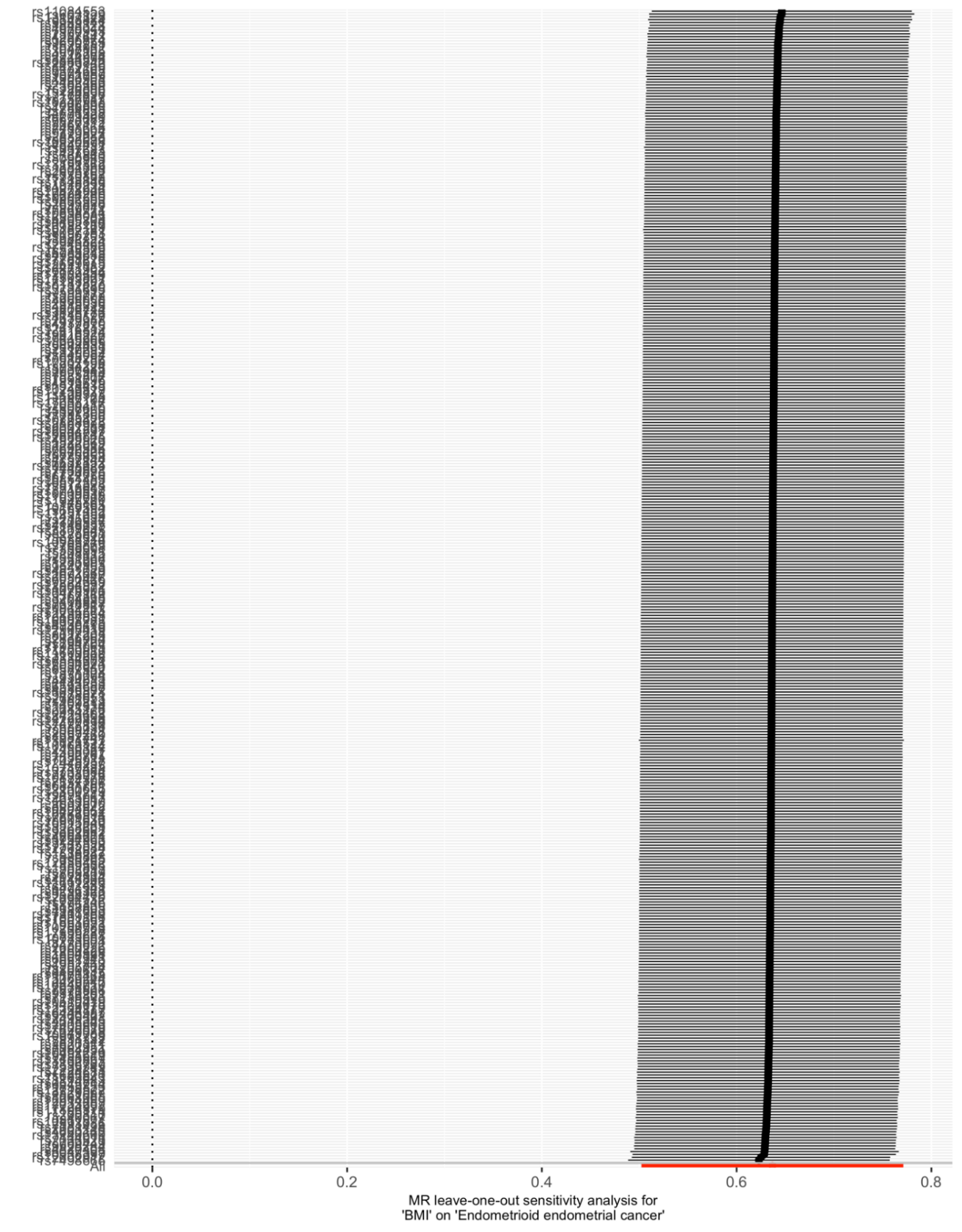

### S10 Figure. Leave-one-out analysis for MR examining the effect of adult BMI on endometrioid endometrial cancer risk.

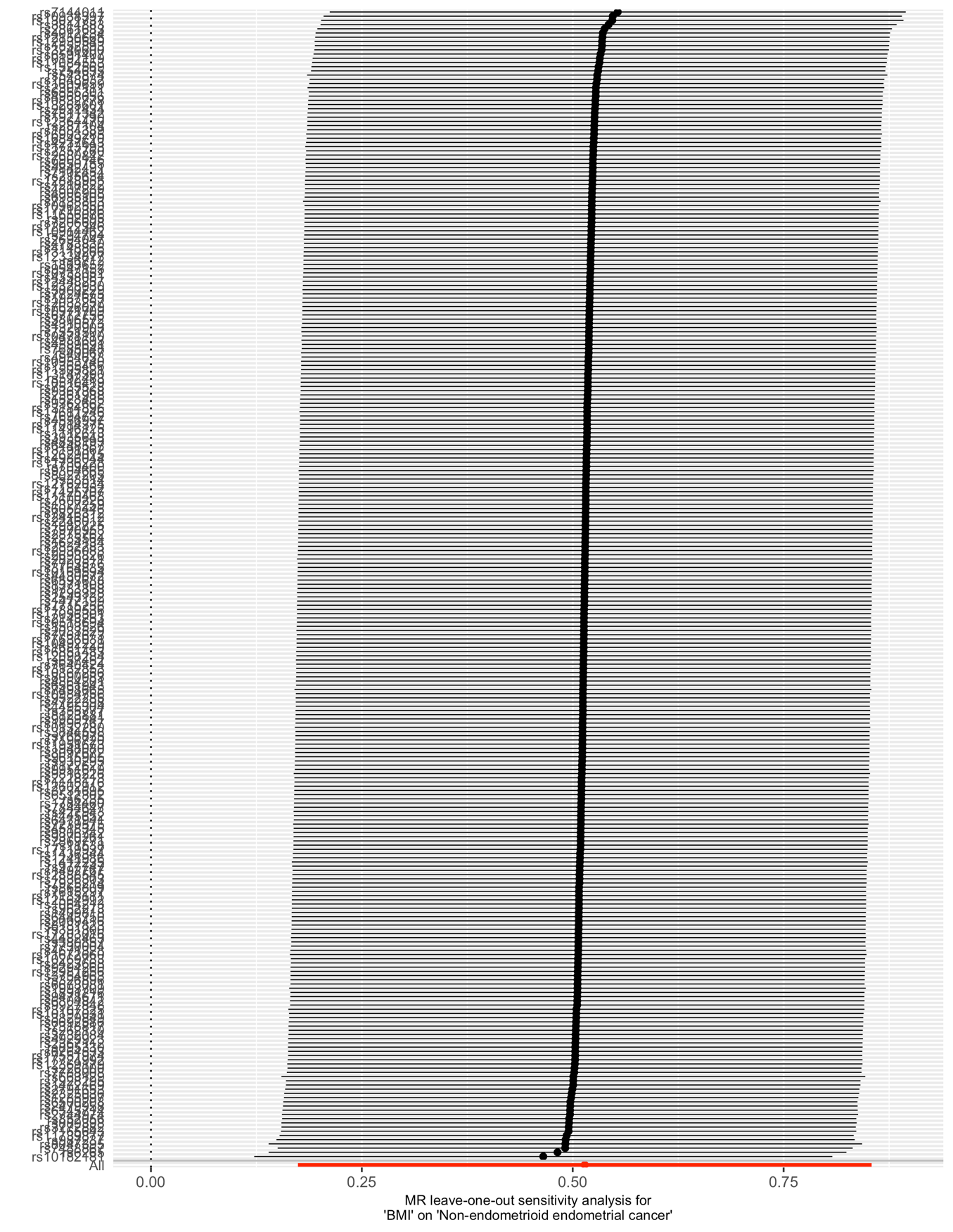

### S11 Figure. Leave-one-out analysis for MR examining the effect of adult BMI on non-endometrioid endometrial cancer risk.

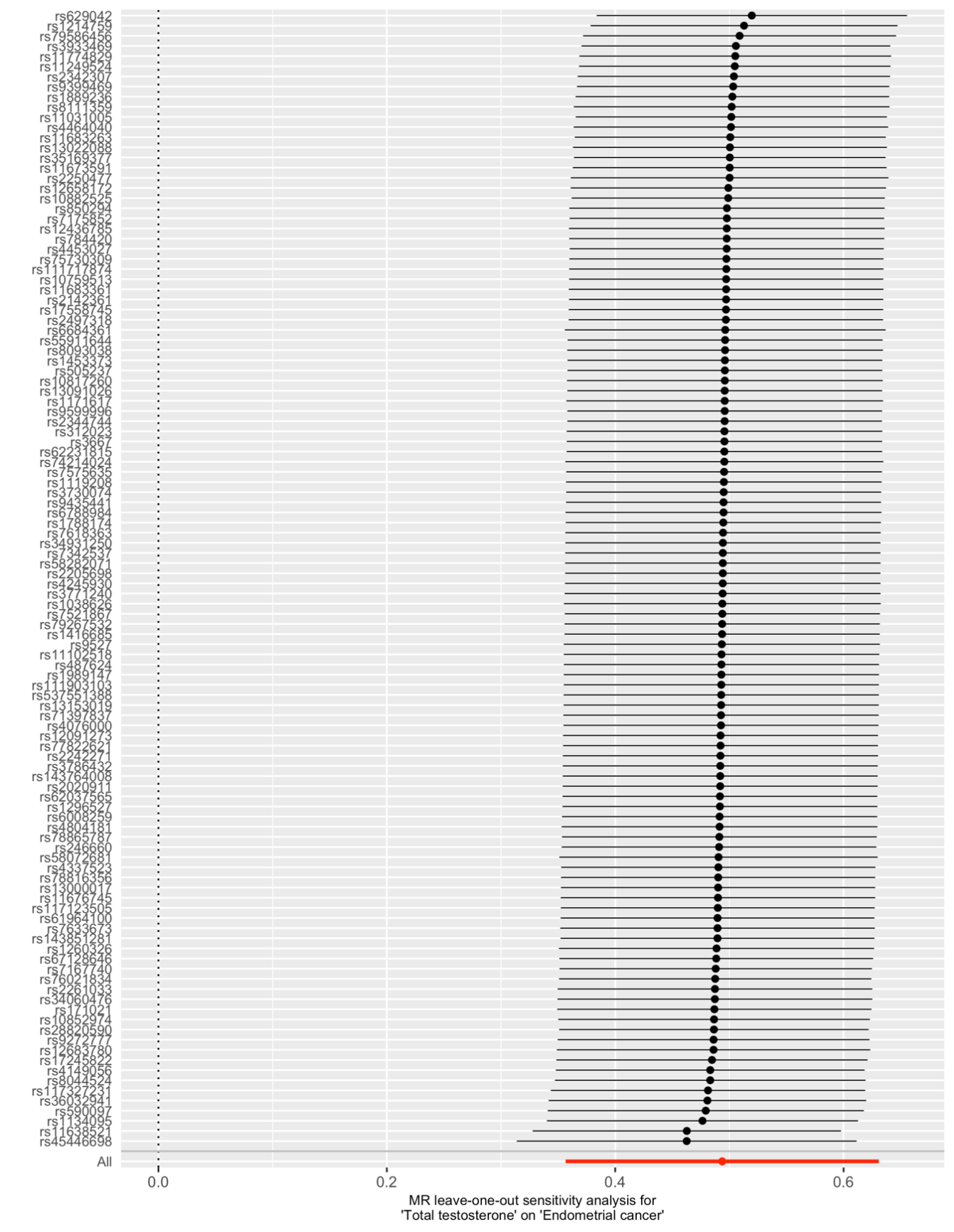

### S12 Figure. Leave-one-out analysis for MR examining the effect of total testosterone level on endometrial cancer risk.

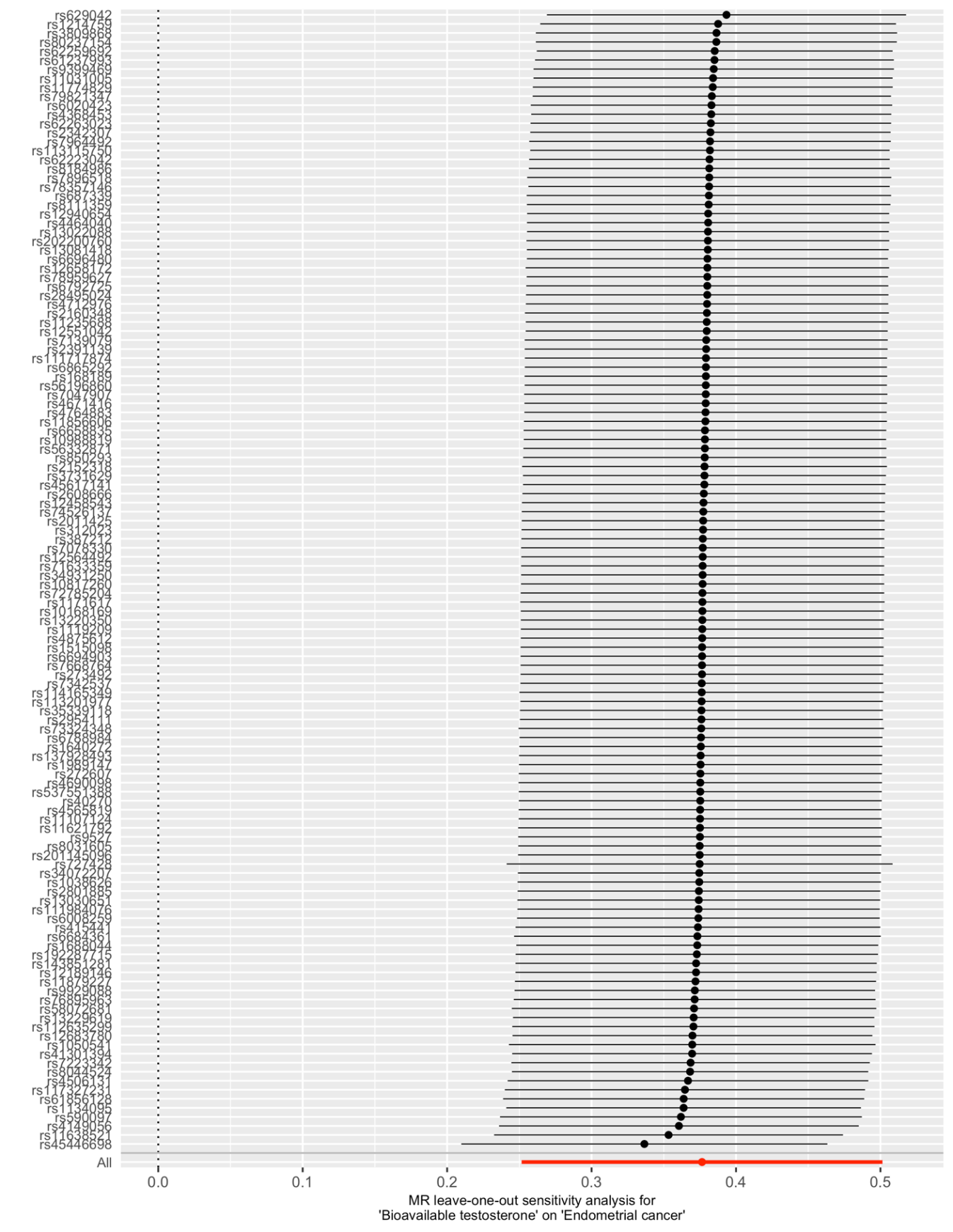

### S13 Figure. Leave-one-out analysis for MR examining the effect of bioavailable testosterone level on endometrial cancer risk.

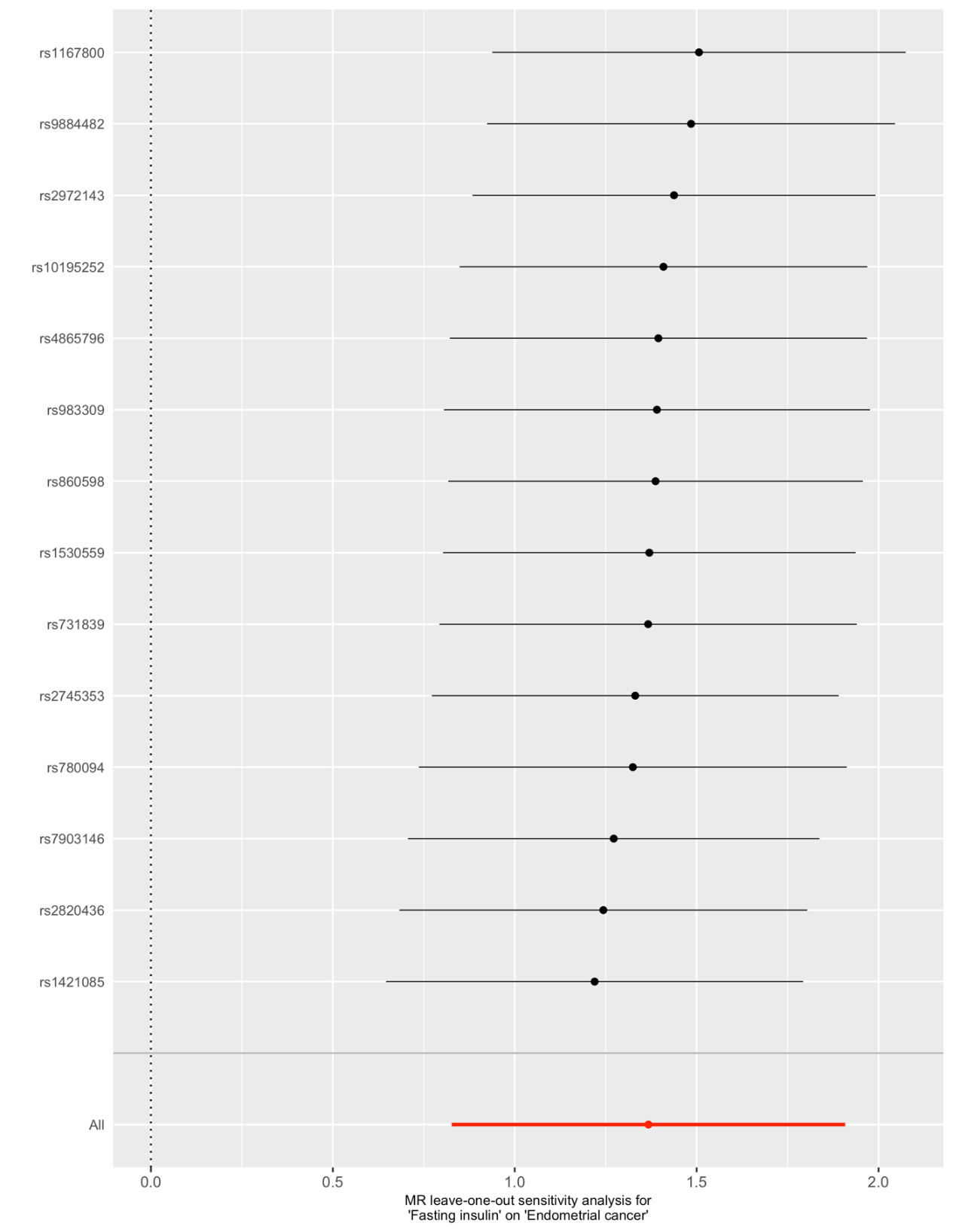

### **S14 Figure. Leave-one-out analysis for MR examining the effect of fasting insulin level on endometrial cancer risk.**

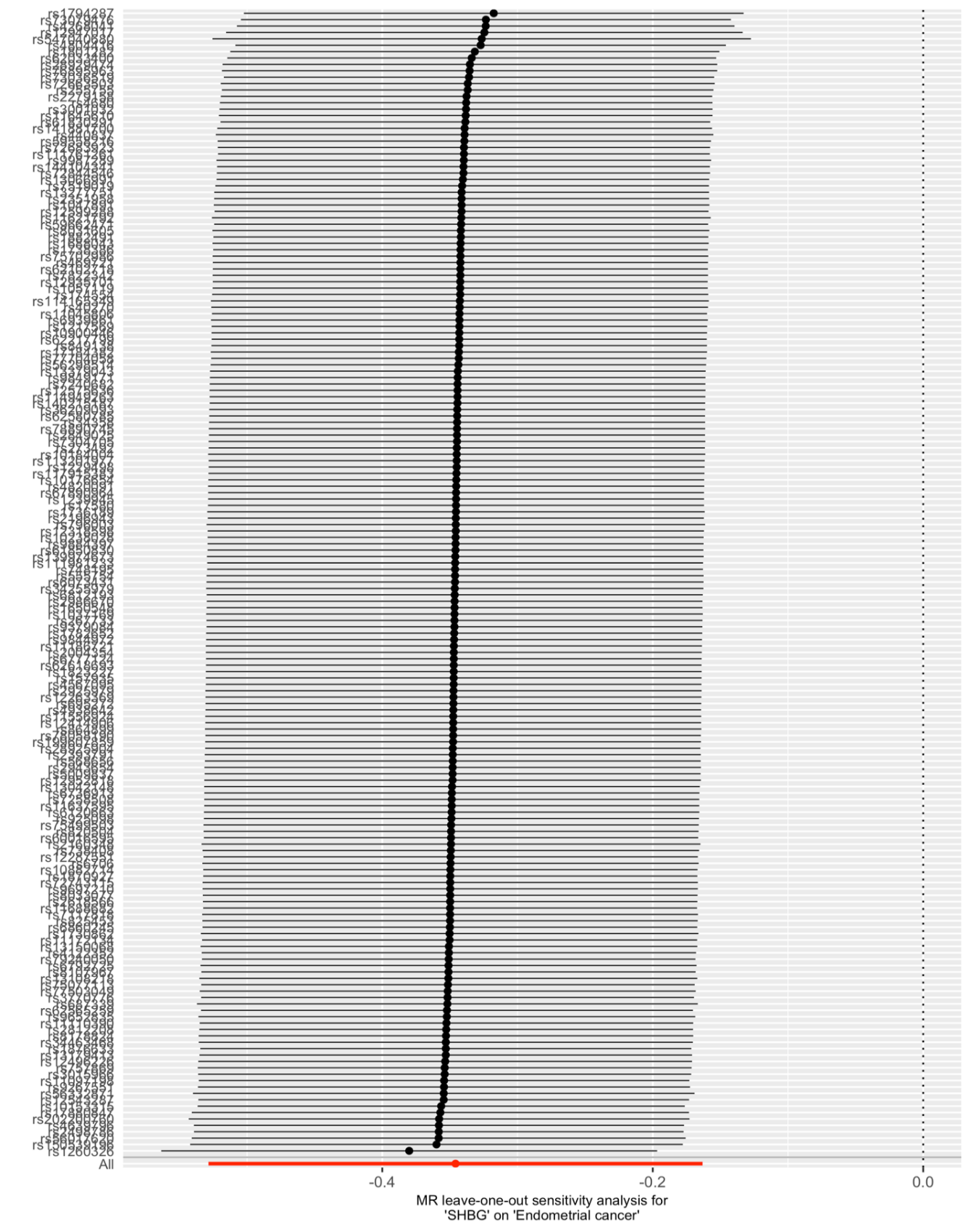

### S15 Figure. Leave-one-out analysis for MR examining the effect of SHBG level on endometrial cancer risk.

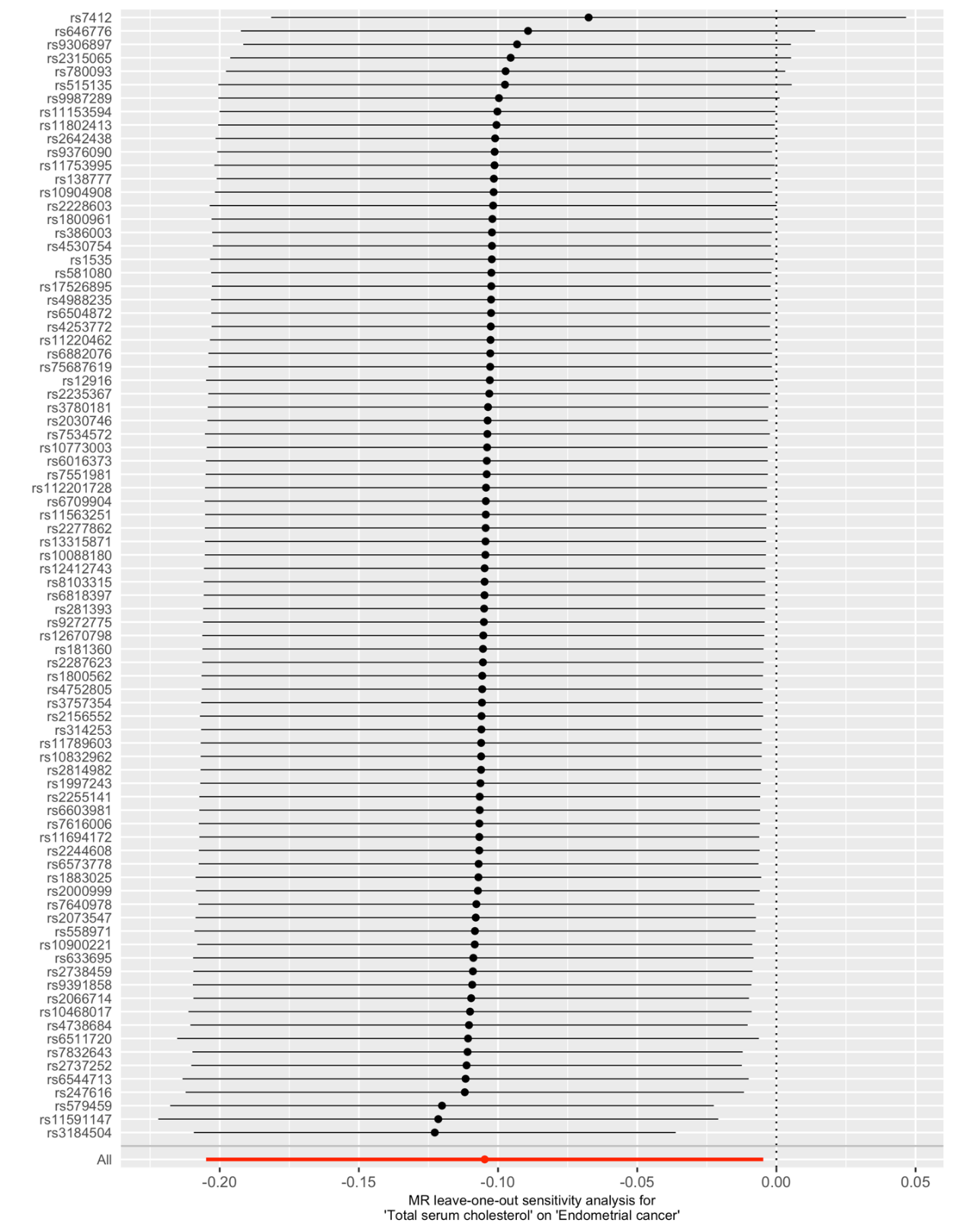

### S16 Figure. Leave-one-out analysis for MR examining the effect of total serum cholesterol level on endometrial cancer risk.

### S17 Table. Results of female-specific SNP fasting insulin sensitivity analysis MR.

| **Exposure** | **Outcome** | **Method** | **OR/Effect estimate (95% CI)** | ***P* value** |
| --- | --- | --- | --- | --- |
| Fasting insulin | Overall endometrial cancer | IVW | 3.71 (2.17 to 6.35) | 1.77 x 10^-6^ |
|  |  | Weighted median | 3.31 (1.56 to 6.99) | 1.76 x 10^-3^ |
|  |  | Weighted mode | 3.28 (0.82 to 13.07) | 1.15 x 10^-1^ |
|  |  | MR Egger | 5.54 (0.37 to 83.13) | 2.39 x 10^-1^ |
|  | Endometrioid endometrial cancer | IVW | 4.29 (2.11 to 8.71) | 5.83 x 10^-5^ |
|  |  | Weighted median | 3.41 (1.39 to 8.35) | 7.17 x 10^-3^ |
|  |  | Weighted mode | 2.92 (0.59 to 14.57) | 2.16 x 10^-1^ |
|  |  | MR Egger | 10.79 (0.26 to 443.04) | 2.35 x 10^-1^ |
| BMI | Fasting insulin | IVW | 0.17 (0.15 to 0.19) | 1.51 x 10^-74^ |
|  |  | Weighted median | 0.18 (0.15 to 0.21) | 1.89 x 10^-30^ |
|  |  | Weighted mode | 0.18 (0.13 to 0.24) | 6.25 x 10^-10^ |
|  |  | MR Egger | 0.20 (0.16 to 0.25) | 1.30 x 10^-16^ |

OR per natural log transformed pmol/L increase in fasting insulin and per SD (4.7 kg/m^2^) increase in BMI. BMI = body mass index. Effect estimate represents change in natural log transformed pmol/L fasting insulin.

### S18 Table. Results of female-specific SNP CRP sensitivity analysis MR.

| **Exposure** | **Outcome** | **Method** | **OR/Effect estimate (95% CI)** | ***P* value** |
| --- | --- | --- | --- | --- |
| CRP | Overall endometrial cancer | IVW | 1.11 (0.97 to 1.27) | 1.42 x 10^-1^ |
|  |  | Weighted median | 1.01 (0.87 to 1.18) | 8.77 x 10^-1^ |
|  |  | Weighted mode | 1.06 (0.93 to 1.20) | 3.89 x 10^-1^ |
|  |  | MR Egger | 1.00 (0.82 to 1.21) | 9.75 x 10^-1^ |
|  | Endometrioid endometrial cancer | IVW | 1.17 (0.99 to 1.38) | 6.04 x 10^-2^ |
|  |  | Weighted median | 1.04 (0.87 to 1.25) | 6.70 x 10^-1^ |
|  |  | Weighted mode | 1.08 (0.93 to 1.24) | 3.14 x 10^-1^ |
|  |  | MR Egger | 1.04 (0.82 to 1.31) | 7.57 x 10^-1^ |

OR per increase in natural log transformed mg/L CRP. Effect estimate represents change in natural log transformed mg/L CRP. CRP = C-reactive protein, BMI = body mass index, IVW= inverse-variance weighted.

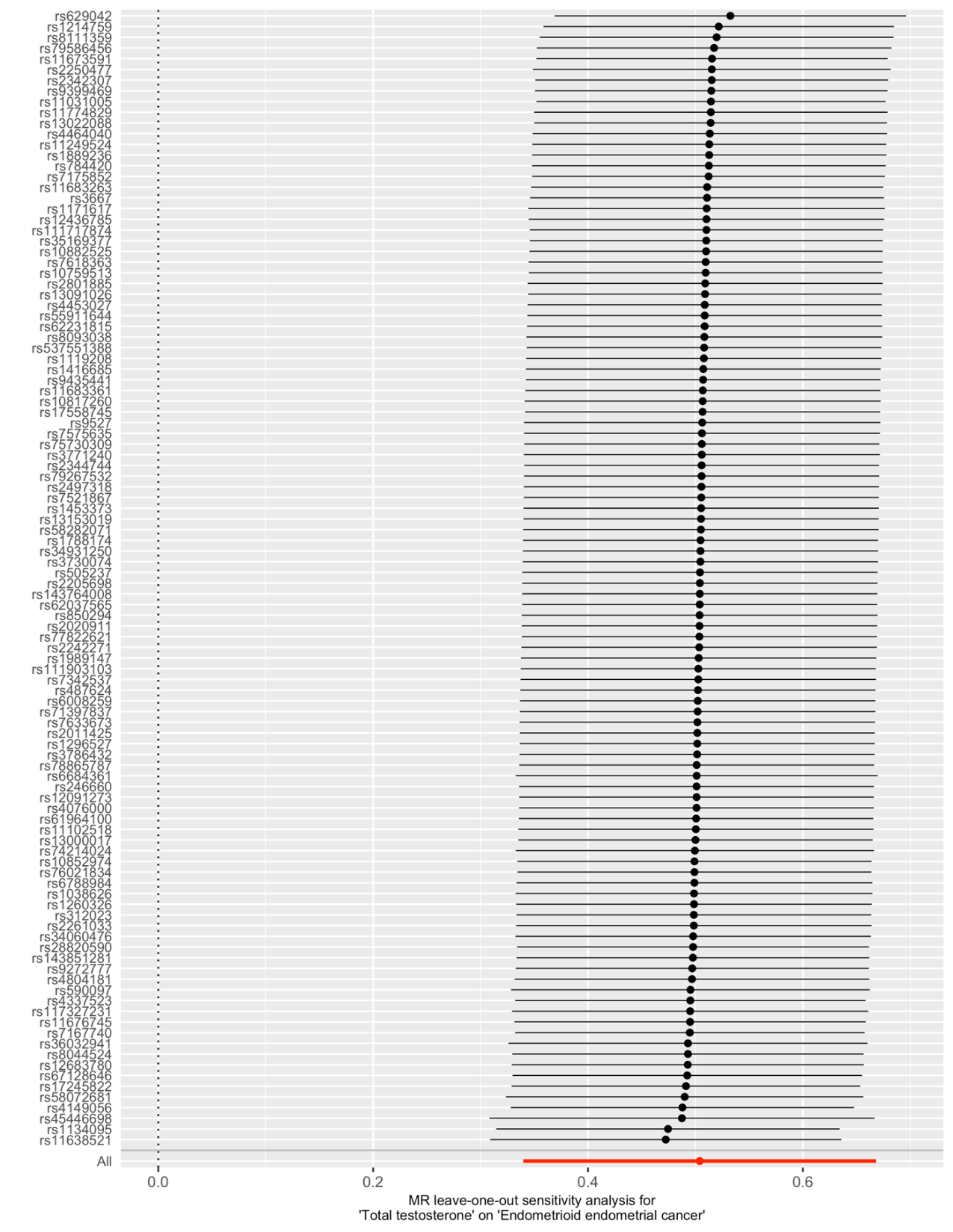

### S19 Figure. Leave-one-out analysis for MR examining the effect of total testosterone level on endometrioid endometrial cancer risk.

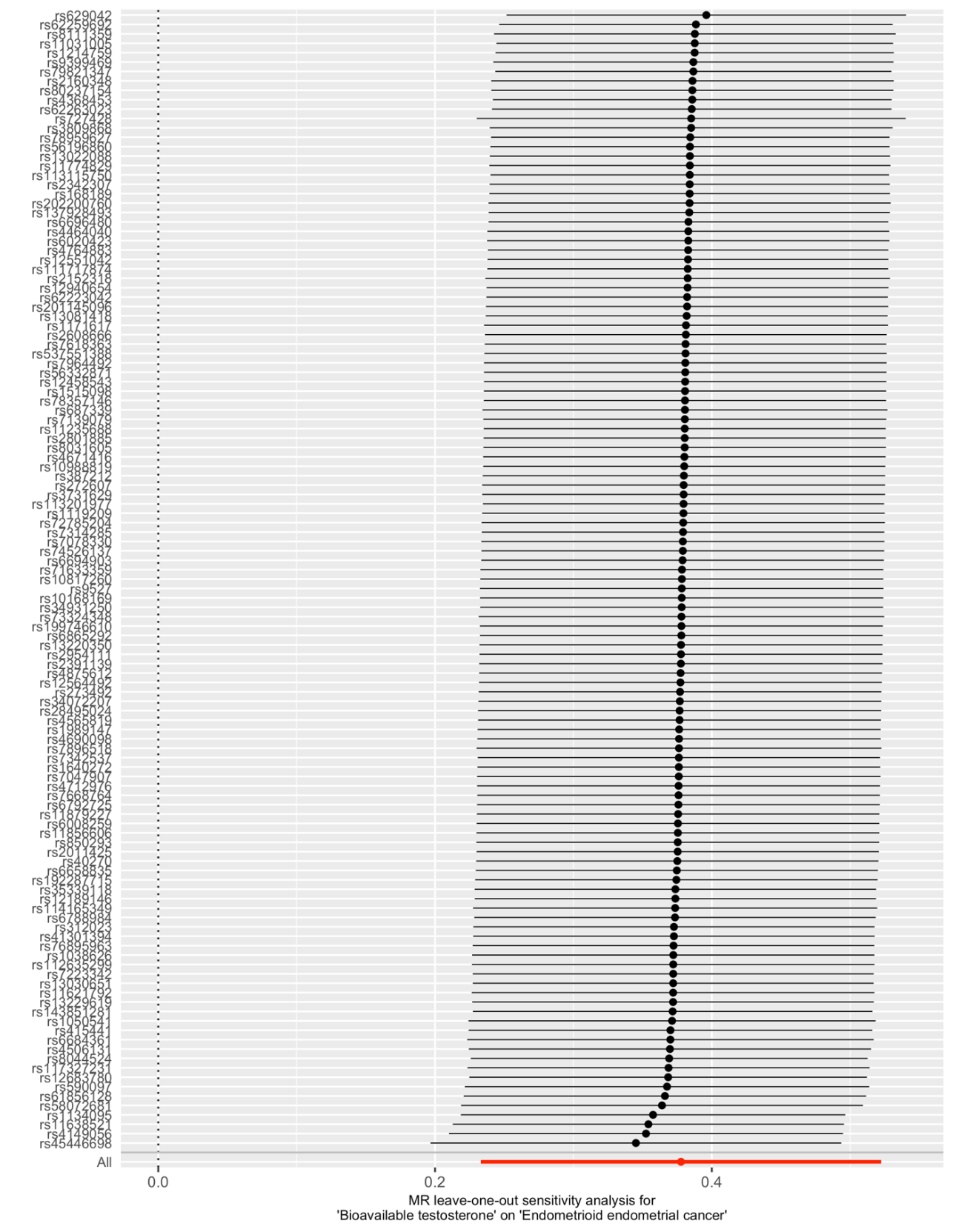

### S20 Figure. Leave-one-out analysis for MR examining the effect of bioavailable testosterone level on endometrioid endometrial cancer risk.

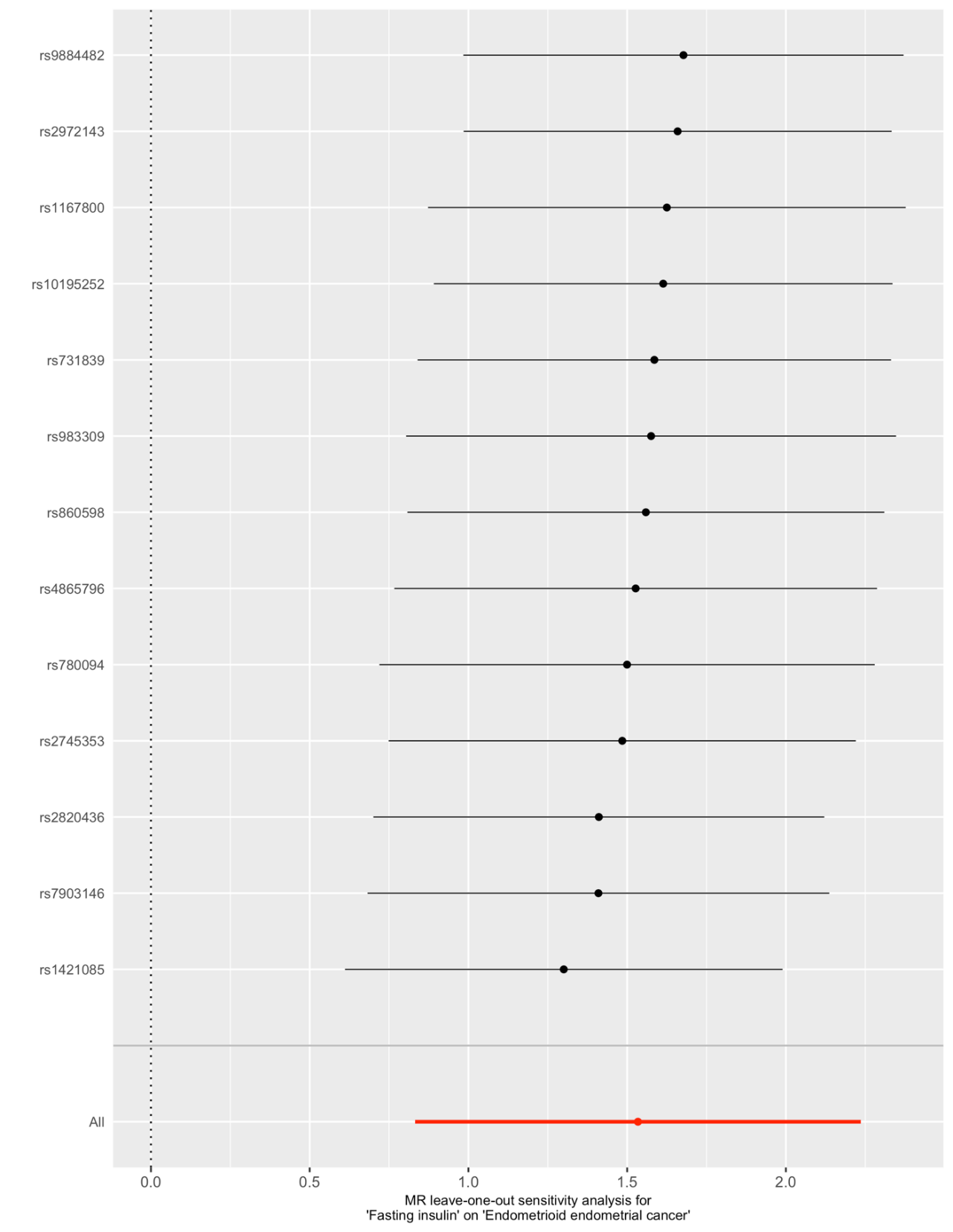

### S21 Figure. Leave-one-out analysis for MR examining the effect of fasting insulin level on endometrioid endometrial cancer risk.

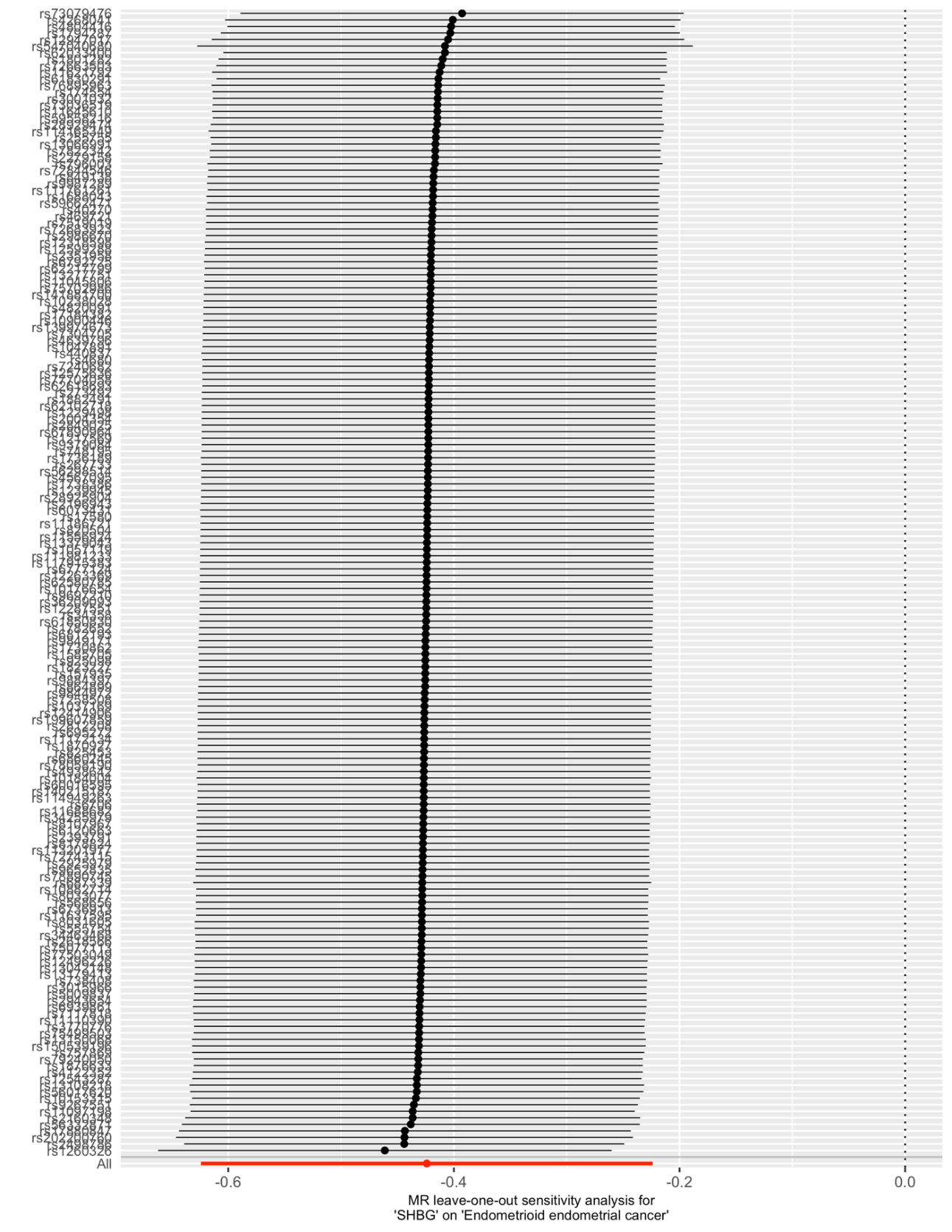

### S22 Figure. Leave-one-out analysis for MR examining the effect of SHBG level on endometrioid endometrial cancer risk.

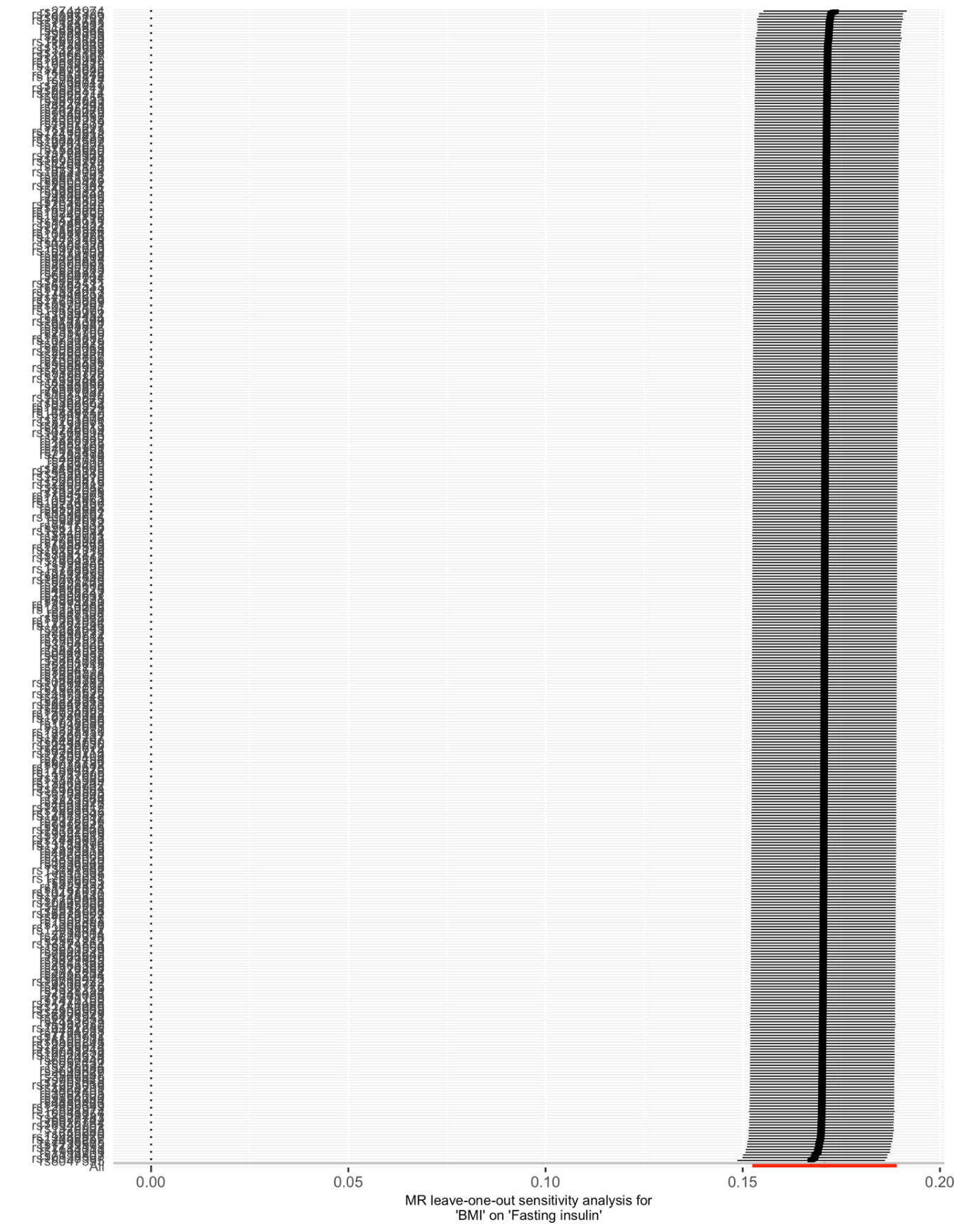

### S23 Figure. Leave-one-out analysis for MR examining the effect of adult BMI on fasting insulin level.

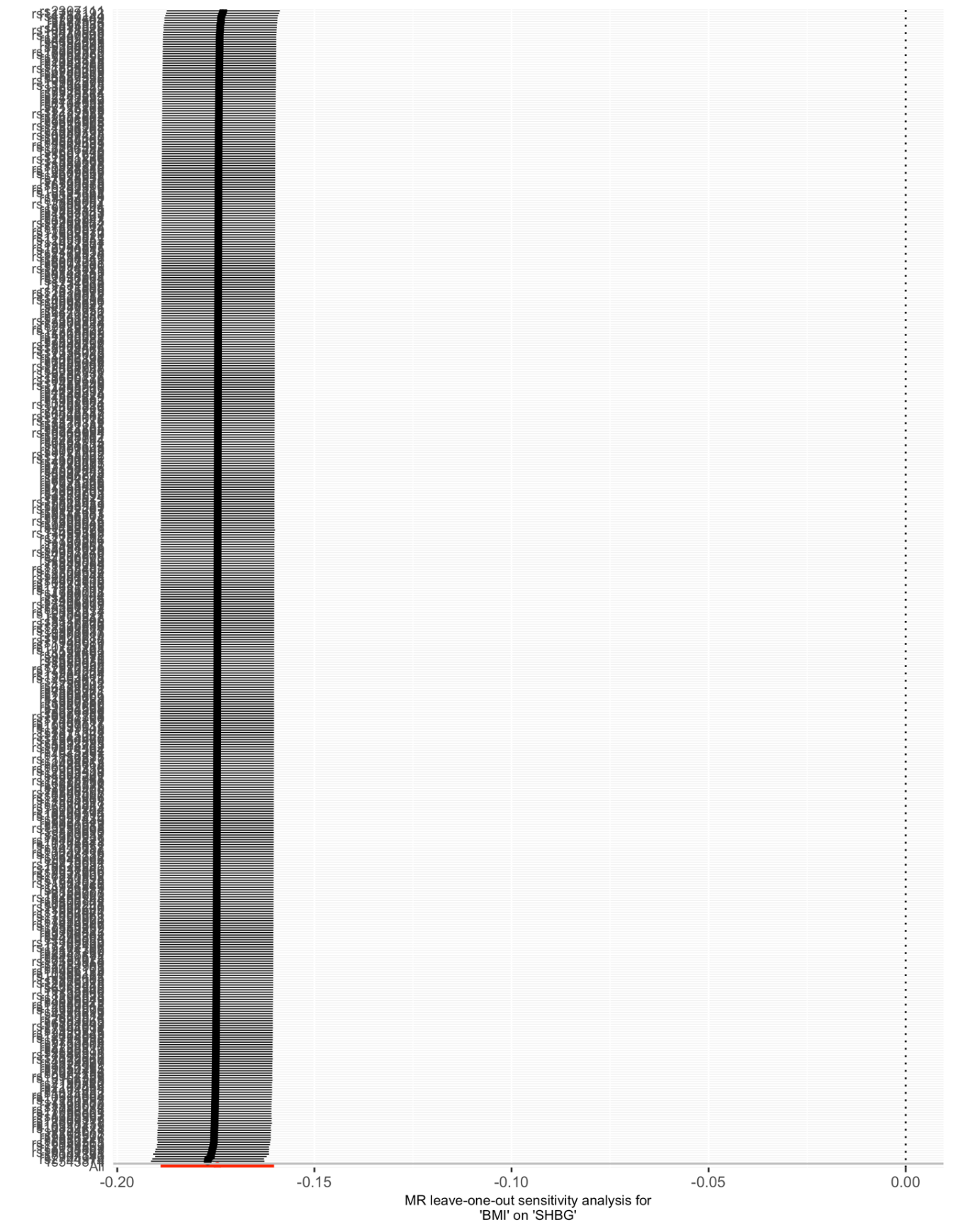

### S24 Figure. Leave-one-out analysis for MR examining the effect of adult BMI on SHBG level.

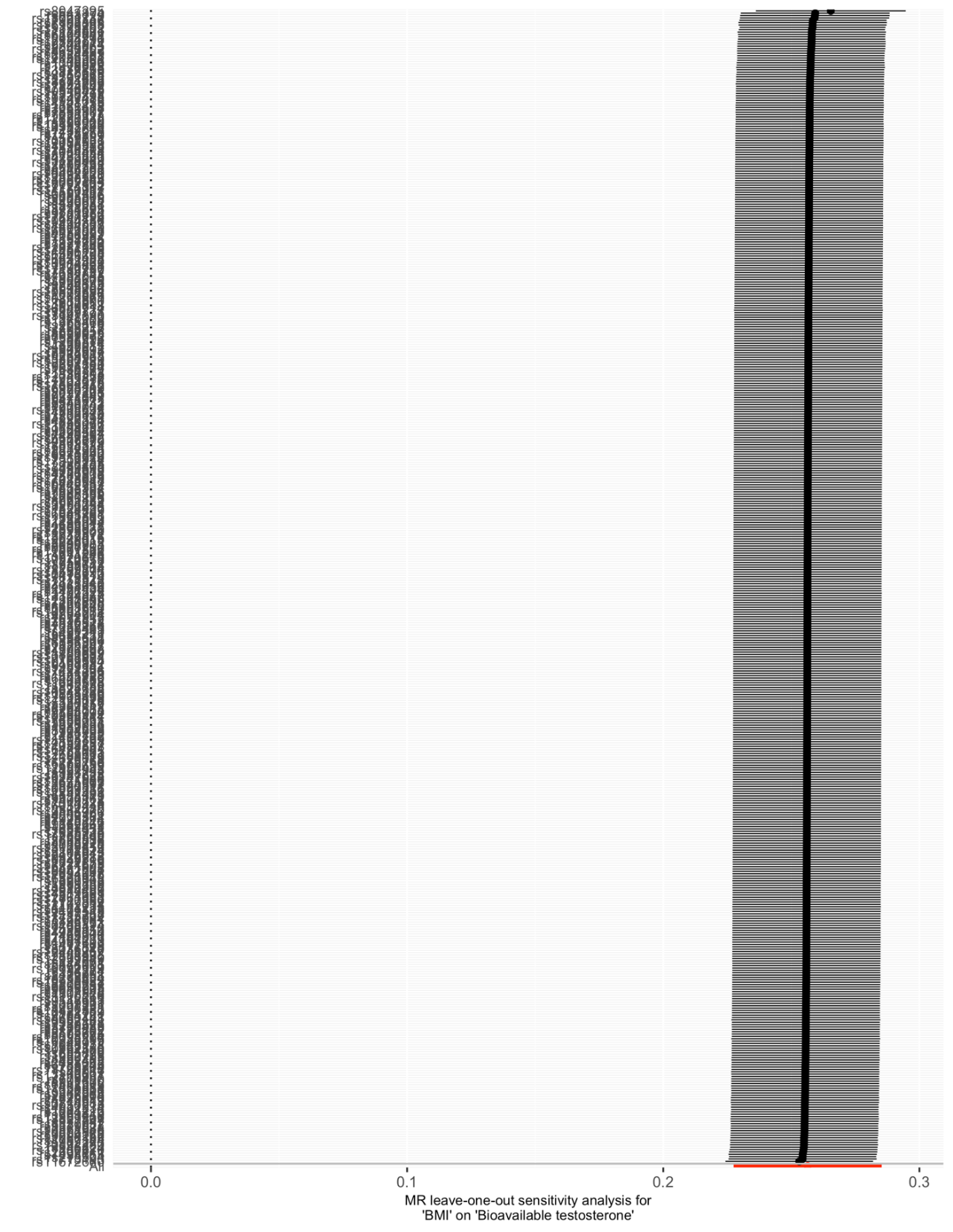

### S25 Figure. Leave-one-out analysis for MR examining the effect of adult BMI on bioavailable testosterone level.

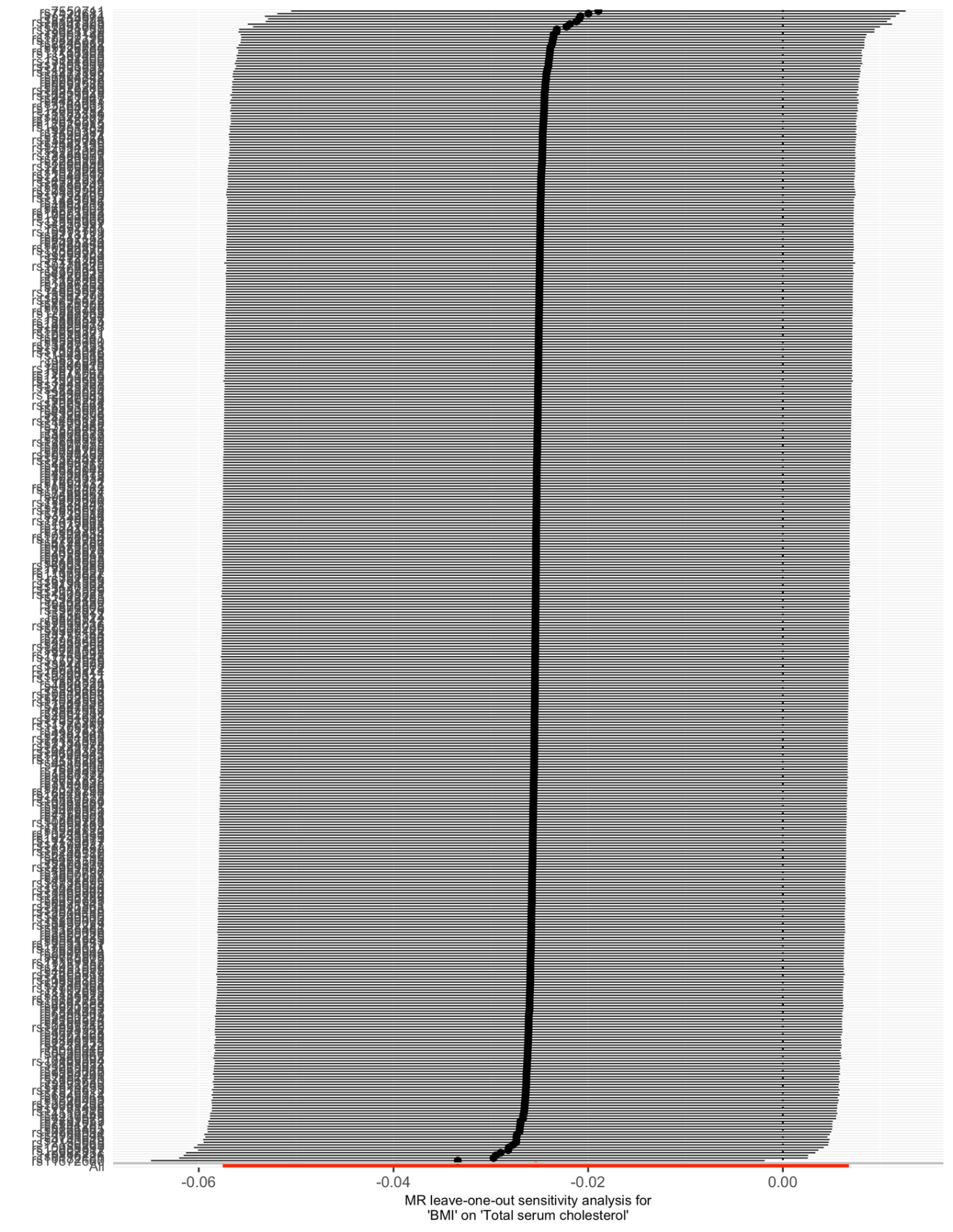

### S26 Figure. Leave-one-out analysis for MR examining the effect of adult BMI on total serum cholesterol level.

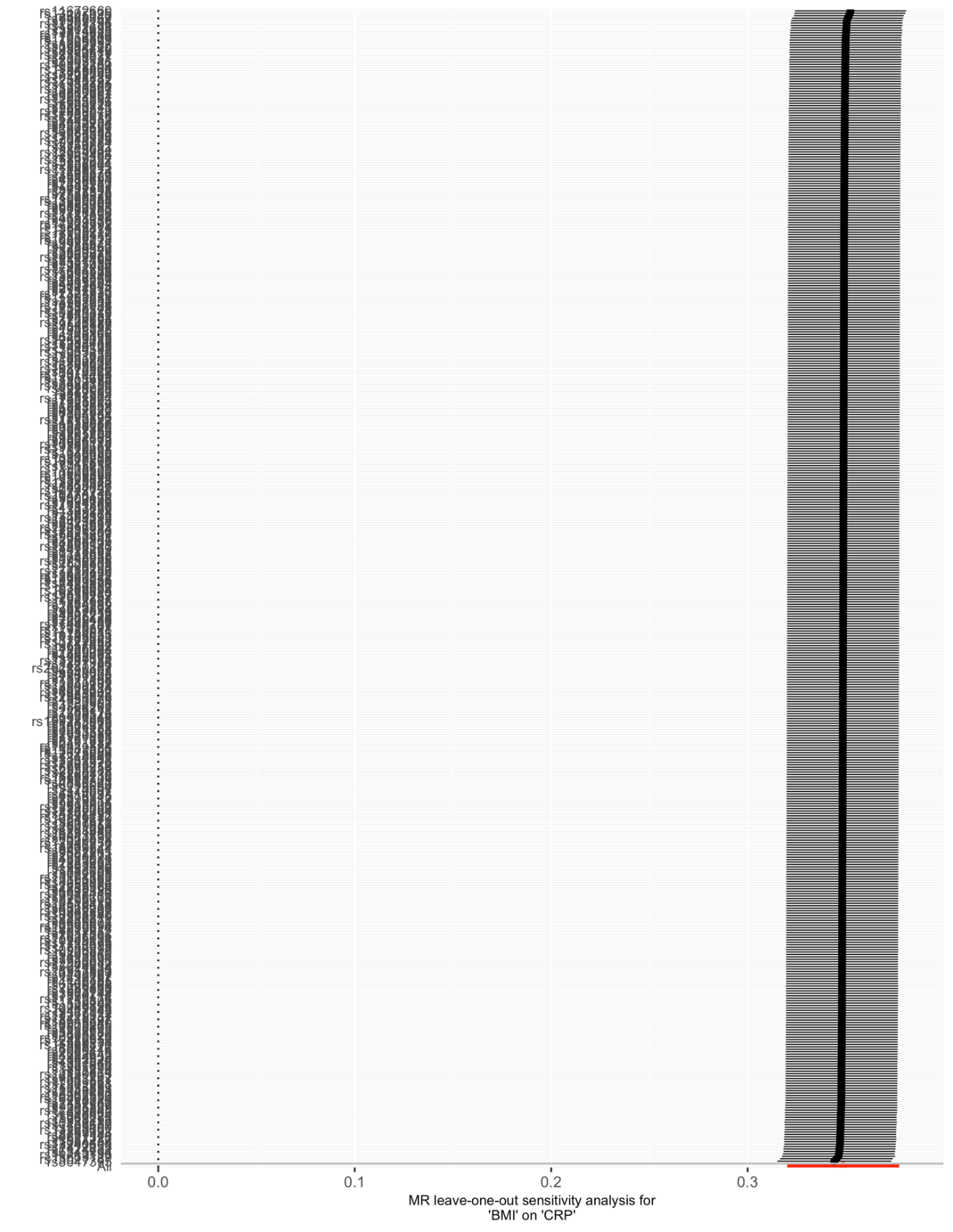

### S27 Figure. Leave-one-out analysis for MR examining the effect of adult BMI on C-reactive protein level.

### S28 Table. Results of female-specific SNP BMI sensitivity mediation analysis.

| **Mediator** | **Outcome** | **Direct Effect of BMI on Outcome** | **Indirect Effect of Mediator on Outcome** | **% Mediated (95% CI)** | ***P* value** |
| --- | --- | --- | --- | --- | --- |
| Fasting insulin | Overall endometrial cancer | 1.42 | 1.23 | 37% (19 to 56%) | 6.58 x 10^-5^ |
|  | Endometrioid endometrial cancer | 1.40 | 1.26 | 41% (19 to 63%) | 3.26 x 10^-4^ |
| Bioavailable testosterone | Overall endometrial cancer | 1.65 | 1.06 | 11% (5 to 16%) | 1.79 x 10^-4^ |
|  | Endometrioid endometrial cancer | 1.65 | 1.06 | 11% (5 to 17%) | 2.49 x 10^-4^ |
| SHBG | Overall endometrial cancer | 1.70 | 1.03 | 5% (1 to 10%) | 2.63 x 10^-2^ |
|  | Endometrioid endometrial cancer | 1.68 | 1.04 | 8% (3 to 13%) | 3.38 x 10^-3^ |

Direct effect is defined as the remaining effect of the exposure (BMI) on the outcome (endometrial cancer risk) when the effect of the candidate mediator on the outcome has been adjusted for. Indirect effect is defined as the effect of the exposure (BMI) on the outcome (endometrial cancer risk) through the candidate mediator. SHBG = sex hormone-binding globulin.

### S29 Table. Results of female-specific SNP fasting insulin mediation analysis.

| **Mediator** | **Outcome** | **Direct Effect of BMI on Outcome** | **Indirect Effect of Mediator on Outcome** | **% Mediated (95% CI)** | ***P* value** |
| --- | --- | --- | --- | --- | --- |
| Fasting insulin | Overall endometrial cancer | 1.43 | 1.21 | 43% (23 to 63%) | 1.71 x 10^-5^ |
|  | Endometrioid endometrial cancer | 1.89 | 1.10 | 16% (4 to 27%) | 7.78 x 10^-3^ |

Direct effect is defined as the remaining effect of the exposure (BMI) on the outcome (endometrial cancer risk) when the effect of the candidate mediator on the outcome has been adjusted for. Indirect effect is defined as the effect of the exposure (BMI) on the outcome (endometrial cancer risk) through the candidate mediator.

### S30 Table. Results of multivariable MR mediation analysis examining the effect of BMI and endometrial cancer with fasting insulin as a potential mediator with BMI-adjusted fasting insulin instrument and 100 SNPs from BMI instrument.

| **BMI Instrument** | **Outcome** | **Direct Effect of BMI on Outcome** | **Indirect Effect of Mediator on Outcome** | **% Mediated (95% CI)** | ***P* value** |
| --- | --- | --- | --- | --- | --- |
| Top 100 SNPs | Overall endometrial cancer | 1.67 | 1.13 | 19% (5 to 34%) | 9.17 x 10^-3^ |
|  | Endometrioid endometrial cancer | 1.65 | 1.15 | 21% (5 to 38%) | 1.17 x 10^-2^ |
| Randomly selected 100 SNPs | Overall endometrial cancer | 1.65 | 1.14 | 21% (4 to 38%) | 1.44 x 10^-2^ |
|  | Endometrioid endometrial cancer | 1.63 | 1.16 | 24% (4 to 43%) | 1.82 x 10^-2^ |

Direct effect is defined as the remaining effect of the exposure (BMI) on the outcome (endometrial cancer risk) when the effect of the candidate mediator on the outcome has been adjusted for. Indirect effect is defined as the effect of the exposure (BMI) on the outcome (endometrial cancer risk) through the candidate mediator.

### S31 Table. Results from sensitivity analyses examining the influence of Winner’s curse on GWAS with overlapping samples in analyses determining the mediating role of traits in the relationship between BMI and endometrial cancer risk.

| **Mediator** | **Outcome** | **Direct Effect of BMI on Outcome** | **Indirect Effect of Mediator on Outcome** | **% Mediated (95% CI)** | ***P* value** |
| --- | --- | --- | --- | --- | --- |
| Fasting insulin | Overall Endometrial Cancer | 1.58 | 1.17 | 26% (4 to 47%) | 1.78 x 10^-2^ |
|  | Endometrioid Endometrial Cancer | 1.63 | 1.17 | 25% (4 to 45%) | 1.78 x 10^-2^ |
| Bioavailable testosterone | Overall Endometrial Cancer | 1.69 | 1.10 | 16% (10 to 21%) | 3.16 x 10^-8^ |
|  | Endometrioid Endometrial Cancer | 1.73 | 1.10 | 15% (9 to 21%) | 1.68 x 10^-6^ |
| SHBG | Overall Endometrial Cancer | 1.76 | 1.05 | 8% (2 to 14%) | 7.04 x 10^-3^ |
|  | Endometrioid Endometrial Cancer | 1.77 | 1.08 | 11% (5 to 18%) | 8.41 x 10^-4^ |

### S32 Table. Results from sensitivity analyses examining the influence of Winner’s curse on GWAS with overlapping samples in analyses determining the interdependent effects of mediators of the relationship between BMI and endometrial cancer risk.

| **Exposure** | **Mediators included in model** | **Outcome** | **OR (95% CI)** | ***P* value** |
| --- | --- | --- | --- | --- |
| Fasting insulin | Bioavailable testosterone | Overall endometrial cancer | 1.10 (0.38 to 3.22) | 8.57 x 10^-1^ |
|  |  | Endometrioid endometrial cancer | 0.74 (0.21 to 2.58) | 6.44 x 10^-1^ |
|  | SHBG | Overall endometrial cancer | 2.34 (1.39 to 3.95) | 1.85 x 10^-3^ |
|  |  | Endometrioid endometrial cancer | 2.48 (1.37 to 4.51) | 3.46 x 10^-3^ |
| Bioavailable testosterone | SHBG | Overall endometrial cancer | 1.52 (1.28 to 1.80) | 2.18 x 10^-6^ |
|  |  | Endometrioid endometrial cancer | 1.52 (1.26 to 1.84) | 2.20 x 10^-5^ |
|  | Fasting insulin | Overall endometrial cancer | 1.52 (1.29 to 1.78) | 2.28 x 10^-6^ |
|  |  | Endometrioid endometrial cancer | 1.56 (1.30 to 1.87) | 1.11 x 10^-5^ |
| SHBG | Bioavailable testosterone | Overall endometrial cancer | 1.08 (0.85 to 1.38) | 5.06 x 10^-1^ |
|  |  | Endometrioid endometrial cancer | 1.03 (0.79 to 1.35) | 8.12 x 10^-1^ |
|  | Fasting insulin | Overall endometrial cancer | 0.75 (0.58 to 0.96) | 2.50 x 10^-2^ |
|  |  | Endometrioid endometrial cancer | 0.65 (0.48 to 0.86) | 4.07 x 10^-3^ |

### S33 Table. Conditional F-statistics for further multivariable Mendelian randomization analyses.

| **Mediator(s)** | **Conditional F-statistics** |
| --- | --- |
| Fasting insulin and BMI (top 100 SNPs) | BMI: 14  Fasting insulin: 6 |
| Fasting insulin and BMI (randomly selected 100 SNPs) | BMI: 15  Fasting insulin: 5 |
| Fasting insulin (adjusted for BMI) and BMI | BMI: 48  Fasting insulin: 4 |
| Fasting insulin (adjusted for BMI) and BMI (top 100 SNPs) | BMI: 70  Fasting insulin: 9 |
| Fasting insulin (adjusted for BMI) and BMI (randomly selected 100 SNPs) | BMI: 54  Fasting insulin: 8 |
| Fasting insulin (adjusted for BMI), SHBG, bioavailable testosterone, BMI | BMI: 38  Fasting insulin: 4  SHBG: 8  Bioavailable testosterone: 5 |
| Fasting insulin (adjusted for BMI), SHBG, BMI | BMI: 32  Fasting insulin: 4  SHBG: 20 |
| SHBG, bioavailable testosterone, BMI | BMI: 48  SHBG: 8  Bioavailable testosterone: 5 |
| Fasting insulin (adjusted for BMI), bioavailable testosterone, BMI | BMI: 28  Fasting insulin: 3  Bioavailable testosterone: 12 |
| Fasting insulin (adjusted for BMI), SHBG, bioavailable testosterone, BMI (top 100 SNPs) | BMI: 52  Fasting insulin: 6  SHBG: 14  Bioavailable testosterone: 9 |
| Fasting insulin (adjusted for BMI), SHBG, bioavailable testosterone, BMI (randomly selected 100 SNPs) | BMI: 41  Fasting insulin: 6  SHBG: 14  Bioavailable testosterone: 9 |
| Fasting insulin (adjusted for BMI), SHBG, BMI (top 100 SNPs) | BMI: 52  Fasting insulin: 7  SHBG: 40 |
| Fasting insulin (adjusted for BMI), SHBG, BMI (randomly selected 100 SNPs) | BMI: 43  Fasting insulin:7  SHBG: 40 |
| SHBG, bioavailable testosterone, BMI (top 100 SNPs) | BMI: 67  SHBG: 15  Bioavailable testosterone: 9 |
| SHBG, bioavailable testosterone, BMI (randomly selected 100 SNPs) | BMI: 41  SHBG: 15  Bioavailable testosterone: 9 |
| Fasting insulin (adjusted for BMI), bioavailable testosterone, BMI (top 100 SNPs) | BMI: 32  Fasting insulin: 4  Bioavailable testosterone: 21 |
| Fasting insulin (adjusted for BMI), bioavailable testosterone, BMI (randomly selected 100 SNPs) | BMI: 30  Fasting insulin: 3  Bioavailable testosterone: 19 |
| Fasting insulin (adjusted for BMI), SHBG, bioavailable testosterone | Fasting insulin: 8  SHBG: 21  Bioavailable testosterone: 13 |
| Fasting insulin (adjusted for BMI), SHBG | Fasting insulin: 10  SHBG: 72 |
| SHBG, bioavailable testosterone | SHBG: 15  Bioavailable testosterone: 15 |
| Fasting insulin (adjusted for BMI), bioavailable testosterone | Fasting insulin: 5  Bioavailable testosterone: 64 |

BMI = body mass index, SHBG = sex hormone-binding globulin.

### S34 Table. Results of multivariable MR mediation analysis examining the effect of endometrial cancer with pairs of confirmed mediating molecular traits without BMI.

| **Exposure** | **Mediators included in model** | **Outcome** | **OR (95% CI)** | ***P* value** |
| --- | --- | --- | --- | --- |
| Fasting insulin | Bioavailable testosterone | Overall endometrial cancer | 1.22 (0.48 to 3.11) | 6.78 x 10^-1^ |
|  |  | Endometrioid endometrial cancer | 1.05 (0.36 to 3.03) | 9.33 x 10^-1^ |
|  | SHBG | Overall endometrial cancer | 2.28 (1.34 to 3.86) | 2.85 x 10^-3^ |
|  |  | Endometrioid endometrial cancer | 3.74 (0.74 to 19.01) | 1.56 x 10^-1^ |
| Bioavailable testosterone | SHBG | Overall endometrial cancer | 1.51 (1.29 to 1.78) | 7.38 x 10^-7^ |
|  |  | Endometrioid endometrial cancer | 1.56 (1.27 to 1.90) | 2.82 x 10^-5^ |
|  | Fasting insulin | Overall endometrial cancer | 1.54 (1.32 to 1.78) | 2.55 x 10^-7^ |
|  |  | Endometrioid endometrial cancer | 1.57 (1.32 to 1.86) | 2.03 x 10^-6^ |
| SHBG | Bioavailable testosterone | Overall endometrial cancer | 1.08 (0.86 to 1.36) | 5.00 x 10^-1^ |
|  |  | Endometrioid endometrial cancer | 1.16 (0.81 to 1.65) | 4.12 x 10^-1^ |
|  | Fasting insulin | Overall endometrial cancer | 0.76 (0.59 to 0.97) | 2.70 x 10^-2^ |
|  |  | Endometrioid endometrial cancer | 0.99 (0.47 to 2.11) | 9.89 x 10^-1^ |

ORs are shown per increase in inverse normal transformed nmol/L SHBG, natural log transformed pmol/L fasting insulin, inverse normal transformed nmol/L bioavailable testosterone, SD (4.7 kg/m^2^) BMI. BMI = body mass index, OR = odds ratio, SHBG = sex hormone-binding globulin.

### S35 Table. Results of multivariable MR mediation analysis examining the effect of BMI and endometrial cancer with fasting insulin as a potential mediator with 100 SNPs from BMI instrument.

| **BMI Instrument** | **Outcome** | **Direct Effect of BMI on Outcome** | **Indirect Effect of Mediator on Outcome** | **% Mediated (95% CI)** | ***P* value** |
| --- | --- | --- | --- | --- | --- |
| Top 100 SNPs | Overall endometrial cancer | 1.62 | 1.16 | 24% (11 to 38%) | 4.44 x 10^-4^ |
|  | Endometrioid endometrial cancer | 1.62 | 1.17 | 25% (10 to 41%) | 1.38 x 10^-3^ |
| Randomly selected 100 SNPs | Overall endometrial cancer | 1.63 | 1.15 | 22% (6 to 37%) | 7.59 x 10^-3^ |
|  | Endometrioid endometrial cancer | 1.65 | 1.15 | 22% (3 to 40%) | 2.25 x 10^-2^ |

Direct effect is defined as the remaining effect of the exposure (BMI) on the outcome (endometrial cancer risk) when the effect of the candidate mediator on the outcome has been adjusted for. Indirect effect is defined as the effect of the exposure (BMI) on the outcome (endometrial cancer risk) through the candidate mediator. BMI = body mass index.

### S36 Table. Results of multivariable MR mediation analysis examining the effect of BMI and endometrial cancer with fasting insulin as a potential mediator with BMI-adjusted fasting insulin instrument.

| **Outcome** | **Direct Effect of BMI on Outcome** | **Indirect Effect of Mediator on Outcome** | **% Mediated (95% CI)** | ***P* value** |
| --- | --- | --- | --- | --- |
| Overall endometrial cancer | 1.73 | 1.08 | 13% (2 to 24%) | 1.62 x 10^-2^ |
| Endometrioid endometrial cancer | 1.67 | 1.14 | 20% (6 to 34%) | 3.91 x 10^-3^ |

Direct effect is defined as the remaining effect of the exposure (BMI) on the outcome (endometrial cancer risk) when the effect of the candidate mediator on the outcome has been adjusted for. Indirect effect is defined as the effect of the exposure (BMI) on the outcome (endometrial cancer risk) through the candidate mediator. BMI = body mass index.

### S37 Table. Conditional F-statistics for fasting insulin (adjusted for BMI) and BMI with differing thresholds used to construct fasting insulin.

| **Trait** | ***P*-value** | **r^2^** | **Conditional F-statistics** |
| --- | --- | --- | --- |
| Fasting insulin | 5x10^-7^ | 0.001 | BMI: 12  Fasting insulin: 3 |
|  | 5x10^-7^ | 0.01 | BMI: 12  Fasting insulin: 3 |
|  | 5x10^-6^ | 0.001 | BMI: 29  Fasting insulin: 3 |
|  | 5x10^-6^ | 0.01 | BMI: 27  Fasting insulin: 3 |
| Fasting insulin (adjusted for BMI) | 5x10^-8^ | 0.001 | BMI: 48  Fasting insulin: 4 |
|  | 5x10^-8^ | 0.01 | BMI: 48  Fasting insulin: 4 |
|  | 5x10^-7^ | 0.001 | BMI: 49  Fasting insulin: 4 |
|  | 5x10^-7^ | 0.01 | BMI: 49  Fasting insulin: 4 |
|  | 5x10^-6^ | 0.001 | BMI: 49  Fasting insulin: 4 |
|  | 5x10^-6^ | 0.01 | BMI: 4  Fasting insulin: 49 |

BMI = body mass index.

### S38 Table. Results of multivariable MR mediation analysis examining the effect of BMI and endometrial cancer with all confirmed mediating molecular traits with BMI-adjusted fasting insulin.

| **Exposure** | **Outcome** | **OR (95% CI)** | ***P* value** |
| --- | --- | --- | --- |
| Fasting insulin | Overall endometrial cancer | 1.68 (1.16 to 2.41) | 5.56 x 10^-3^ |
|  | Endometrioid endometrial cancer | 1.84 (1.04 to 3.24) | 3.63 x 10^-2^ |
| Bioavailable testosterone | Overall endometrial cancer | 1.11 (0.93 to 1.31) | 2.36 x 10^-1^ |
|  | Endometrioid endometrial cancer | 1.10 (0.88 to 1.37) | 4.00 x 10^-1^ |
| SHBG | Overall endometrial cancer | 0.89 (0.69 to 1.14) | 3.55 x 10^-1^ |
|  | Endometrioid endometrial cancer | 0.86 (0.60 to 1.23) | 3.97 x 10^-1^ |

ORs are shown per increase in inverse normal transformed nmol/L SHBG, natural log transformed pmol/L fasting insulin, inverse normal transformed nmol/L bioavailable testosterone. BMI = body mass index, OR = odds ratio, SHBG = sex hormone-binding globulin.

### S39 Table. Results of multivariable MR mediation analysis examining the effect of BMI and endometrial cancer with pairs of confirmed mediating molecular traits.

| **Exposure** | **Mediators included in model** | **Outcome** | **OR (95% CI)** | ***P* value** |
| --- | --- | --- | --- | --- |
| Fasting insulin | Bioavailable testosterone, BMI | Overall endometrial cancer | 1.26 (0.81 to 1.96) | 3.10 x 10^-1^ |
|  |  | Endometrioid endometrial cancer | 1.61 (0.93 to 2.80) | 8.94 x 10^-2^ |
|  | SHBG, BMI | Overall endometrial cancer | 1.68 (1.18 to 2.40) | 4.45 x 10^-3^ |
|  |  | Endometrioid endometrial cancer | 2.28 (1.29 to 4.03) | 4.66 x 10^-3^ |
| Bioavailable testosterone | SHBG, BMI | Overall endometrial cancer | 1.53 (1.30 to 1.80) | 4.06 x 10^-7^ |
|  |  | Endometrioid endometrial cancer | 1.55 (1.26 to 1.90) | 4.19 x 10^-5^ |
|  | Fasting insulin, BMI | Overall endometrial cancer | 1.43 (1.27 to 1.62) | 9.06 x 10^-9^ |
|  |  | Endometrioid endometrial cancer | 1.44 (1.24 to 1.67) | 2.22 x 10^-6^ |
| SHBG | Bioavailable testosterone, BMI | Overall endometrial cancer | 1.16 (0.91 to 1.48) | 2.24 x 10^-1^ |
|  |  | Endometrioid endometrial cancer | 1.17 (0.83 to 1.65) | 3.72 x 10^-1^ |
|  | Fasting insulin, BMI | Overall endometrial cancer | 0.79 (0.65 to 0.97) | 2.33 x 10^-2^ |
|  |  | Endometrioid endometrial cancer | 0.85 (0.55 to 1.32) | 4.77 x 10^-1^ |

ORs are shown per increase in inverse normal transformed nmol/L SHBG, natural log transformed pmol/L fasting insulin, inverse normal transformed nmol/L bioavailable testosterone, SD (4.7 kg/m^2^) BMI. BMI = body mass index, SHBG = sex hormone-binding globulin.

### S40 Table. Results of multivariable MR mediation analysis examining the effect of BMI and endometrial cancer with all confirmed mediating molecular traits including fasting insulin adjusted for BMI with 100 SNPs from BMI instrument.

| **BMI instrument** | **Exposure** | **Outcome** | **OR (95% CI)** | ***P* value** |
| --- | --- | --- | --- | --- |
| Top 100 SNPs | Fasting insulin | Overall endometrial cancer | 1.98 (1.26 to 3.11) | 3.36 x 10^-3^ |
|  |  | Endometrioid endometrial cancer | 1.58 (0.68 to 3.70) | 2.92 x 10^-1^ |
|  | Bioavailable testosterone | Overall endometrial cancer | 1.08 (0.88 to 1.31) | 4.63 x 10^-1^ |
|  |  | Endometrioid endometrial cancer | 1.13 (0.89 to 1.45) | 3.22 x 10^-1^ |
|  | SHBG | Overall endometrial cancer | 0.86 (0.65 to 1.15) | 3.13 x 10^-1^ |
|  |  | Endometrioid endometrial cancer | 0.90 (0.59 to 1.37) | 6.24 x 10^-1^ |
| Randomly selected 100 SNPs | Fasting insulin | Overall endometrial cancer | 2.07 (1.29 to 3.35) | 2.98 x 10^-3^ |
|  |  | Endometrioid endometrial cancer | 1.73 (0.66 to 4.50) | 2.65 x 10^-1^ |
|  | Bioavailable testosterone | Overall endometrial cancer | 1.07 (0.87 to 1.31) | 5.22 x 10^-1^ |
|  |  | Endometrioid endometrial cancer | 1.10 (0.85 to 1.44) | 4.61 x 10^-1^ |
|  | SHBG | Overall endometrial cancer | 0.86 (0.64 to 1.16) | 3.26 x 10^-1^ |
|  |  | Endometrioid endometrial cancer | 0.87 (0.55 to 1.38) | 5.62 x 10^-1^ |

ORs are shown per increase in inverse normal transformed nmol/L SHBG, natural log transformed pmol/L fasting insulin, inverse normal transformed nmol/L bioavailable testosterone, SD (4.7 kg/m^2^) BMI. BMI = body mass index, SHBG = sex hormone-binding globulin.

### S41 Table. Results of multivariable MR mediation analysis examining the effect of BMI and endometrial cancer with pairs of confirmed mediating molecular traits with 100 SNPs from BMI instrument.

| **BMI instrument** | **Exposure** | **Mediators included in model** | **Outcome** | **OR (95% CI)** | ***P* value** |
| --- | --- | --- | --- | --- | --- |
| Top 100 SNPs | Fasting insulin | Bioavailable testosterone, BMI | Overall endometrial cancer | 1.16 (0.59 to 2.27) | 6.61 x 10^-1^ |
|  |  |  | Endometrioid endometrial cancer | 1.18 (0.55 to 2.55) | 6.77 x 10^-1^ |
|  |  | SHBG, BMI | Overall endometrial cancer | 1.99 (1.30 to 3.05) | 1.80 x 10^-3^ |
|  |  |  | Endometrioid endometrial cancer | 2.96 (1.30 to 6.74) | 1.11 x 10^-2^ |
|  | Bioavailable testosterone | SHBG, BMI | Overall endometrial cancer | 1.58 (1.32 to 1.90) | 1.69 x 10^-6^ |
|  |  |  | Endometrioid endometrial cancer | 1.68 (1.35 to 2.09) | 6.49 x 10^-6^ |
|  |  | Fasting insulin, BMI | Overall endometrial cancer | 1.48 (1.29 to 1.70) | 5.89 x 10^-8^ |
|  |  |  | Endometrioid endometrial cancer | 1.51 (1.29 to 1.76) | 7.37 x 10^-7^ |
|  | SHBG | Bioavailable testosterone, BMI | Overall endometrial cancer | 1.20 (0.92 to 1.58) | 1.84 x 10^-1^ |
|  |  |  | Endometrioid endometrial cancer | 1.31 (0.90 to 1.92) | 1.64 x 10^-1^ |
|  |  | Fasting insulin, BMI | Overall endometrial cancer | 0.80 (0.64 to 1.00) | 4.80 x 10^-2^ |
|  |  |  | Endometrioid endometrial cancer | 1.22 (0.64 to 2.34) | 5.53 x 10^-1^ |
| Randomly selected 100 SNPs | Fasting insulin | Bioavailable testosterone, BMI | Overall endometrial cancer | 1.07 (0.49 to 2.32) | 8.68 x 10^-1^ |
|  |  |  | Endometrioid endometrial cancer | 1.05 (0.43 to 2.58) | 9.09 x 10^-1^ |
|  |  | SHBG, BMI | Overall endometrial cancer | 2.08 (1.32 to 3.29) | 1.92 x 10^-3^ |
|  |  |  | Endometrioid endometrial cancer | 3.58 (1.31 to 9.78) | 1.42 x 10^-2^ |
|  | Bioavailable testosterone | SHBG, BMI | Overall endometrial cancer | 1.58 (1.31 to 1.91) | 3.92 x 10^-6^ |
|  |  |  | Endometrioid endometrial cancer | 1.65 (1.30 to 2.09) | 4.80 x 10^-5^ |
|  |  | Fasting insulin, BMI | Overall endometrial cancer | 1.49 (1.29 to 1.72) | 1.60 x 10^-7^ |
|  |  |  | Endometrioid endometrial cancer | 1.51 (1.28 to 1.78) | 3.14 x 10^-6^ |
|  | SHBG | Bioavailable testosterone, BMI | Overall endometrial cancer | 1.19 (0.90 to 1.58) | 2.18 x 10^-1^ |
|  |  |  | Endometrioid endometrial cancer | 1.26 (0.84 to 1.90) | 2.71 x 10^-1^ |
|  |  | Fasting insulin, BMI | Overall endometrial cancer | 0.80 (0.63 to 1.01) | 6.33 x 10^-2^ |
|  |  |  | Endometrioid endometrial cancer | 1.26 (0.59 to 2.69) | 5.51 x 10^-1^ |

ORs are shown per increase in inverse normal transformed nmol/L SHBG, natural log transformed pmol/L fasting insulin, inverse normal transformed nmol/L bioavailable testosterone, SD (4.7 kg/m^2^) BMI. BMI = body mass index, SHBG = sex hormone-binding globulin.

### S42 Table. Results of multivariable MR mediation analysis examining the effect of endometrial cancer with all confirmed mediating molecular traits without BMI.

| **Exposure** | **Outcome** | **OR (95% CI)** | ***P* value** |
| --- | --- | --- | --- |
| Fasting insulin | Overall endometrial cancer | 2.07 (1.20 to 3.55) | 9.23 x 10^-3^ |
|  | Endometrioid endometrial cancer | 1.60 (0.44 to 5.87) | 4.79 x 10^-1^ |
| Bioavailable testosterone | Overall endometrial cancer | 1.09 (0.88 to 1.37) | 4.31 x 10^-1^ |
|  | Endometrioid endometrial cancer | 1.14 (0.83 to 1.54) | 4.21 x 10^-1^ |
| SHBG | Overall endometrial cancer | 0.87 (0.63 to 1.20) | 4.07 x 10^-1^ |
|  | Endometrioid endometrial cancer | 0.85 (0.50 to 1.45) | 5.63 x 10^-1^ |

ORs are shown per increase in inverse normal transformed nmol/L SHBG, natural log transformed pmol/L fasting insulin, inverse normal transformed nmol/L bioavailable testosterone. BMI = body mass index, SHBG = sex hormone-binding globulin.

### S43 Table. Results of sensitivity analysis examining the effect of only including 100 SNPs for the BMI instrument in MR analyses.

| **BMI instrument** | **Outcome** | **Method** | **OR (95% CI)** | ***P* value** |
| --- | --- | --- | --- | --- |
| Top 100 SNPs | Overall endometrial cancer | IVW | 1.94 (1.68 to 2.24) | 6.03 x 10^-19^ |
|  |  | Weighted median | 1.92 (1.53 to 2.40) | 1.23 x 10^-8^ |
|  |  | Weighted mode | 1.95 (1.37 to 2.77) | 3.12 x 10^-4^ |
|  |  | MR Egger | 1.97 (1.33 to 2.92) | 9.91 x 10^-4^ |
|  | Endometrioid endometrial cancer | IVW | 1.95 (1.64 to 2.30) | 1.01 x 10^-14^ |
|  |  | Weighted median | 1.93 (1.48 to 2.52) | 1.53 x 10^-6^ |
|  |  | Weighted mode | 1.77 (1.21 to 2.61) | 4.48 x 10^-3^ |
|  |  | MR Egger | 1.94 (1.24 to 3.05) | 4.77 x 10^-3^ |
|  | Fasting insulin | IVW | 1.21 (1.17 to 1.25) | 2.41 x 10^-27^ |
|  |  | Weighted median | 1.21 (1.17 to 1.26) | 3.78 x 10^-22^ |
|  |  | Weighted mode | 1.20 (1.13 to 1.28) | 6.06 x 10^-8^ |
|  |  | MR Egger | 1.21 (1.10 to 1.32) | 1.02 x 10^-4^ |
|  | SHBG | IVW | 0.85 (0.83 to 0.88) | 6.83 x 10^-34^ |
|  |  | Weighted median | 0.87 (0.85 to 0.89) | 1.55 x 10^-31^ |
|  |  | Weighted mode | 0.86 (0.84 to 0.89) | 1.49 x 10^-15^ |
|  |  | MR Egger | 0.92 (0.86 to 0.98) | 1.82 x 10^-2^ |
|  | Bioavailable testosterone | IVW | 1.25 (1.20 to 1.31) | 4.93 x 10^-23^ |
|  |  | Weighted median | 1.18 (1.12 to 1.25) | 2.66 x 10^-10^ |
|  |  | Weighted mode | 1.10 (1.02 to 1.19) | 1.64 x 10^-2^ |
|  |  | MR Egger | 1.09 (0.97 to 1.23) | 1.31 x 10^-1^ |
| Randomly selected 100 SNPs | Overall endometrial cancer | IVW | 1.75 (1.39 to 2.19) | 1.89 x 10^-6^ |
|  |  | Weighted median | 1.72 (1.23 to 2.41) | 1.58 x 10^-3^ |
|  |  | Weighted mode | 1.68 (1.16 to 2.43) | 7.57 x 10^-3^ |
|  |  | MR Egger | 1.66 (1.02 to 2.70) | 4.31 x 10^-2^ |
|  | Endometrioid endometrial cancer | IVW | 1.88 (1.43 to 2.47) | 5.53 x 10^-6^ |
|  |  | Weighted median | 2.06 (1.40 to 3.05) | 2.86 x 10^-4^ |
|  |  | Weighted mode | 1.82 (1.22 to 2.72) | 4.43 x 10^-3^ |
|  |  | MR Egger | 1.70 (0.96 to 3.02) | 7.41 x 10^-2^ |
|  | Fasting insulin | IVW | 1.21 (1.16 to 1.25) | 6.68 x 10^-21^ |
|  |  | Weighted median | 1.22 (1.15 to 1.29) | 3.07 x 10^-11^ |
|  |  | Weighted mode | 1.21 (1.13 to 1.30) | 3.19 x 10^-7^ |
|  |  | MR Egger | 1.26 (1.17 to 1.37) | 1.51 x 10^-7^ |
|  | SHBG | IVW | 0.85 (0.83 to 0.87) | 5.94 x 10^-35^ |
|  |  | Weighted median | 0.86 (0.83 to 0.89) | 2.02 x 10^-19^ |
|  |  | Weighted mode | 0.86 (0.83 to 0.89) | 5.11 x 10^-14^ |
|  |  | MR Egger | 0.85 (0.80 to 0.90) | 1.10 x 10^-7^ |
|  | Bioavailable testosterone | IVW | 1.24 (1.17 to 1.31) | 3.48 x 10^-13^ |
|  |  | Weighted median | 1.14 (1.04 to 1.24) | 4.69 x 10^-3^ |
|  |  | Weighted mode | 1.10 (1.01 to 1.18) | 2.20 x 10^-2^ |
|  |  | MR Egger | 1.04 (0.93 to 1.17) | 4.67 x 10^-1^ |

ORs are shown per increase in inverse normal transformed nmol/L SHBG, natural log transformed pmol/L fasting insulin, inverse normal transformed nmol/L bioavailable testosterone, SD (4.7 kg/m^2^) BMI. BMI = body mass index, SHBG = sex hormone-binding globulin.

### S44 Table. Conditional F-statistics with different levels of genetic correlation for SHBG and BMI in multivariable Mendelian randomization analyses.

| **Mediator** | **Genetic correlation** | **Conditional F-statistics** |
| --- | --- | --- |
| SHBG | 0 | BMI: 45  SHBG: 23 |
| SHBG | 0.25 | BMI: 37  SHBG: 20 |
| SHBG | 0.5 | BMI: 32  SHBG: 19 |
| SHBG | 0.75 | BMI: 27  SHBG: 17 |
| SHBG | 1 | BMI: 24  SHBG: 16 |
| Bioavailable testosterone | 0 | BMI: 43  Bioavailable testosterone: 13 |
| Bioavailable testosterone | 0.25 | BMI: 55  Bioavailable testosterone: 14 |
| Bioavailable testosterone | 0.5 | BMI: 77  Bioavailable testosterone: 16 |
| Bioavailable testosterone | 0.75 | BMI: 127  Bioavailable testosterone: 17 |
| Bioavailable testosterone | 1 | BMI: 361  Bioavailable testosterone: 19 |

BMI = body mass index, SHBG = sex hormone-binding globulin.

1.	Sinnott-Armstrong N, Tanigawa Y, Amar D, Mars NJ, Aguirre M, Venkataraman GR, et al. Genetics of 38 blood and urine biomarkers in the UK Biobank. bioRxiv. 2019:660506. doi: 10.1101/660506.

2.	Kettunen J, Demirkan A, Würtz P, Draisma HH, Haller T, Rawal R, et al. Genome-wide study for circulating metabolites identifies 62 loci and reveals novel systemic effects of LPA. Nat Commun. 2016;7:11122. Epub 2016/03/24. doi: 10.1038/ncomms11122. PubMed PMID: 27005778.

3.	Ruth KS, Day FR, Tyrrell J, Thompson DJ, Wood AR, Mahajan A, et al. Using human genetics to understand the disease impacts of testosterone in men and women. Nature Medicine. 2020;26(2):252-+. doi: 10.1038/s41591-020-0751-5. PubMed PMID: WOS:000512529400005.

4.	Locke AE, Kahali B, Berndt SI, Justice AE, Pers TH, Felix R, et al. Genetic studies of body mass index yield new insights for obesity biology. Nature. 2015;518(7538):197-U401. doi: 10.1038/nature14177. PubMed PMID: WOS:000349190300031.

### S45 Table. Results of HEIDI test-filtered low-density lipoprotein (LDL) cholesterol and overall endometrial cancer MR.

| **Method** | **OR (95% CI)** | ***P* value** |
| --- | --- | --- |
| IVW | 0.93 (0.86 to 1.00) | 4.10 x 10^-2^ |
| Weighted median | 0.91 (0.82 to 1.02) | 9.88 x 10^-2^ |
| Weighted mode | 0.90 (0.81 to 1.00) | 5.66 x 10^-2^ |
| MR Egger | 0.91 (0.82 to 1.02) | 1.01 x 10^-1^ |

ORs are shown per increase in SD (38.7 mg/dL) LDL-cholesterol. IVW = inverse-variance weighted.
